# A digital twin for hospital antimicrobial resistance forecasting and constrained intervention optimisation

**DOI:** 10.64898/2026.05.15.26353296

**Authors:** Charalampos P. Triantafyllidis, Ricardo Aguas

## Abstract

Hospital antimicrobial resistance (AMR) emanates from an array of complex interactions between patient turnover, heterogeneous patient–staff contact patterns, antibiotic-driven within-host selection, and imperfect surveillance. We present a hospital AMR digital twin that combines mechanistic simulation with temporal graph learning to forecast resistance emergence from evolving daily contact networks and enable support intervention planning. Our approach is twofold: graph neural networks and transformers to model predictions and mathematical programming optimization to provide decision support. The main predictive task asks whether future spread of resistant infections is more likely to be driven by endogenous hospital transmission and selection or from importation on admission. We evaluated this task under both fully observed and partially observed settings, using baseline benchmarks together with ablations, surveillance perturbations, and distribution-shift stress tests. Under canonical conditions, the model achieved very strong predictive performance, especially when ground-truth system states were available, and remained informative under partial observation. Ablations showed that contact-weight information was relatively robust, whereas compressed node-feature representations weakened performance more noticeably when observations were incomplete. Surveillance stress tests further showed that delayed or less frequent reporting can be tolerated in some settings, but threshold calibration becomes fragile under more severe observation changes. Across broader epidemiological and surveillance shifts, the ground-truth model generally preserved strong ranking ability, while partial-observation performance was less stable. When models were trained directly in the shifted regime, performance improved compared with zero-shot transfer, indicating that the digital twin can adapt to new and previously unseen operating conditions but that portability across regimes can improve, particularly when only partial surveillance data are available. We also evaluated intervention-conditioned forecasting by branching hospital states into a small library of screening and isolation policies under a shock-and-superspreader regime. The learned models supported useful within-state action ranking and frequently identified policies that improved on the baseline containment protocols or avoided worsening outcomes. The same digital twin can also support constrained intervention selection, although reliable deployment will require careful calibration, improved robustness under partial observation, and broader policy libraries.

## Introduction

Hospital antimicrobial resistance (AMR) remains a challenge for modern global health systems. It threatens patient safety with increased morbidity and subsequent mortality, complicates treatment pathways, and places additional strain on already pressured healthcare systems and services worldwide [1, 2, 3, 4, 5, 6, 7]. Hospital processes that shape AMR are not static, but dynamically reflective of patients’ admissions, transfers, discharges, tests, isolations, and treatments. Patient’s probability of acquiring a nosocomial AMR infection is a function of cumulative potential exposures in the different hospital environments they experience, which are in turn dependent on the existence of environmental sources of contagion and contact with other co-located infectious patients or hospital staff. Staff shape the hospital fabric, connecting wards through repeated patient-facing and colleague-facing interactions. Furthermore, patients carrying bacterial infections can become hosts for de-novo generated mutants after exposure to antibiotics. In these settings, AMR is not simply a microbiological phenomenon; it is also a systems problem spanning transmission, treatment, observability, operational practice and decision making. This systems perspective matters because the questions facing infection prevention and control teams (IPC) are inherently temporal, and the solutions rely on a mechanistic understanding of the underlying sources of AMR spread. Statistical and mathematical models provide frameworks proposing to inform those solutions.

Recent work suggests that forecasting nosocomial AMR infection spread from routinely collected data is possible, but much of that literature still operates at the level of aggregate hospital signals [8, 9]. These approaches offer limited value of information to address questions such as: What can prevent an increase in resistance emergence over the coming week? To meaningfully inform such a decision making process one would have to address whether a potential resistance emergence is a reflection of importation on admission, transmission within the hospital, or within-host selection under antibiotic pressure. Also relevant would be to explore to what extent screening frequency and delayed test reporting modify what actionable information is available for decision making. These questions require models that track both the evolving contact structure of the hospital and the policy-dependent state of observation. Indeed, a growing body of empirical and modelling work has shown that hospital contact structure is heterogeneous, role-dependent, and epidemiologically consequential [10, 11, 12, 13, 14]. An ideal construct would then be one in which AMR is treated as a time-evolving relational process rather than as a purely aggregate signal. Digital twins offer a natural way to materialise this approach, offering computational environments that combine mechanistic understanding, heterogeneous data, and decision support [15, 16, 17, 18, 19]. In the present context, a hospital AMR digital twin should do more than reproduce outbreaks or generate synthetic data. It should preserve the mechanisms that matter clinically, generate trajectories under operationally motivated assumptions, and support prediction of outcomes that are interpretable to IPC teams.

In this work, the digital twin has two tightly connected components. First, a simulator generates daily directed contact graphs with ward structure, patient turnover, antibiotic exposure, colonisation and infection dynamics, screening, test reporting and other seasonal forcing mechanisms or super-spreaders. Second, a temporal graph model maps recent graph history to future AMR outcomes. The primary predictive endpoint studied here is deliberately mechanism-aware: whether future resistance emergence over the next time horizon is predominantly driven by endogenous transmission or importation. Throughout this work, endogenous emergence refers to nosocomial transmission plus within-host selection, whereas importation refers to resistant cases introduced on admission from the general population. This distinction is clinically meaningful. It does not capture the whole complexity of AMR biology, but it does align with a practical IPC question: are resistant infections mainly being brought in, or are they being amplified within the hospital?

To meaningfully inform policies aiming to control nosocomial transmission of AMR infections, we need to resolve which modulators of infection spread need to be tuned, given a dynamic set of complex data.Decisions need to be made on what is the best option out of a set of candidate intervention measures, having a partially observed set of time and ward specific information on the occurrence of cases in both patients and staff, and full knowledge of the context of screening frequency and isolation events,. Here, we explore if and how graph neural networks (GNNs) can inform such decisions in real-time. We expand on previous work [20, 21], by developing a framework of temporally evolving daily graphs that are integrated through a Transformer-based temporal encoder and a mathematical model of nosocomial infection. We demonstrate the limits of the mechanistic reproducibility of outbreak dynamics within hospitals, i.e., to what extent GNNs can predict how nosocomial infections will spread when informed on curtailed outbreak data. We then illustrate how the integrated framework, bridging learnings from temporal contact graphs and features with a mechanistic model of intervention dynamics accounting for a particular set of evaluation metrics, can be translated into a causal pathway optimisation tool supporting AMR policy decision making. Rather than aiming to fully resolve AMR forecasting or intervention planning in hospital settings, we seek to demonstrate that mechanism-aware prediction from evolving hospital contact networks is possible but carries known sources of fragility, such as minimum data requirements and contextual operational constraints. We thus present a framework that can support the realistic real-time design of both actionable AMR surveillance and intervention planning in hospital settings.

### Related work

This study spans hospital contact-network epidemiology, mechanistic modelling of transmission, digital twins and deep learning. Each of these areas is computationally polished, but they have often progressed in separate contexts. In the hospital infection and AMR-modelling literature, a consistent message is that contact structure is neither homogeneous nor incidental. English *et al*. showed that both contact patterns and spatial movement vary markedly across healthcare-worker roles, indicating that hospital mixing is structured by occupation rather being uniform [22]. Wilson-Aggarwal *et al*. further showed, using routinely collected door-access logs and electronic medical records, that healthcare-worker mobility and patient connectivity change over time and differ across clinical speciality [23]. In addition, Myall *et al*. found that dynamic patient-contact networks substantially improve prediction of hospital-onset COVID-19 infections [24], while Bridgen *et al*. used Bayesian inference to identify the importance of hospital structure and staff interactions in nosocomial transmission [25]. Taken together, these studies support a view of hospital transmission as role dependent, structured, and dynamic.

Wearable-sensor and network-reconstruction studies reinforce this inference. Hornbeck *et al*. used sensor data to show that heterogeneity in healthcare-worker movement and contact patterns can substantially alter infection diffusion [26]. Mastrandrea *et al*. reported that contact duration and structure among healthcare workers are highly uneven and strongly organised [27]. Furthermore, Illingworth *et al*. showed that a relatively small subset of cases can account for a disproportionate share of onward transmission in hospital settings [28]. Additionally, Rocha *et al*. demonstrated that patient mobility and contact heterogeneity create high-risk pathways for methicillin-resistant *Staphylococcus aureus* spread across hospital systems [29], while Darbon *et al*. highlighted the temporal structure of hospital contacts and the centrality of particular professional groups, especially nurses, in sustaining transmission opportunity [30]. Earlier work remains relevant as Tahir *et al*. showed that patient movement within hospitals shapes spread across locations [31], while Donker *et al*. demonstrated that transfer networks between hospitals influence regional surveillance and dissemination [32]. More recent work has moved toward finer-grained inference on hospital transmission pathways and asymptomatic carriage [33, 34, 35]. Collectively, this literature establishes that hospital transmission is relational, structured, and mechanistically important.

Our platform is aligned with that view, but differs in three ways. First, rather than analysing a single observed network or reconstructing a static movement graph, it generates a daily sequence of directed, weighted hospital contact graphs with evolving node sets, explicit patient and staff roles, ward structure, and both latent and observed surveillance states. Second, it couples simulation to supervised temporal graph learning rather than stopping at simulation or retrospective pathway inference. Third, it preserves mechanism-specific bookkeeping for importation, transmission, and antibiotic-driven selection, so that prediction targets can be defined in mechanistic rather than purely aggregate terms. This framing is also consistent with recent work on synthetic contact reconstruction and intervention analysis. Duval *et al*. showed that realistic synthetic temporal contacts can recover much of the structure missing from simpler baselines [36]. Our study extends that direction by building a programmable digital twin that can generate counterfactually varied hospital trajectories under user-defined surveillance and IPC assumptions, and then training predictive models on the resulting graph sequences. From the digital-twin perspective, recent reviews have emphasised the need to integrate mechanistic knowledge, data, and decision support, while noting that much of the healthcare literature remains focused on individual patients or narrow physiological systems [37, 38]. The present platform operates at a different level: it is not a physiological twin of a single patient, but a digital twin of a hospital AMR ecosystem in which admissions, contacts, treatment exposure, latent colonisation, observed surveillance state, and IPC interventions co-evolve.

On the machine-learning side, the modelling layer is related to dynamic-graph representation learning, but in a setting with a clearer operational interpretation. Recent surveys have highlighted challenges in dynamic graph learning, including scalability, heterogeneity, temporal sparsity, and limited domain-specific benchmarks [39]. Our formulation represents hospital workflows at discrete time points: each day is represented as a directed attributed graph, each daily graph is embedded through edge-aware message passing and learned graph pooling, and a Transformer aggregates graph embeddings across a finite observation window to account for temporal evolution and learning. This extends our earlier graph-learning work from static to temporal graphs [20]. Here, the model must learn not only structural regularities but also temporal ones, and the prediction targets are tied directly to future hospital dynamics.

The novelty therefore does not lie in any single ingredient. Hospital contact-network epidemiology, stochastic transmission modelling, digital-twin framing, and temporal graph learning all have established literatures. The contribution lies in bringing them together in a single framework with ward-structured daily graph generation, staff-mediated cross-ward connectivity, explicit latent and observed AMR infection states, imperfect screening and delayed reporting, dynamic patient turnover with importation on admission, mechanism-resolved supervision, temporal graph learning under trajectory-based train/test protocols, and post hoc translational summaries that aggregate learned attribution signals to clinically legible ward- and staff-level views. To our knowledge, the particular combination developed here is not provided by the hospital AMR platforms in the literature cited above. A further distinction is the evaluation design: rather than using the platform only to quantify spread under one chosen scenario, we organise the study around a reproducible benchmark that supports comparison of observational regimes, representation choices, train-side distribution shifts, and deliberately shifted external benchmarks against fixed references, such that we can effectively identify the limits of applying digital twins to nosocomial infections.

#### Contributions

This study makes a substantive contribution to AMR control policy design in hospital settings by presenting an end-to-end hospital AMR digital twin that unifies mechanistic simulation, temporal graph-based prediction, and constrained intervention selection in a single reproducible framework. Although the platform endeavours to answer a specific IPC and clinically interesting question which we explore in depth, a modular design allows the users to engineer new learning tasks and probe the platform under their own set of settings and experimental hypotheses. Three novel technical approaches enable this framework. First, the layered integration of network-based simulation platforms with predictive graph neural networks and transformers, regulated by a policy decision module. The simulation layer captures the evolving hospital ecology of transmission, importation, antibiotic usage, surveillance, and isolation; the predictive layer learns from sequences of daily contact graphs to anticipate future AMR outcomes; and the decision module can then convert intervention-conditioned forecasts into feasible policy choices under explicit operational constraints (e.g. budgets, human resources, regulatory constraints, etc.). Second, a modular mechanism-aware and evaluation layer built on event-resolved simulator bookkeeping, such that forecasting, attribution, and policy-response endpoints can be specified from shared simulator outputs with or without altering the core simulator, graph representation, or learning architecture. This provides a scalable route to benchmarking while retaining mechanistic interpretability. Third, a validation module, benchmarked on the most dominant driver of real-time resistance emergence, and coupled to the policy decision regulator module in order to enable counterfactual evaluation of prospective intervention policies. Collectively, these contributions move hospital AMR digital twins beyond descriptive simulation and passive risk prediction toward mechanism-aware, evaluation-ready, and decision-oriented computational frameworks.

### Platform overview and experimental design

The platform generates daily directed hospital graphs with explicit ward structure, tracking admissions and discharges, infection status of new admissions, staff movement, screening logistics, and isolation policy. These graphs carry both node- and graph-level epidemiological quantities, which are then used for label construction and downstream learning. The resulting graph sequences are processed by an edge-aware GraphSAGE–Transformer architecture [40, 41] (see Methods and Supplementary Information) that combines daily graph encoding, learned graph pooling, and temporal self-attention across contiguous windows. At a high level, the framework has three linked layers. The first is a mechanistic simulation layer that produces a daily sequence of directed, weighted hospital contact graphs *G*_*d*_

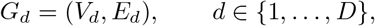

with dynamic node sets, role-aware contacts, ward structure, and surveillance-sensitive attributes. The second is a tensorisation and task-construction layer that converts each daily graph into a transformed graph-learning representation that preserves node features, edge features, trajectory identity, day index, and ward-linked metadata needed for downstream aggregation, and contains labels derived from future simulator event streams. The third is a temporal graph-learning layer that maps a rolling graph window of sequential daily contact graphs of fixed length and same trajectory origin:𝒲_*d*_ {*G*_*d−T* +1_, …, *G*_*d*_}= to future labels defined over days *d* + 1, …, *d* + *H*, given a horizon window *H*. Each daily graph is embedded by an edge-aware message-passing encoder with learned node-attention pooling, and the resulting sequence of graph embeddings is then processed by a Transformer to model temporal dependence. This separation is important. The simulator, graph representation, task definition, and learning model are coupled through a common pipeline, but not entangled into a single task-specific implementation. In practice, that makes it possible to vary forecasting targets at the task layer while leaving the underlying generation and learning machinery intact. In this line of work, we exploit this flexible framework to study one primary endpoint in depth: whether resistant emergence within the hospital over the next time horizon is predominantly driven by endogenous transmission or importation. The same modularity also supports the benchmark design used here. Canonical baseline training, representation ablations, surveillance perturbations, broader regime sweeps, an explicitly shifted train–test benchmark, and zero-shot probing of a baseline-trained model onto that shifted benchmark are all evaluated within the same pipeline. The platform should therefore be understood not simply as a simulator plus predictor, but as an integrated digital twin framework in which generation, supervision, learning, evaluation, and post hoc translational attribution are explicitly modular, scalable and connected. All benchmark analyses were conducted under two parallel representation tracks. In the *ground-truth* track, node features included the latent AMR state generated by the simulator. In the *partial-observation* track, that latent-state channel was replaced by a binary observed-positive indicator derived from surveillance, and the latent same-day incident-event channels for new resistant acquisition and new resistant infection were masked to zero; the role, antibiotic, isolation, and edge features were otherwise retained. The ground-truth track therefore measures performance under full simulator-state access, whereas the partial-observation track evaluates the same benchmark under a deliberately observation-limited representation that more closely reflects routine surveillance. Importantly, the state-related channel in the partial-observation track is not a noisy copy of latent state, but a surveillance-mediated proxy that depends on who is screened, when sampling occurs, whether results are delayed, and whether observations persist once recorded.

### Predictive experiment design

We studied the endogenous-versus-importation task in a medium-scale synthetic hospital (10 wards, 200 patients, and 300 staff). Canonical trajectories were generated over 60 simulated days, whereas the complete distribution-shift benchmark in Step 8 (Table 1) used 360-day trajectories, with 12 independent replicates per trajectory family. Staff were assigned a home ward together with multi-ward coverage, and in the main configuration each staff member was linked to two wards on average, preserving strong local ward structure while allowing staff-mediated cross-ward mixing. Prediction used rolling windows of *T* = 7 daily graphs and a forecasting horizon of *H* = 7 days. Full training parameters and experimental design can be found in Appendix A (SI) for the predictive experiment. This configuration is large enough to induce non-trivial ward structure, staff-mediated cross-ward connectivity, and temporal variation, while remaining stable enough for controlled benchmarking.

**Table 1:** Summary of the predictive experimental design. Across all experiments, analysis was conducted in parallel under the ground-truth and partial-observation representation tracks. Unless otherwise noted, the canonical frozen external test benchmark was kept fixed and restored before each downstream run.

| Step | Experiment | What was changed | What was held fixed | Purpose |
| --- | --- | --- | --- | --- |
| 1 | Canonical trajectory generation | Four independently simulated canonical trajectory families were generated: two training-side families and two test-side families, spanning endogenous-favouring and importation-favouring regimes. | Common simulator framework, common hospital scale, disjoint seed ranges for train and test. | Establish the canonical benchmark and enforce train/test separation at the simulation stage. |
| 2 | Common graph-learning conversion | All trajectories were converted into a common graph-learning representation. Parallel ground-truth and partial-observation tracks were created. | Underlying trajectories and preprocessing logic. | Ensure that later performance differences cannot be attributed to inconsistent preprocessing. |
| 3 | Frozen benchmark construction | The two canonical training-side families were pooled into one training set and the two canonical test-side families into one frozen external test set. | Same canonical cohorts and same representation track definitions. | Create a reusable held-out benchmark for all downstream comparisons. |
| 4 | Baseline external generalisation | The model was trained on the canonical training set and evaluated on the frozen canonical test set. | Canonical train/test generative logic; frozen external test set. | Measure external generalisation under matched train-test conditions. |
| 5 | Representation ablation | Training graphs were altered either by neutralising edge weights or by retaining only the role indicator and main state channel as informative node-level inputs. | Frozen canonical external test set. | Assess how strongly predictive performance depends on specific parts of the graph representation. |
| 6 | Surveillance and response perturbations | Training cohorts were regenerated under modified surveillance assumptions, specifically altered screening delay and altered routine screening frequency. For each delay or frequency level, paired endogenous-favouring and importation-favouring cohorts matching the Step 4 epidemiological regimes were generated and pooled after conversion while preserving complete simulated trajectories. | Frozen canonical external test set, canonical admission-importation and turnover regimes, screen-on-admission policy, and observation persistence. | Test whether models trained under shifted surveillance-and-response conditions transfer back to the canonical benchmark. |
| 7 | Regime robustness | Training trajectories were generated across a broader sweep of epidemiological regimes spanning different combinations of admission importation and turnover. | Frozen canonical external test set. | Evaluate whether broader training diversity improves robustness when tested on the canonical reference benchmark. |
| 8 | Complete distribution shift benchmark | Both training and test cohorts were regenerated under a deliberately shifted regime with longer temporal span, superspreader dynamics, seasonal modulation of admission importation, and paired endogenous-favouring/importation-favouring shifted trajectory families. | Shared shifted generative logic for train and test within this step, disjoint train/test seeds, and a pre-training mechanism-label balance gate. | Create a substantially more demanding shifted benchmark in which both epidemiology and temporal organisation differ from the canonical setting. |
| 9 | Baseline-to-shift transfer probe | The canonical baseline model from Step 4 was evaluated directly on the shifted external test cohort from Step 8 without retraining. | Canonical trained model; shifted test set from Step 8. | Isolate zero-shot transfer from the canonical benchmark to a markedly shifted hospital regime. |

The benchmark was designed around a fixed external reference. Four independently generated canonical trajectory cohorts were created: two for training and two for testing, spanning endogenous-favouring and importation-favouring regimes. The endogenous-favouring regime used lower admission-importation pressure and lower patient turnover, increasing the expected contribution of within-hospital emergence, whereas the importation-favouring regime used higher admission-importation pressure and higher turnover, increasing the contribution of resistant colonisation or infection introduced on admission. Admission importation is sampled over colonised-sensitive, colonised-resistant, infected-sensitive, and infected-resistant states using jointly normalised state weights, with uncolonised admissions as the residual state when the total weight is below one. Training and test cohorts were generated independently with disjoint seed ranges, to enforce train/test separation. After conversion to a common graph-learning representation, the two training-side cohorts were pooled into one canonical training set and the two test-side cohorts into one frozen external test set, which was restored before each downstream further test and used unchanged throughout the study.

Within this common framework, the experimental programme addressed three policy relevant questions: (1) Can we ascertain the mechanistic drivers of nosocomial AMR emergence in real-time? (2) How is the ability to answer the first question dependent on the type, quality and abundance of information? (3) Can we extract generalisable principles when external temporal forcing terms are present? Table 1 summarises the role of each predictive experimental step and the corresponding train–test comparison logic.

### CAUSAL policy-response experiment design

To examine whether intervention-aware temporal graph representations can support downstream decision-making, we introduced a causal policy-response assessment module integrated with the hospital simulator platform, but with explicit branching from shared pre-intervention states. The objective was not to replace the primary predictive benchmark, but to create a controlled setting in which the model could be trained to identify the best action from a small and operationally transparent intervention menu. The causal branch is trained and evaluated as a within-state action-selection problem over baseline-relative improvement in combined transmission-plus-importation resistant burden. The full description of the methodological steps is shown in Table 2 and the full parameters are given in Appendix B (SI). Let

**Table 2:** Implemented causal policy-response experiment design. The causal branch reuses the simulator framework but replaces single-path forecasting with action-conditioned counterfactual futures generated from matched simulator reruns. Policy learning and evaluation are aligned to baseline-relative improvement in future transmission-plus-importation resistant burden over a 7-day horizon.

| Step | Component | Implemented design | Held fixed | Purpose |
| --- | --- | --- | --- | --- |
| C1 | Base trajectory generation | Generated 60-day hospital trajectories under disjoint train, validation, and test seed ranges. The simulator used patient CS/CR/IS/IR day-0 seeding, low staff CS/CR day-0 seeding, shock-modulated CS/CR/IS/IR admission importation, weekly baseline screening with a 3-day reporting delay, no universal admission screening, symptomatic IS/IR admission testing, a 0.8 isolation multiplier for 3 days, and a day 14–21 super-spreader episode. | Common simulator framework and hospital setting within the causal branch. | Create temporal hospital trajectories from which intervention decision states are extracted. |
| C2 | Decision-state extraction | For each seed and each decision day $d \in \{7, 14, \dots, 49\}$, extracted a pre-intervention temporal window of length $T = 7$. | Window length $T = 7$, horizon $H = 7$, and seed-level split assignment. | Define shared observed histories for action-conditioned comparison. |
| C3 | Action-conditioned counterfactual generation | Generated each candidate future by rerunning the simulator with the same base seed and shared causal-noise seed. The intervention was inactive through day $d$ and applied from day $d + 1$. The supervised input window was fixed to the baseline pre-intervention window for all actions from the same decision state. The action set contained baseline plus four hospital-wide screening-response actions: 2-day/same-day/resistant-only, 7-day/2-day/resistant-only, 2-day/same-day/all-positive, and 7-day/2-day/all-positive. Admission screening was enabled for the four nonbaseline actions. | Base seed, causal-noise seed, decision day, and baseline pre-intervention input window. | Construct intervention-conditioned counterfactual data in which the post-decision action is the intended source of variation. |
| C4 | Outcome construction | Computed horizon-7 outcomes over days $d + 1$ through $d + 7$. The main target was baseline-relative gain, $\Delta B_7(a) = B_7(\text{baseline}) - B_7(a)$, so positive values indicate improvement. The implemented burden $B_7(a)$ is the horizon sum of `new.trans.cr.total`, `new.import.cr.total`, and `new.import.ir.total`. | Common horizon, burden definition, and baseline-relative gain construction. | Convert counterfactual futures into supervised intervention-response targets. |
| C5 | Split assignment without leakage | Assigned all action-conditioned samples derived from the same base seed to the same train, validation, or test split. Training and evaluation used manifest split labels rather than randomly splitting overlapping windows. | Seed-level split definition. | Prevent leakage across overlapping windows and related counterfactual futures. |
| C6 | Intervention-conditioned prediction | Trained the temporal graph model to score candidate actions conditional on the graph-history window, action descriptor, and operational-state summary. Training used a state-wise oracle-best-action loss plus an active within-state ranking regularizer on the continuous baseline-relative gain target. | Shared causal data construction, finite action set, and disjoint seed-level splits. | Learn the intervention-response surface needed for action ranking and decision support. |
| C7 | Policy-response evaluation | Evaluated predictions against simulator-defined oracle responses on held-out decision states. The oracle-best action was the action with the largest simulator-defined $\Delta B_7(a)$, not the action with the largest model-predicted value. Only states with complete action sets and finite oracle values were retained. | Held-out test split, common action set, and simulator-defined improvement target. | Assess whether intervention-aware representations recover action-conditioned differences in future resistant burden. |
| C8 | Constrained decision layer | Passed predicted action scores to a mixed-integer linear programming layer using baseline-relative predicted utilities. The implemented layer enforces one action per state and intervention-family cooldown rules. Action costs are loaded and reported, but cost constraints are non-binding in the current configuration unless a budget is supplied. | Learned response surface, finite action set, cooldown rules, and optional budget settings. | Translate action-conditioned forecasts into feasible intervention choices without changing the causal data construction. |

Note. The implemented horizon burden is $B_7(a) = \sum_{t=d+1}^{d+7} [\text{new.trans.cr.total}_t + \text{new.import.cr.total}_t + \text{new.import.ir.total}_t]$ . The causal-policy target is $\Delta B_7(a) = B_7(\text{baseline}) - B_7(a)$ .

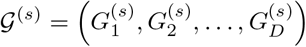

denote a base simulated trajectory generated under seed *s*, where 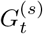 is the hospital contact graph at day *t*, and *D* is the total simulated duration. For a fixed window length *T*, forecast horizon *H*, and a set of decision days

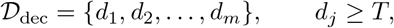

we extracted the pre-intervention temporal state

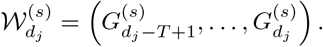

This window was held fixed and interpreted as the observed hospital history available at decision time *d*_*j*_. From the terminal state of 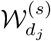, we generated a finite action set

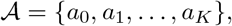

where *a*_0_ denotes the baseline policy and *a*_*k*_ for *k* ≥ 1 denotes a candidate intervention. In the present action menu, all non-baseline candidates are hospital-wide screening-response policies. They alter routine screening frequency, admission screening criteria, reporting delay, and the isolation response attached to positive tests, including whether isolation is triggered only by resistant-positive tests or by all positive tests. For each action *a* ∈ 𝒜, the simulator was rolled forward from the same pre-intervention state using matched branching logic, yielding an intervention-conditioned future trajectory

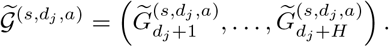

Figure 2 gives a visual representation of branching and counterfactual rollout windows. To ensure the different counter-factual trajectories are comparable, we had to control the exogenous random-number streams for turnover, admission importation, screening-test outcomes, contact formation, within-host transitions, staff-removal events, and transmission. These streams were indexed deterministically by a shared branch seed together with event type, day, and entity identity.

**Figure 1:**
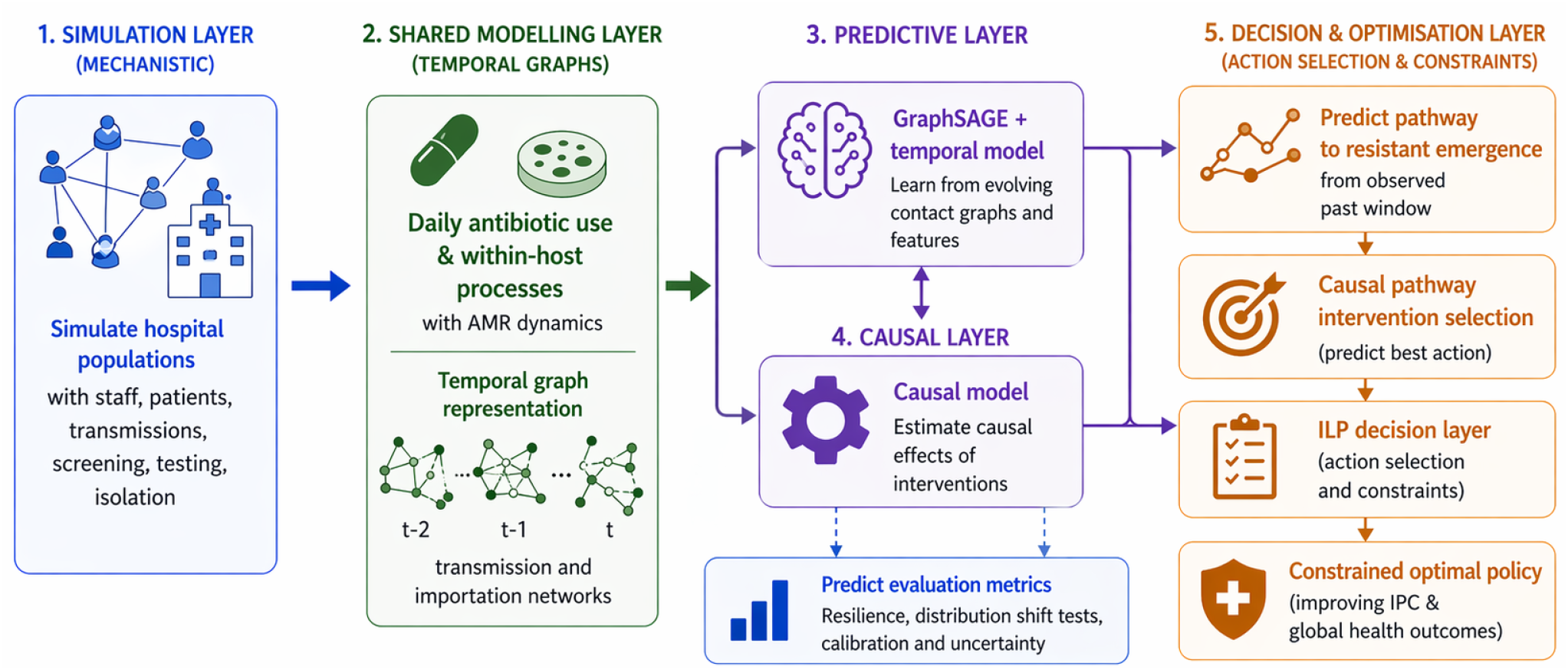
Overview of the platform. The simulation layer generates daily hospital contact graphs from a mechanistic simulator of wards, staff, patients, transmission, screening, and isolation. These temporal graphs are converted into a shared representation and processed through a common modelling backbone combining a GraphSAGE graph encoder with a Transformer temporal module. From this shared backbone, the platform branches into a predictive pathway for AMR risk forecasting and a causal pathway for intervention-conditioned prediction and constrained decision support via an integer linear programming modelling formulation (ILP). Each branch is evaluated separately according to its respective prediction or decision task.

**Figure 2:**
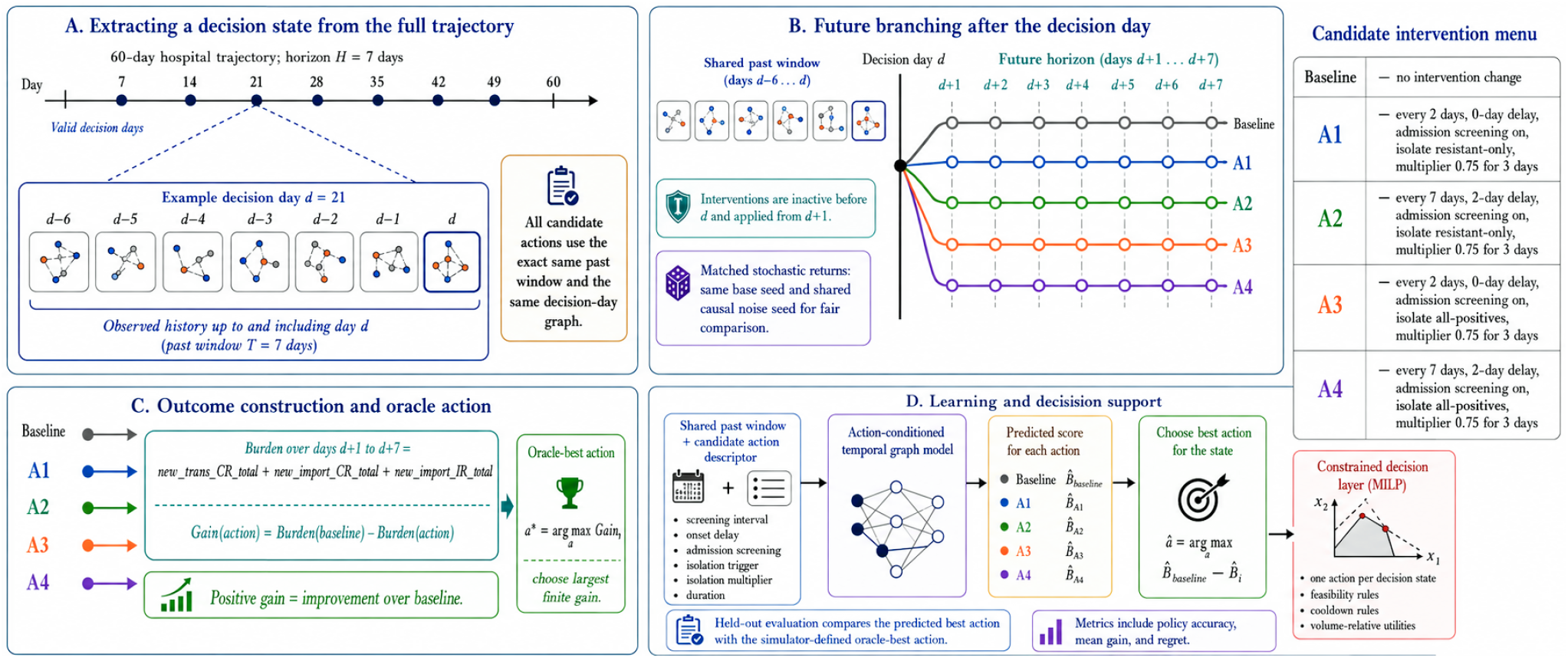
Schematic overview of past–future branching in the causal AMR policy experiment. **A: Decision-state extraction.** A 60-day simulated hospital trajectory is sampled at valid decision days, with each decision state represented by a shared temporal graph window of length *T* = 7, covering days *d* −6, …, *d*. The observed hospital history up to and including the decision day *d* is used as the common input state for all candidate actions. **B: Action-conditioned future branching**. From the same decision-day state, the baseline policy and each candidate intervention branch into matched future trajectories over the horizon *H* = 7, spanning days *d* + 1, …, *d* + 7. Interventions are inactive before the decision point and are applied only after branching, so all actions are compared from the same past state under matched stochastic reruns. **C: Outcome construction and oracle action**. For each future branch, the resistant burden over the prediction horizon is computed such that positive values indicate improvement over baseline. The simulator-defined oracle action is the candidate with the largest finite gain. **D: Learning and constrained decision support**. The action-conditioned temporal graph model receives the shared past window together with an intervention descriptor and predicts an action-specific score for each candidate intervention. The predicted best action is compared with the simulator-defined oracle action in held-out evaluation, and the resulting baseline-relative utilities can subsequently be passed to a constrained mixed-integer linear programming decision layer to select feasible intervention plans across decision states.

Thus, multiple action-labelled samples share the same observed past 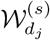but differ in their post-decision futures. For a single seed *s* and decision day *d*_*j*_, the dataset contains one sample for each action *a* ∈ 𝒜,

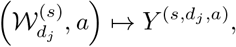

where 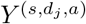 is a horizon-*H* outcome derived from the corresponding branched trajectory. In the present configuration, *T* = 7, *H* = 7, the action set is shown in Table 3, and branching is performed weekly until no more windows can be generated using all simulated days, with the intervention activated from day *d*_*j*_ + 1. The main oracle used for policy learning and evaluation is the simulator-derived quantity defined by

**Table 3:** Candidate intervention menu used for counterfactual policy evaluation. All candidate interventions are applied globally at hospital level. The baseline action is included as the no-change comparator.

| ID | Action | Screening schedule | Admission screening | Isolation response after positive test |
| --- | --- | --- | --- | --- |
| A0 | Baseline comparator | Baseline simulator policy | Baseline simulator policy | Baseline simulator policy |
| A1 | Screen every 2 days; resistant-only isolation trigger | Every 2 days; same-day result | Yes | Resistant-positive tests only; contact multiplier 0.75 for 3 days |
| A2 | Screen every 7 days; resistant-only isolation trigger | Every 7 days; 2-day reporting delay | Yes | Resistant-positive tests only; contact multiplier 0.75 for 3 days |
| A3 | Screen every 2 days; all-positive isolation trigger | Every 2 days; same-day result | Yes | All positive tests; contact multiplier 0.75 for 3 days |
| A4 | Screen every 7 days; all-positive isolation trigger | Every 7 days; 2-day reporting delay | Yes | All positive tests; contact multiplier 0.75 for 3 days |

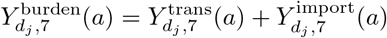

and

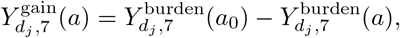

where *a*_0_ denotes the baseline policy. Larger values are preferred, since they correspond to greater improvement relative to the baseline policy outcomes for the same decision state. When actions tie exactly on this oracle quantity, the winning label is resolved by a fixed action ordering recorded at dataset construction and kept unchanged during training and evaluation.

To prevent leakage across overlapping temporal windows from the same underlying simulator realization, dataset splits were defined at trajectory generation time rather than by random row-level partitioning. Separate seed ranges were assigned to training, validation, and test partitions, and all action-conditioned decision samples derived from a given seed inherited the same split assignment. Consequently, the train/validation/test sets correspond to disjoint sets of simulated hospital trajectories with a fixed temporal windows.

The implementation of this additional feature ensures the model can be trained on repeated counterfactual branches from shared decision states. This allows for the direct comparison of the expected evolution of hospital states following the implementation of different policies, thus facilitating nosocomial AMR policy optimisation. These predicted action-conditioned utilities are then passed to the downstream constrained mixed-integer decision layer, so the optimizer acts on the learned intervention-response surface rather than altering how the counterfactual branches are generated. Full implementation details and parameters are given in Appendix C (SI).

## Results

Quantitative summaries for the predictive benchmark are shown in Figure 3, Figure 4, Figure 5, and Figure 6, with the corresponding metrics reported in Table 4 and Table 5. Because the same frozen external test cohort was restored before each downstream canonical run, all post-baseline canonical comparisons are made against the same held-out reference. The expanded results for the causal experiment are given separately in Figure 7 which also contain the performance metrics for both tracks.

**Table 4:**
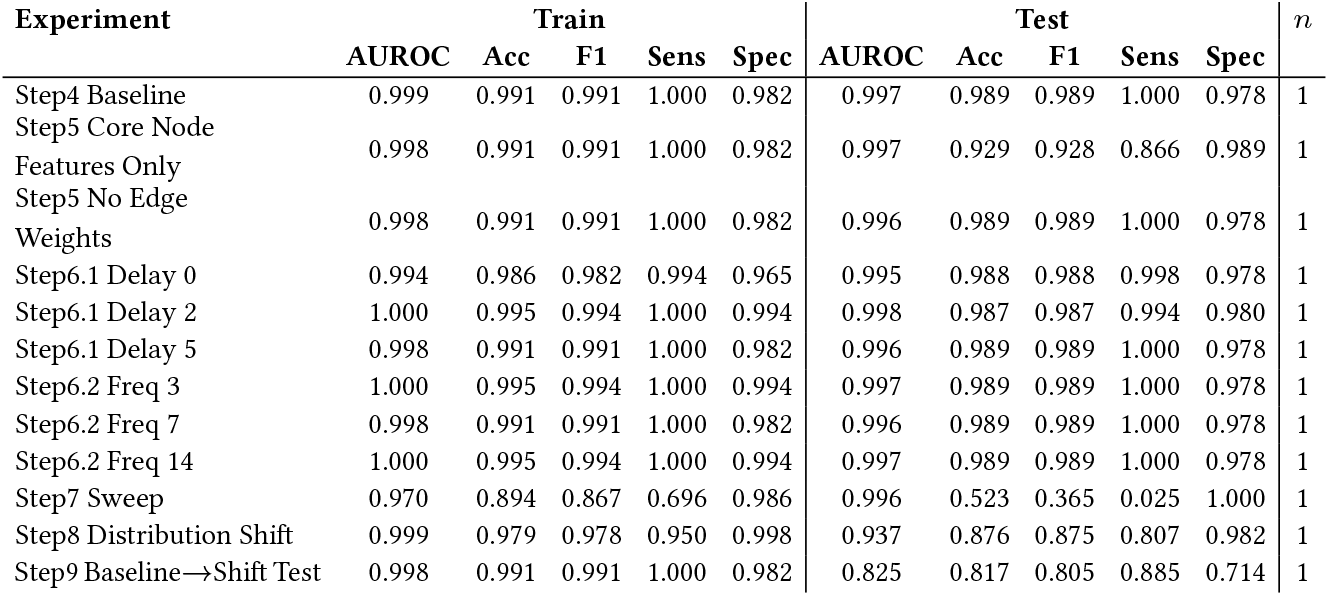
Compact quantitative summary for ground truth track at *T* = 7 and *H* = 7.

**Table 5:**
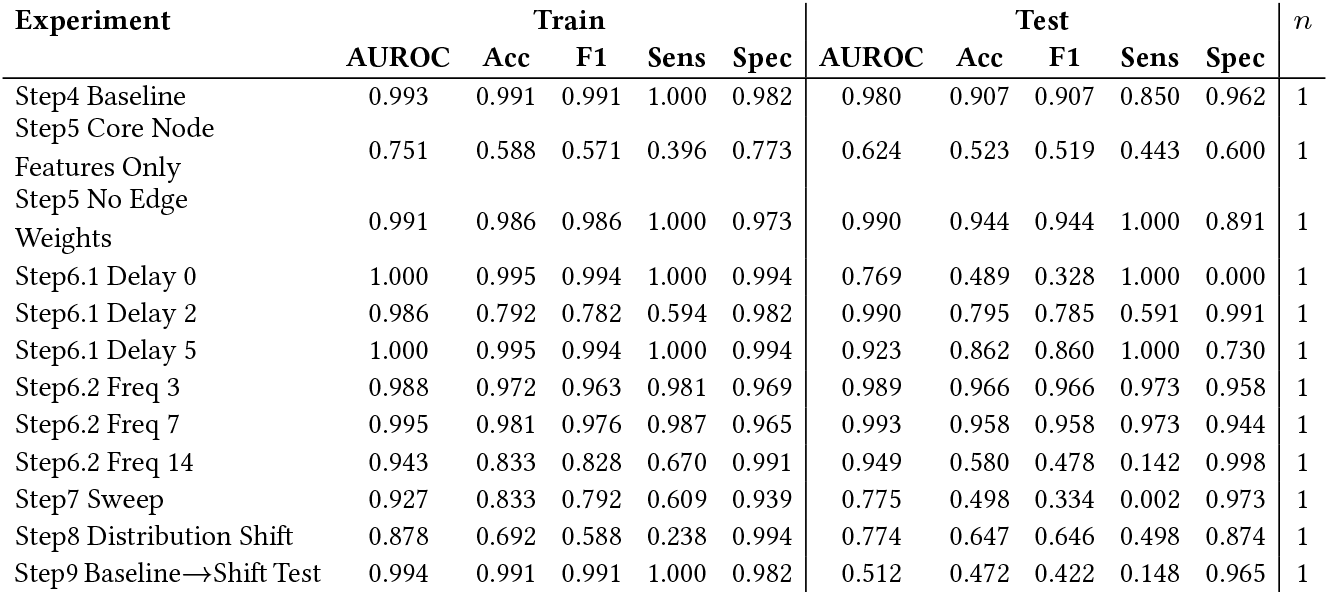
Compact quantitative summary for partial observation track at *T* = 7 and *H* = 7.

**Figure 3:**
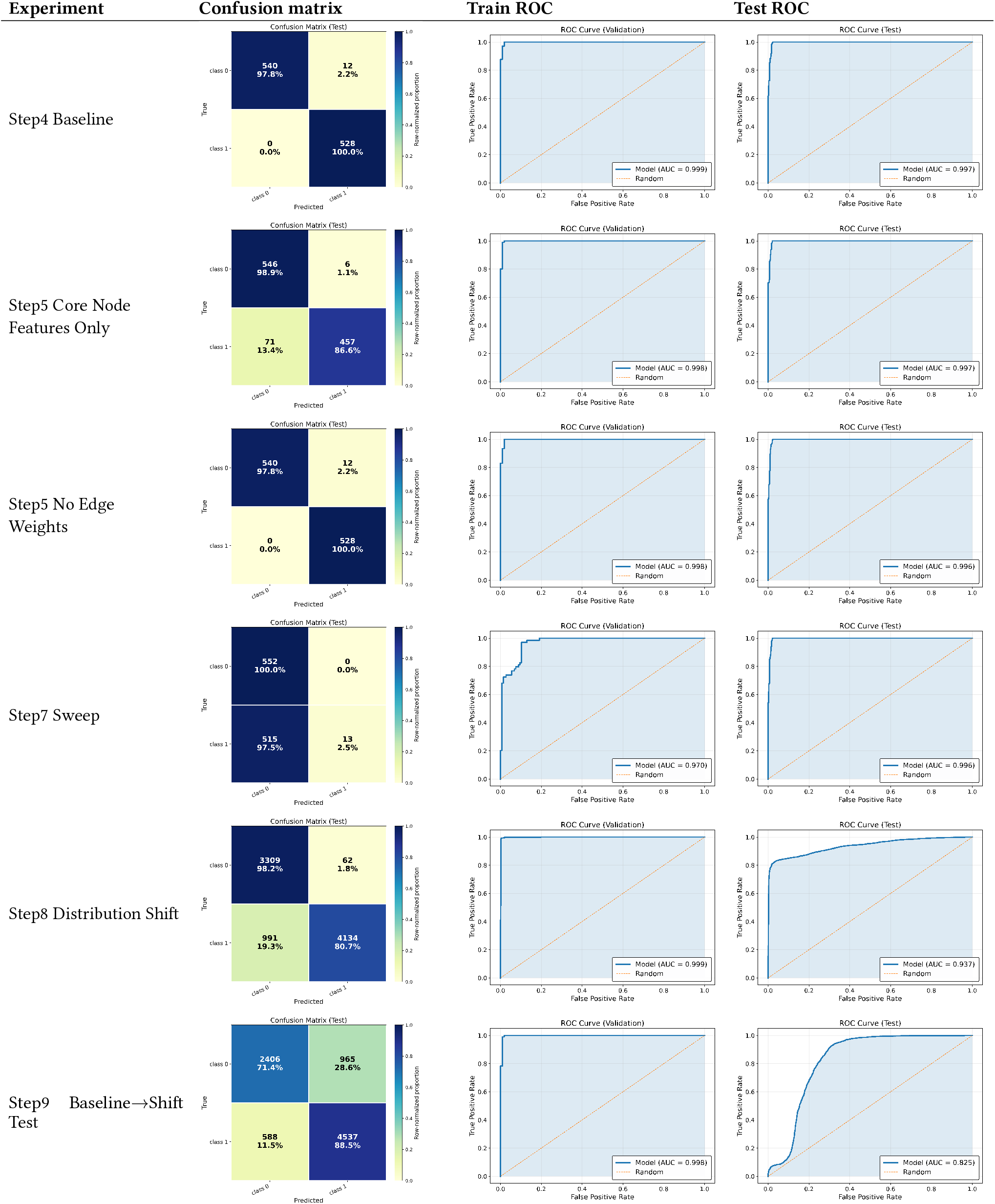
Main comparison grid for ground truth track at *T* = 7 and *H* = 7. Step 6 delay/frequency ablations are shown separately in Figures5, 6.

**Figure 4:**
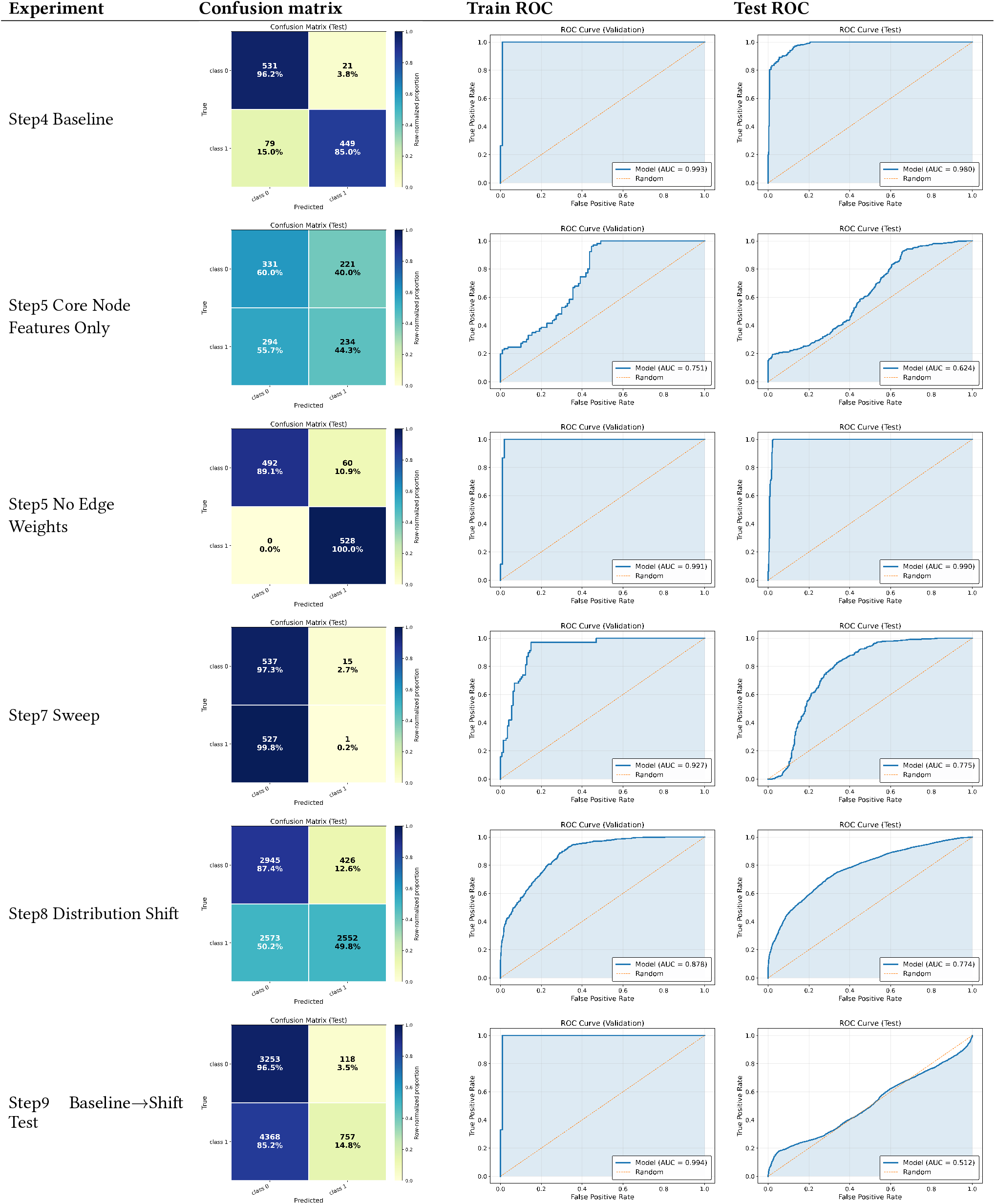
Main comparison grid for partial observation track at *T* = 7 and *H* = 7. Step 6 delay/frequency ablations are shown separately in Figures5, 6.

**Figure 5:**
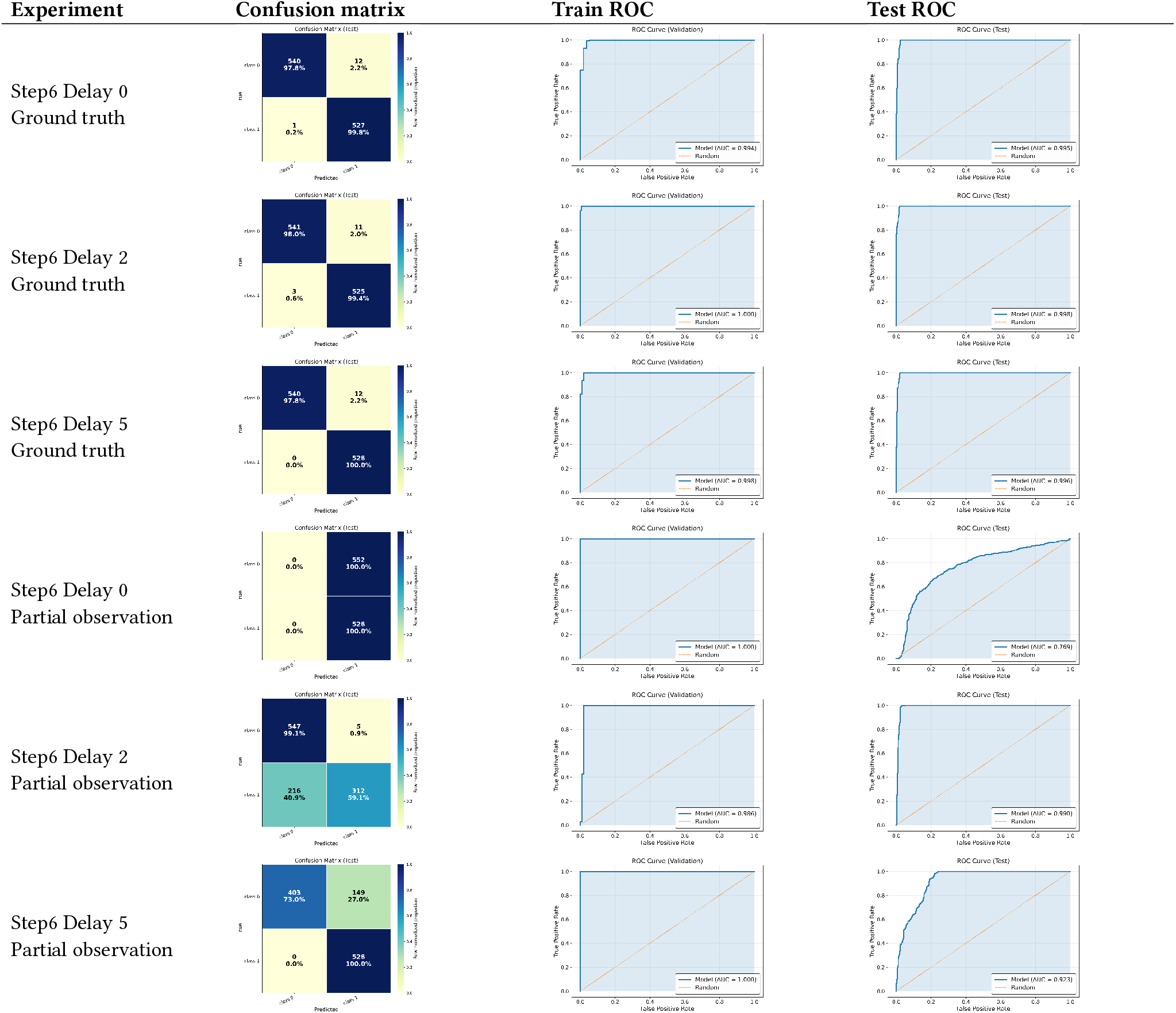
Cross-track comparison of Step 6 delay ablations at *T* = 7 and *H* = 7. Rows are grouped by intervention level and state-observation regime.

**Figure 6:**
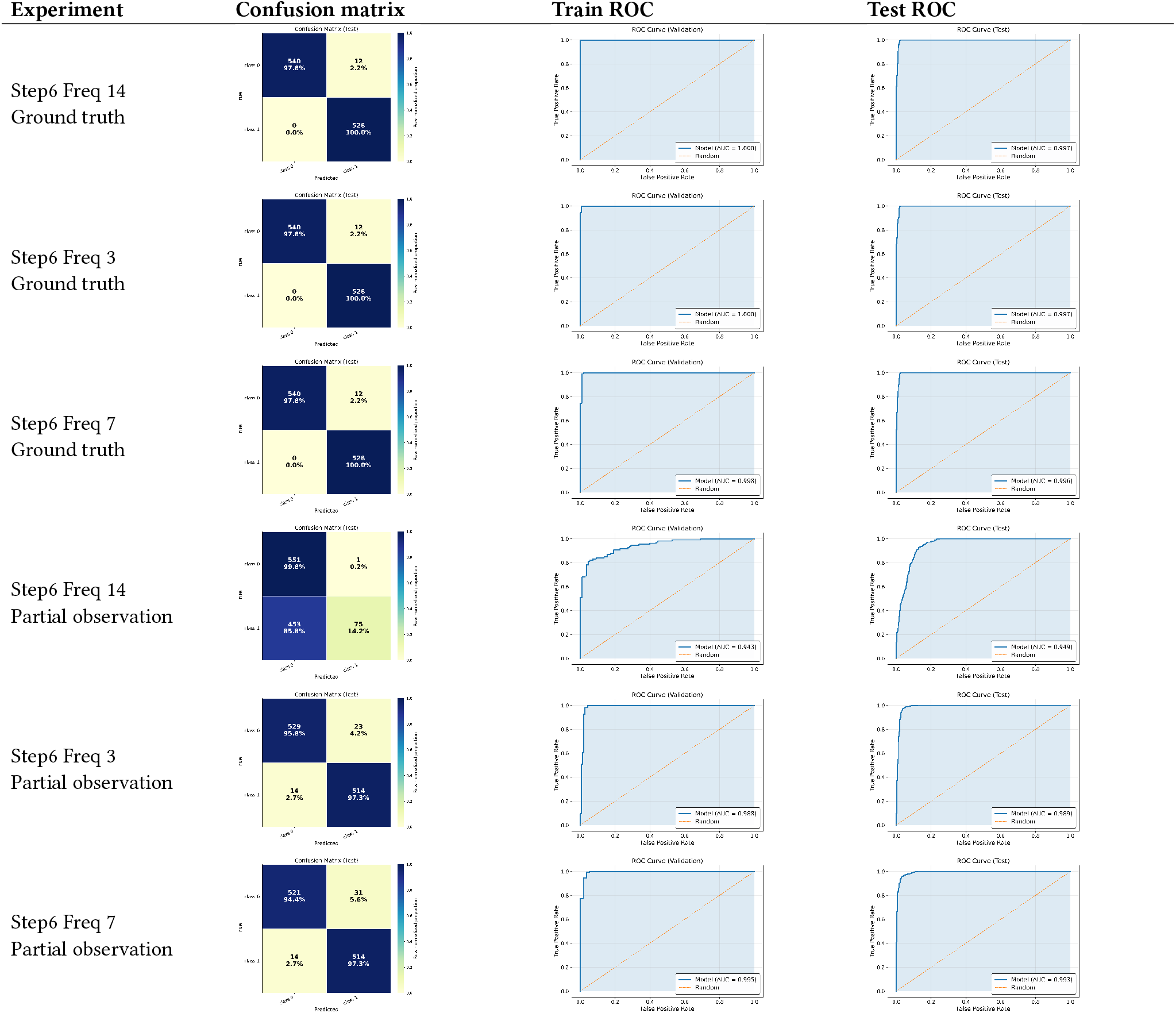
Cross-track comparison of Step 6 screening frequency ablations at *T* = 7 and *H* = 7. Rows are grouped by intervention level and state-observation regime.

**Figure 7:**
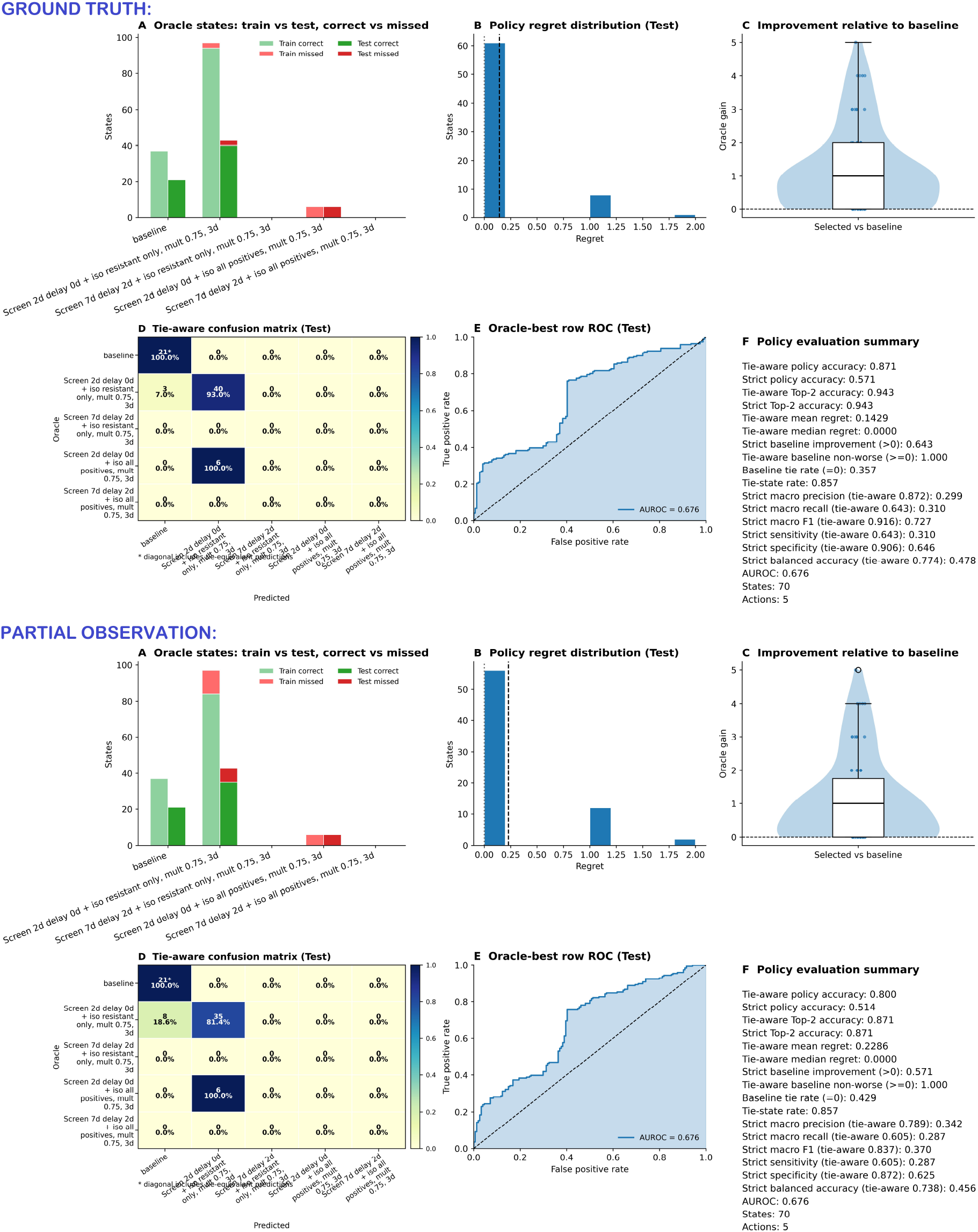
Policy-selection performance under ground-truth and partial-observation state representations. The top dashboard summarises the ground-truth track and the lower the partial-observation track. Panel A compares the oracle-best action distribution across training and test states, separating correctly selected and missed actions. Panel B shows the test-set regret distribution, quantifying the loss incurred when the selected intervention differs from an oracle-optimal action. Panel C summarises the predicted improvement relative to the baseline comparator across decision states. Panel D presents the tie-aware confusion matrix, where predictions matching any oracle-tied optimal action are credited as correct. Panel E shows the one-vs-rest receiver operating characteristic curve for oracle-best action discrimination. Panel F reports the corresponding policy-evaluation summary metrics. Together, the panels show that the policy layer retains broadly consistent intervention-selection behaviour across fully observed and partially observed hospital states, while exposing the expected degradation under incomplete surveillance.

### Baseline external generalisation

The baseline provides the clearest reference point. Four canonical trajectory families were generated using disjoint seeds: two under a low-importation, low-turnover regime favouring endogenous amplification, and two under a high-importation, higher-turnover regime favouring externally driven burden. After conversion and pooling, the model was trained on the canonical training set and evaluated on the frozen external test set. The primary task was endogenous-versus-importation majority over a seven-day horizon. Writing

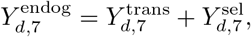

and comparing it against 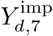, the binary label indicates whether endogenous resistant emergence, defined as transmission plus within-host selection, exceeds importation over days *d* + 1, …, *d* + 7. The strongest result was obtained under the canonical ground-truth benchmark. In that setting, the model generalised almost perfectly to the frozen external test set, with test AUROC = 0.997, accuracy = 0.989, F1 = 0.989, sensitivity = 1.000, and specificity = 0.978 (Table 4). In Figure 3, this appears as both an almost ideal ROC curve and an almost error-free confusion matrix. The partial-observation baseline was also strong: test AUROC reached 0.980, with accuracy = 0.907, F1 = 0.907, sensitivity = 0.850, and specificity = 0.962 (Table 5). The main message is therefore not that observation limitation breaks the task, but that replacing the latent-state channel with a surveillance-derived observed-positive proxy reduces recall and thresholded accuracy while preserving a strong ranking signal.

### Representation ablation

The ablation experiments examined how strongly performance depended on particular parts of the graph representation. Both ablation datasets were derived from the canonical training cohort, while the frozen external test cohort was left unchanged. The first ablation removed variation in edge weights while preserving graph connectivity and edge type. The second reduced the node representation to a smaller informative subset by suppressing antibiotic class, isolation state, and same-day event indicators while retaining the role indicator and the main state channel.

In the ground-truth track, both ablations retained substantial predictive utility on the external benchmark, but their effects were not identical. The core-node-features-only ablation achieved test AUROC = 0.997, accuracy = 0.929, F1 = 0.928, sensitivity = 0.866, and specificity = 0.989, while the no-edge-weights ablation achieved test AUROC = 0.996, accuracy = 0.989, F1 = 0.989, sensitivity = 1.000, and specificity = 0.978. Relative to baseline, removing edge-weight variation was nearly cost-free at the operating threshold, whereas the compressed node representation reduced sensitivity and accuracy.

The partial-observation track showed a sharper separation between the two ablations. Core-node-features-only weakened materially, with test AUROC = 0.624, accuracy = 0.523, F1 = 0.519, sensitivity = 0.443, and specificity = 0.600. The no-edge-weights ablation remained strong, with test AUROC = 0.990, accuracy = 0.944, F1 = 0.944, sensitivity = 1.000, and specificity = 0.891. Thus, in the current benchmark, the edge-weight channel is not indispensable, but the partial-observation model is much more dependent on the richer node-feature set than the ground-truth model.

### Observation and surveillance perturbations

The surveillance perturbation analyses were designed as transfer experiments under altered training-side screening assumptions. New training cohorts were generated under modified screening conditions, but evaluation still took place on the same frozen canonical benchmark. For each Step 6 delay or screening-frequency condition, paired endogenous-favouring and importation-favouring regimes were generated before pooling, so that surveillance perturbations are not confounded with a severe mechanism-label imbalance in the training data. Because the simulator allows positive surveillance signals to trigger isolation and thereby alter subsequent transmission, these experiments should still be read as joint perturbations of surveillance and the response pathways that depend on surveillance, not as pure observability ablations.

The first set of experiments varied screening-result delay. In the ground-truth track, transfer back to the canonical benchmark remained very strong. Test AUROC values were 0.995, 0.998, and 0.996 for delays of 0, 2, and 5 days, respectively, with corresponding accuracies of 0.988, 0.987, and 0.989 (Table 4). Sensitivity remained close to one (0.998, 0.994, 1.000) and specificity remained high (0.978, 0.980, 0.978). Thus, under full simulator-state access, the delay perturbations did not materially weaken either ranking or thresholded classification.

The partial-observation delay results were more heterogeneous. Test AUROC values were 0.769, 0.990, and 0.923 for delays of 0, 2, and 5 days, with accuracies of 0.489, 0.795, and 0.862, respectively. Delay 0 collapsed to an all-positive operating point, with sensitivity = 1.000 and specificity = 0.000. Delay 2 achieved excellent ranking and specificity (AUROC = 0.990, specificity = 0.991) but lower sensitivity (0.591), whereas delay 5 gave a stronger thresholded balance with sensitivity = 1.000 and specificity = 0.730.

The second set of experiments varied routine screening frequency. In the ground-truth track, all three frequency perturbations remained near baseline: test AUROC values were 0.997, 0.996, and 0.997 for frequencies of 3, 7, and 14 days, respectively, with accuracies of 0.989 in all three cases. Sensitivity was 1.000 throughout and specificity was 0.978 in all three cases.

In the partial-observation track, the 3- and 7-day frequency conditions were also strong, with test AUROC/accuracy of 0.989*/*0.966 and 0.993*/*0.958, respectively. The 14-day condition retained high AUROC (0.949) but produced a poor thresholded operating point, with accuracy = 0.580, F1 = 0.478, sensitivity = 0.142, and specificity = 0.998. Overall, the Step 6 experiments indicate that surveillance-linked training changes are not inherently damaging in the ground-truth track, but they can induce threshold instability under partial observation.

### Regime robustness

The regime-sweep experiment broadened the training distribution beyond the two canonical extremes by adding simulations across a wider range of importation and turnover settings. The aim was to test whether greater training diversity improves transfer back to the canonical benchmark. The results showed a clear ranking-versus-threshold separation. In the ground-truth track, the sweep model achieved test AUROC = 0.996, indicating that the score ordering transferred very well. However, the fixed operating point was poor, with accuracy = 0.523, F1 = 0.365, sensitivity = 0.025, and specificity = 1.000. In the partial-observation track, both ranking and thresholded classification were weaker: test AUROC = 0.775, accuracy = 0.498, F1 = 0.334, sensitivity = 0.002, and specificity = 0.973. Thus, broader training diversity did not stabilise the canonical decision threshold. In the ground-truth track it preserved excellent ranking information but yielded a sub-optimal operating point, whereas under partial observation it was substantially weaker overall.

### Complete distribution-shift benchmark

Step 8 moves both training and testing into a deliberately shifted generative regime. In contrast to the canonical benchmark, where train and test follow the same core simulation logic and differ only through independent stochastic realisations, the Step 8 benchmark introduces a broader temporal and mechanistic shift through longer trajectories, admission-importation seasonality, and superspreader-driven contact amplification. The shifted benchmark is constructed from paired endogenous-favouring and importation-favouring shifted trajectory families for both training and testing. Performance remained strong in the ground-truth track when both learning and evaluation took place within the shifted world. Step 8 achieved test AUROC = 0.937, accuracy = 0.876, F1 = 0.875, sensitivity = 0.807, and specificity = 0.982. In the partial-observation track, performance was more modest, with test AUROC = 0.774, accuracy = 0.647, F1 = 0.646, sensitivity = 0.498, and specificity = 0.874. These results show that the platform can still learn a useful endogenous-versus-importation decision boundary inside a materially shifted regime, but that the shifted partial-observation setting is substantially harder than the shifted ground-truth setting.

### Baseline-to-shift transfer probe

Step 9 isolates a stricter transfer question. Instead of retraining on the shifted Step 8 training distribution, the model learned from the canonical Step 4 baseline was evaluated directly on the shifted Step 8 external test cohort. This creates a zero-shot transfer setting in which the model must operate under changed temporal structure, changed importation dynamics, and additional mechanisms such as seasonality and superspreader effects without any exposure to those regimes during training. Here the transfer pattern differed sharply by representation track. In the ground-truth track, zero-shot transfer retained usable performance, with test AUROC = 0.825, accuracy = 0.817, F1 = 0.805, sensitivity = 0.885, and specificity = 0.714. In the partial-observation track, test AUROC fell to 0.512, with accuracy = 0.472, F1 = 0.422, sensitivity = 0.148, and specificity = 0.965. Compared with Step 8, these results indicate that the main difficulty is not learning under the shifted world per se in the ground-truth setting, but transporting a partial-observation decision rule learned under the canonical world into that shifted world without retraining.

### Causal experiment

The causal branch now evaluates a 7-day intervention-selection task. In each track, policy evaluation retained 70 held-out decision states with a complete five-action set consisting of the baseline comparator and four hospital-wide screening-response interventions. Because the simulator can assign the same best gain to more than one action, we report both tie-aware metrics, which count any oracle-best action as correct, and strict metrics, which compare against the fixed tie-broken oracle label stored in the manifest. Table 6 contains the detailed summary metrics per track that are also given in panels *F* of the respective dashboards Figure 7.

**Table 6:**
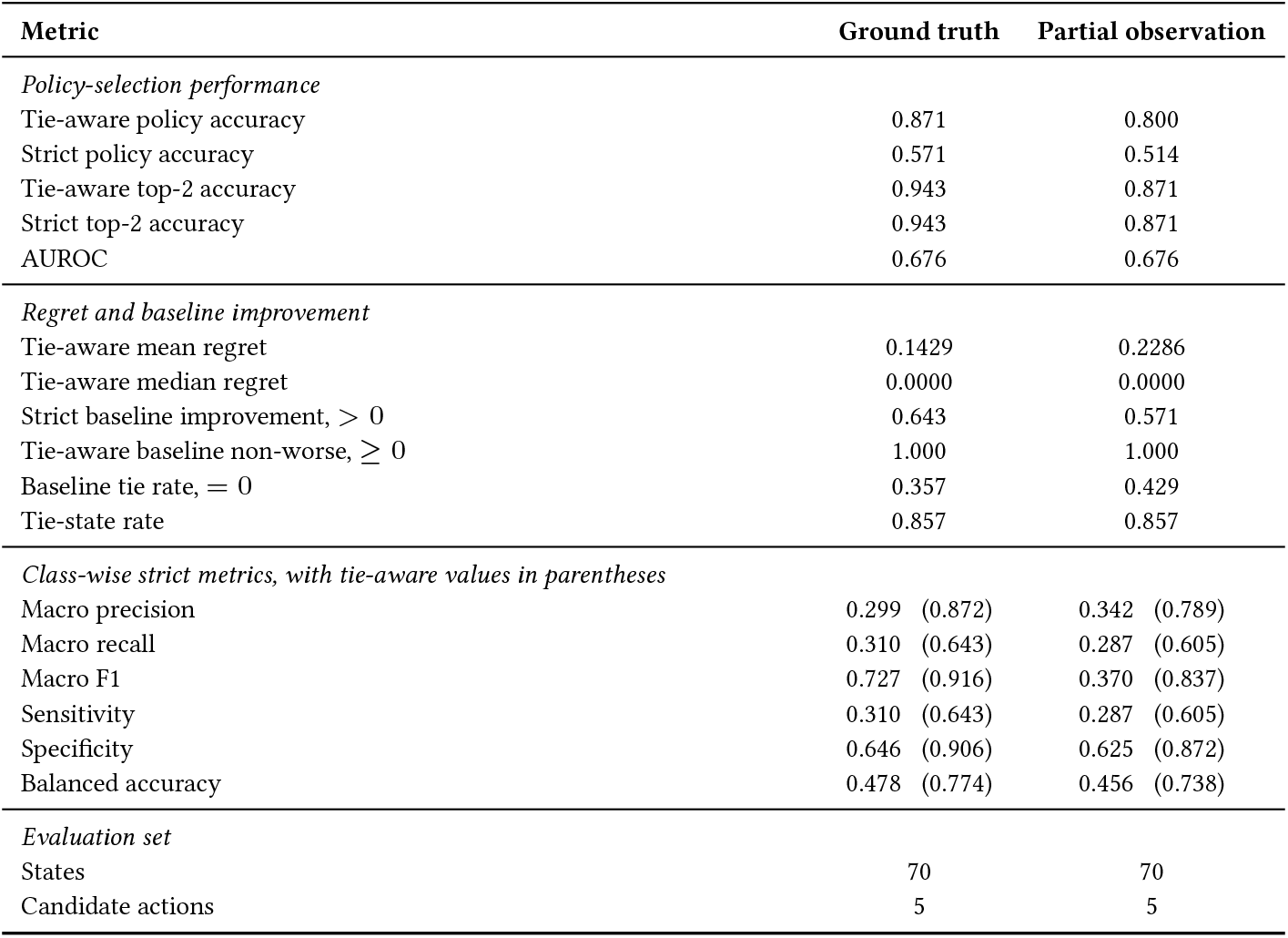
Policy selector evaluation metrics for the causal experiment. Metrics are reported separately for the ground-truth and partial-observation tracks.

| Metric | Ground truth | Partial observation |
| --- | --- | --- |
| *Policy-selection performance* |  |  |
| Tie-aware policy accuracy | 0.871 | 0.800 |
| Strict policy accuracy | 0.571 | 0.514 |
| Tie-aware top-2 accuracy | 0.943 | 0.871 |
| Strict top-2 accuracy | 0.943 | 0.871 |
| AUROC | 0.676 | 0.676 |
| *Regret and baseline improvement* |  |  |
| Tie-aware mean regret | 0.1429 | 0.2286 |
| Tie-aware median regret | 0.0000 | 0.0000 |
| Strict baseline improvement, $> 0$ | 0.643 | 0.571 |
| Tie-aware baseline non-worse, $\geq 0$ | 1.000 | 1.000 |
| Baseline tie rate, $= 0$ | 0.357 | 0.429 |
| Tie-state rate | 0.857 | 0.857 |
| *Class-wise strict metrics, with tie-aware values in parentheses* |  |  |
| Macro precision | 0.299 (0.872) | 0.342 (0.789) |
| Macro recall | 0.310 (0.643) | 0.287 (0.605) |
| Macro F1 | 0.727 (0.916) | 0.370 (0.837) |
| Sensitivity | 0.310 (0.643) | 0.287 (0.605) |
| Specificity | 0.646 (0.906) | 0.625 (0.872) |
| Balanced accuracy | 0.478 (0.774) | 0.456 (0.738) |
| *Evaluation set* |  |  |
| States | 70 | 70 |
| Candidate actions | 5 | 5 |

In the ground-truth track, tie-aware policy accuracy was 0.871 and strict policy accuracy was 0.571; both tie-aware and strict top-2 accuracies were 0.943. The tie-aware mean regret was 0.1429 and the median regret was 0, indicating that most held-out states incurred no loss relative to an oracle-best action but that a small positive regret tail remained. The chosen action strictly improved on baseline in 64.3% of held-out states, and the tie-aware non-worse rate relative to baseline was The oracle-best-row AUROC was 0.676. The partial-observation track was weaker at the same operating point: tie-aware policy accuracy 0.800, strict policy accuracy 0.514, tie-aware and strict top-2 accuracies 0.871, mean regret 0.2286, strict baseline-improvement rate 0.571, and tie-aware non-worse rate 1.000. Its oracle-best-row AUROC was also 0.676, indicating modest score-level discrimination in both tracks rather than a ground-truth-specific separation advantage.

The dashboards also clarify where the remaining policy errors occur. In the ground-truth track, the baseline comparator was oracle-best in 21 held-out states and all 21 were recovered under tie-aware accounting. The 2-day same-day screening policy with resistant-only isolation was oracle-best in 43 states, of which 40 were recovered exactly and 3 were assigned to baseline. The 2-day same-day screening policy with all-positive isolation was oracle-best in 6 states, and all 6 were assigned to the 2-day resistant-only screening action. In the partial-observation track, the same oracle-state distribution appeared, but the 2-day resistant-only action was recovered in 35*/*43 states and 8 were assigned to baseline; the 6 all-positive oracle-best states were again assigned to the 2-day resistant-only action. The 7-day delayed screening variants were not oracle-best in the retained test states. Consistent with this pattern, the regret histograms are concentrated at zero but include short positive tails, and the improvement violin/box plots show that selected actions were non-worse than baseline under tie-aware accounting while strict positive baseline-relative gains were achieved in a smaller fraction of states.

The current causal results therefore support a bounded but meaningful claim. The intervention-conditioned temporal graph model is not merely distinguishing baseline from non-baseline actions; it is ranking a five-action menu with moderate-to-high tie-aware held-out accuracy, high top-2 recovery, and universal tie-aware non-worsening relative to baseline. At the same time, score-level separation is modest, strict accuracy is lower than tie-aware accuracy, the retained test set is dominated by the baseline comparator and the 2-day resistant-only screening action, and the 7-day delayed screening variants are never oracle-best. The present branch should therefore be read as promising within-regime policy ranking over the sampled decision states rather than as a fully calibrated intervention engine across a balanced policy space.

### Summary of results and limitations

First, the canonical ground-truth baseline is genuinely excellent and shows that the temporal graph model can recover a clinically meaningful mechanism-aware endpoint under a clean external benchmark. Second, the partial-observation baseline remains strong, but it is not near-ceiling: ranking is high, while thresholded classification loses sensitivity and overall accuracy relative to the ground-truth track. Third, among the ablations, removing edge weights is far less harmful than compressing node information, and under partial observation the compressed core-node-feature representation is the clearest representation-level failure mode.

Beyond these settings, the picture depends strongly on whether ranking or fixed-threshold performance is being assessed. Step 6 perturbations are highly robust in the ground-truth track, and several partial-observation settings also remain strong. However, partial-observation delay 0 and frequency 14 demonstrate that altered surveillance-linked training dynamics can destabilise the operating threshold even when AUROC remains informative. Step 7 makes this distinction sharper: the ground-truth sweep preserves excellent AUROC but collapses at the threshold, whereas the partial-observation sweep is weak on both ranking and thresholded classification. The combination of Table 4, Table 5, Figure 3, Figure 4, Figure 5, and Figure 6 is therefore essential for interpretation, because the tables quantify the operating-point failures while the figure panels show whether any broader score ordering has survived.

The broader shift results require a specific interpretation. Step 8 does not indicate general failure under harder epidemiological conditions in the ground-truth track; when training and testing are matched within the deliberately shifted regime, the model still learns a strong endogenous-versus-importation boundary. Under partial observation, however, the shifted benchmark is only moderate. The sharper weakness is Step 9 in the partial-observation track, where the canonical baseline model is transported directly to the shifted test set without retraining and AUROC falls close to random, but the interpretation should be cautious as the broader regime shift test set from Step 8 contains previously unseen strongly affecting mechanisms such as seasonality and superspreading. The main robustness limitation is therefore threshold calibration and zero-shot portability across regimes, especially when the representation is surveillance-mediated. The Step 6 experiments should moreover be interpreted carefully: because changing surveillance can also alter isolation and hence downstream spread in the simulator, they probe transfer under joint surveillance-and-response perturbations rather than under pure missingness. The natural follow-up questions are therefore calibration, threshold selection, repeated-seed robustness assessment, and cleaner decoupling of observation from intervention in future sensitivity analyses, rather than simply larger model capacity. Step 6 cohorts are constructed by paired endogenous-favouring and importation-favouring training regimes, and Step 8 additionally applies an explicit mechanism-label balance gate before shifted training. Threshold calibration remains a separate issue because a paired or gated training cohort does not guarantee transportability of a fixed decision threshold.

The causal branch adds a distinct but bounded result. Under the current 7-day intervention-ranking task, the ground-truth track achieved tie-aware policy accuracy 0.871 and tie-aware top-2 accuracy 0.943 on 70 evaluable held-out states, with strict baseline improvement in 64.3% of states and tie-aware non-worsening in 100.0%. The partial-observation track achieved tie-aware policy accuracy 0.800, tie-aware top-2 accuracy 0.871, strict baseline improvement in 57.1% of states, and the same tie-aware non-worsening rate of 100.0%. Oracle-best-row AUROC was equal in both tracks. Most residual error was concentrated around the baseline and closely related 2-day screening-response variants, while the 7-day delayed screening variants were not oracle-best in the retained test states. This suggests that the learned representation supports useful within-state action ranking under the sampled intervention menu, but also that score calibration and policy-space balance remain important challenges.

## Methods

This section describes the methodological components of the digital twin in the order in which they operate. We begin with the mechanistic simulator that generates daily hospital contact graphs and associated epidemiological event streams. We then describe graph tensorisation and the construction of mechanism-aware forecasting labels from future simulator outcomes. Finally, we present the temporal graph-learning model and the intervention-conditioned policy layer used for causal decision support. Full implementation-level parameter definitions, feature encodings, and extended mathematical details are provided in the Supplementary Information.

### Daily hospital graph simulator

The simulator generates a sequence of daily directed weighted graphs

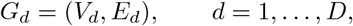

where *V*_*d*_ = *P*_*d*_ ∪ *S* consists of patients present on day *d* and staff. Each patient is assigned to one ward, while each staff member has a home ward and a multi-ward coverage set. This produces a role-aware and ward-aware graph in which within-ward patient interactions, staff–patient contacts, and staff–staff contacts are all represented. Each directed edge (*u, v*) ∈ *E*_*d*_ carries a relation type

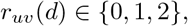

corresponding to patient–patient, staff–patient, or staff–staff contact, and a continuous contact weight *ω*_*uv*_(*d*). Thus, the daily graph is a marked directed graph

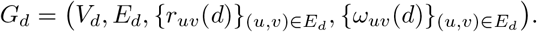

The patient census evolves through ward-specific admissions and discharges. Patients who remain in hospital retain stable identifiers over time, while newly admitted patients receive fresh identifiers and are initialised with latent AMR state, antibiotic status, isolation status, observation status, and event indicators. Admission can introduce uncolonised patients, patients colonised with sensitive or resistant strains, or patients infected with sensitive or resistant strains. The admission importation parameters are interpreted as CS/CR/IS/IR state weights and are sampled jointly; when their total exceeds one, they are normalised to preserve the relative composition, and otherwise the remaining mass corresponds to uncolonised admissions. These weights may be constant, seasonally modulated, or shock-modulated, allowing the simulator to represent both stable endemic pressure and transient importation surges. Each individual occupies a latent AMR state

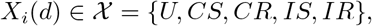

where *U* denotes uncolonised, *CS* and *CR* denote sensitive and resistant colonisation, and *IS* and *IR* denote sensitive and resistant infection. Individuals also carry an antibiotic exposure state

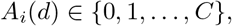

where *A*_*i*_(*d*) = 0 denotes no current antibiotic exposure and *A*_*i*_(*d*) *>* 0 denotes treatment under one of *C* antibiotic classes. Antibiotic initiation is more likely among infected individuals than among non-infected individuals, while discontinuation is stochastic. Latent-state transitions include antibiotic-associated selection from sensitive to resistant states, progression from colonisation to infection, and stochastic clearance. Consequently, resistant emergence can arise through direct resistant importation, contact-mediated resistant transmission, or within-host treatment-associated selection. Transmission is evaluated over directed incoming contacts. For an uncolonised target *i*, let

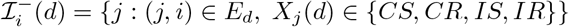

be its infectious incoming source set. For source *j*, the state-specific infectivity multiplier (*γ*) of the source individual depends on clinical state rather than resistance phenotype alone (*γ*_*colonized*_ = 0.5, *γ*_*infected*_ = 1.0). Thus colonised carriers are less infectious than infected carriers within both sensitive and resistant phenotypes. The resistant-type multiplier is retained as a separate term, but in the present experiments it is neutral, so resistant carriage or infection is not made globally more transmissible than sensitive carriage or infection. In the implementation, isolation and cross-ward hand hygiene attenuate the realised edge-level exposure probability after the baseline exponential dose-response calculation. If *q*_*j*_(*d*), *q*_*i*_(*d*) ∈ {0, 1} indicate whether source and target are isolated, respectively, then the isolation attenuation factor is

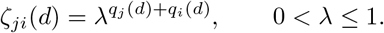

so that each isolated endpoint independently reduces the realised exposure probability by a multiplicative factor *λ*. Cross-ward staff–patient contacts can also be attenuated by a leaky hand-hygiene multiplier when enabled in the present experiments. Let

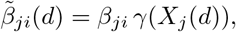

where *β*_*ji*_ depends on contact type. The unattenuated edge-level exposure probability is

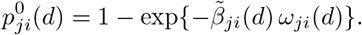

The probability used for the Bernoulli transmission draw on that directed edge is then

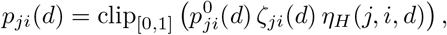

where *η*_*H*_ (*j, i, d*) is the cross-ward hand-hygiene factor and clip_[0,1]_ truncates probabilities to the unit interval. Assuming conditional independence of edge-level exposure events given the daily graph and agent states, the probability of at least one contact-mediated acquisition opportunity is

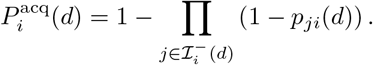

The simulator records successful transmission events explicitly. Each successful infectious contact stores the source, target, source state, transmitted bacterial phenotype, target context, contact type, edge weight, isolation status, and antibiotic context. Newly colonised agents inherit the bacterial phenotype from the selected successful transmitting source. If more than one successful exposure occurs on the same day, the final infecting event is chosen stochastically from the successful events with antibiotic-context-dependent weights. Under antibiotic exposure, resistant events receive an increased selector weight controlled by a parameter weight; without antibiotic exposure, sensitive events receive an increased selector weight controlled by parameters again. Thus, antibiotic exposure favours resistant acquisition and absence of antibiotic exposure favours sensitive acquisition, but neither phenotype is made deterministic unless the configured selector weights make the competing event probability zero. This makes the transmission process graph-based, weight-aware, relation-specific, state-dependent, phenotype-traceable, and stochastic under mixed same-day exposure.

The simulator also supports a superspreader mechanism used in shifted benchmark scenarios. A designated staff member can receive elevated patient-contact intensity over a specified interval, with optional amplification of the associated edge weights. This introduces a temporally localised staff-centred amplification of exposure pressure without changing the underlying graph formalism. The simulator distinguishes latent AMR state *X*_*i*_(*d*) from observed surveillance state *O*_*i*_ (*d*) ∈ {0, 1, 2}, denoting unknown, observed negative, and observed positive respectively. Screening can occur through admission testing, passive routine testing, and active symptom-triggered testing. When universal admission screening is enabled, all newly admitted patients are tested regardless of latent state; when universal admission screening is disabled but symptomatic admission screening is enabled, only newly admitted IS/IR patients are tested. Passive screening is scheduled and can either screen all eligible individuals or a capacity-limited convenience sample of eligible individuals. Eligible individuals include patients and staff, with staff excluded when they are not present on that day; individuals already screened that day or carrying a pending test are not resampled. Active screening is triggered when a patient progresses from colonisation to infection. Test results may be immediate or delayed. If a positive result is delayed, the observation state and any resulting temporary isolation are applied when the pending-result countdown reaches zero, not on the original sampling day. The isolation trigger is configurable: in the baseline it is resistant-only, whereas causal interventions can use an all-positives trigger.

At the end of each day, the graph stores event totals used to define forecasting targets, including resistant acquisition, resistant infection, resistant importation, contact-mediated resistant transmission, and within-host resistant selection. Resistant importation is stored separately for CR and IR admissions and is combined downstream when resistant importation is used as a label component. The exported within-host selection component used for the predictive endpoint is the CR selection total, corresponding to CS →CR events; IS→IR events contribute to the resistant-infection total rather than to a separate exported IR-selection target. The simulator also stores daily resistant burden fractions. These graph-level summaries preserve the mechanism of emergence and allow downstream learning tasks to distinguish importation-driven from endogenously generated resistant burden.

### Graph tensorisation and temporal windows

Each daily GraphML snapshot is converted into a graph-learning object:

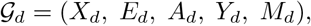

where *X*_*d*_ is the node-feature matrix, *E*_*d*_ is the directed edge-index tensor, *A*_*d*_ is the edge-feature matrix, *Y*_*d*_ contains graph-level horizon labels, and *M*_*d*_ stores metadata such as trajectory identifier, day index, stable node names, roles, and ward-linked information. In the predictive conversion, each fully observed node is encoded by role, latent AMR state, antibiotic exposure, isolation status, same-day new resistant acquisition, and same-day new resistant infection. In the observation-limited predictive representation, the feature dimensionality is retained, but the latent AMR-state channel is replaced by observed positivity and the two latent same-day incident-event channels are set to zero. Thus, unknown and observed-negative individuals are collapsed in the observed-state channel, and the partial-observation model is not given ground-truth same-day acquisition or infection flags. Each edge is represented by contact weight and contact type:

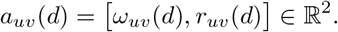

Ward metadata, including home ward and staff multi-ward coverage, is preserved for post hoc translational visualisation but is not injected into the default predictive node-feature vector. Temporal windows are formed from strictly contiguous graphs within the same simulated trajectory:

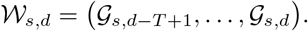

A window is included only when all graphs belong to the same simulation and the day indices are consecutive. Prediction targets are constructed from future graph totals over a horizon *H*, for example

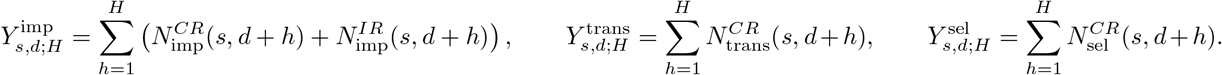

Endogenous resistant burden is defined as

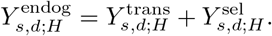

### Forecast labels and mechanism-aware tasks

The primary predictive endpoint is endogenous-versus-importation majority at horizon *H* = 7. It is based on the endogenous share

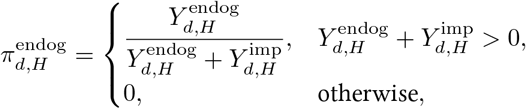

and the corresponding binary label

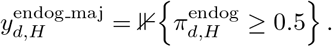

This endpoint asks whether contact-mediated resistant transmission plus within-host resistant selection dominates importation-driven resistant emergence over the future horizon. The converter also stores mechanism-specific future totals, three-way importation/transmission/selection shares, resistant acquisitions, resistant infections, maximum future resistant fraction, and auxiliary labels used for stable pipeline compatibility.

### Temporal graph model

A temporal sample is an ordered sequence of daily directed graphs:

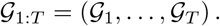

The model combines a daily graph encoder with a temporal Transformer. Each daily graph is encoded by an edge-aware GraphSAGE-style message-passing stack. For node *i* on day *t*, incoming neighbour messages concatenate the neighbour embedding with edge attributes before projection:

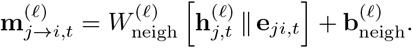

Incoming messages are averaged and combined with a transformed root representation. In the predictive benchmark, GraphSAGE-style neighbour sampling is used during training, with sampled fanouts of up to 15 neighbours at the first sampled layer and 10 at the second sampled layer. In the current causal-policy configuration, true neighbour sampling is disabled and the model is trained on the converted graph windows directly.

Daily node embeddings are pooled into graph-level embeddings using learned attention. The sequence of daily graph embeddings is then passed to a Transformer encoder with learned positional embeddings. The final temporal representation is obtained either by mean pooling across transformed daily tokens or by using a learned classification token. A linear prediction head maps the resulting sequence representation to task-specific outputs.

### Intervention-conditioned causal policy model

For causal policy-response experiments, supervision is aligned to baseline-relative improvement in future transmission-plus-importation resistant burden over horizon *H* = 7. For decision state *d* and action *a*,

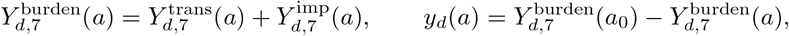

where *a*_0_ denotes the baseline action, 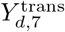 is the horizon sum of new CR transmitted events and 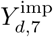 is the horizon sum of new imported CR and IR cases. Larger values therefore correspond to greater predicted improvement. The primary supervised task is grouped oracle-action classification; the continuous gain target is retained for within-state ranking regularisation and downstream evaluation.

The causal policy model extends the temporal graph backbone with two additional inputs: an explicit state-summary vector and an action descriptor. The causal graph conversion uses an enriched node representation relative to the predictive converter. In the ground-truth causal track, nodes carry a 22-dimensional feature vector containing role, one-hot latent AMR state, antibiotic exposure, isolation status, same-day resistant acquisition and infection indicators, ward and ward-coverage features, observation and testing information, pending-test information, admission-screening need, presence status, remaining isolation duration, and admission age. In the partial-observation causal track, nodes carry a 16-dimensional feature vector in which latent state is replaced by observed positivity and observation-knownness, and same-day resistant acquisition and resistant infection channels are set to zero. Edge channels remain the same as in the predictive branch: contact weight and contact type.

The state-summary vector is computed from the terminal graph of the decision window and contains 49 features: node-feature means and standard deviations, staff–patient composition, graph size and edge structure, testing and isolation burden, pending test information, admission-screening need, presence status, admission age, and a nine-feature operational testing-and-isolation context. The manifest-based action descriptor is a fixed 33-dimensional vector containing intervention-family indicators, target-scope indicators, baseline and policy-valid flags, screening cadence, result delay, admission-screening flag, isolation-strength parameters, temporal-bound fields, and hashed action and target identifiers. It does not contain a separate categorical channel for isolation trigger policies parametrization; in the current action menu, actions that differ by resistant-only versus all-positive isolation trigger are separated through their manifest action identifiers and simulator-generated counterfactual outcomes rather than through an explicit trigger-policy feature.

The graph, summary, and action pathways are projected separately and combined through concatenation, element-wise interactions, and absolute differences. A multilayer perceptron produces the intervention-conditioned score 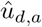. The primary causal loss is state-wise softmax cross-entropy over the available action set:

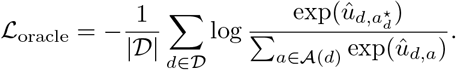

In the current configuration, the auxiliary continuous gain-matching loss is disabled, while a within-state ranking regulariser on the current task remains active. Train, validation, and test partitions are defined by manifest seed ranges, ensuring that all action-conditioned branches from the same simulator trajectory remain in the same partition.

Policy evaluation is performed on held-out decision states with complete action sets and finite oracle quantities. The trained model scores every candidate action, the predicted action is selected by maximal predicted utility, and the oracle-best action is defined by maximal simulator-derived gain. Because multiple actions can share the same oracle gain, evaluation reports both strict tie-broken metrics and tie-aware metrics that count any oracle-best action as correct.

### MILP *proof-of-concept* decision layer for constrained policy selection

To convert intervention-conditioned predictions into explicit operational decisions, we introduce a mixed-integer linear programming [42, 43, 44] layer on top of the held-out action scores. Let 𝒟_dec_ denote the set of retained decision states and let denote the finite intervention library. For each state–action pair (*d, a*) ϵ 𝒟_dec_ × 𝒜, the policy-evaluation stage exports a predicted score 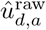. In the present implementation, the optimizer uses a baseline-delta transformation, so the MILP utility is:

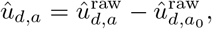

where *a*_0_ is the baseline action for the same decision state. Because the oracle metric is improvement-oriented, larger values are better. We define binary decision variables

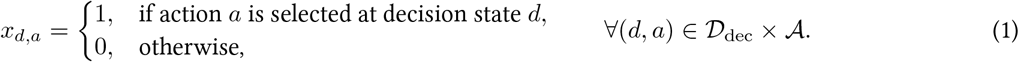

Each action also carries a feasibility-group label used for cooldown and max-use constraints. In the present implementation, identifier-specific rules take precedence when available; otherwise actions inherit group membership from their intervention family. Consequently, all current screening-response variants belong to the same constrained screening family even when they correspond to different screening-frequency templates.

For notational clarity, 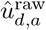 denotes the model-predicted intervention score exported by the policy-evaluation stage for applying action *a* at retained decision state *d*. In the current grouped oracle-action configuration, this score is a task-aware utility used for action ranking rather than a calibrated estimate on the numerical scale of the simulator-derived gain. Larger values nevertheless correspond to actions ranked as more favourable by the trained policy model. The baseline action is denoted by *a*_*0*_ ϵ 𝒜. Each action has an associated cost *c*_*d,a*_ *≥* 0, which may be constant across decision states or vary with the state if state-dependent resource costs are specified. We write

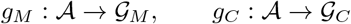

for the action-to-family maps used by the maximum-use and cooldown constraints, respectively. These maps may coincide, but are kept distinct to allow different operational groupings for different constraint types. For a cooldown family *g*, let Δ_*g*_ denote its cooldown length, and let *t*(*d*) denote the calendar day associated with decision state *d*. We define

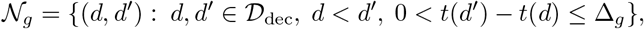

so 𝒩_*g*_ that contains pairs of decision states that cannot both select actions from cooldown family *g*. The optimisation problem then becomes:

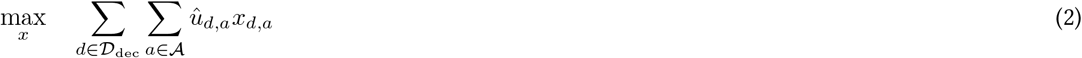

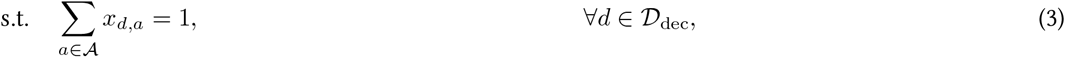

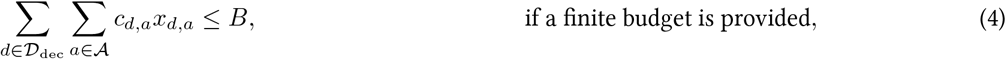

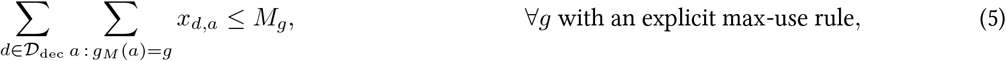

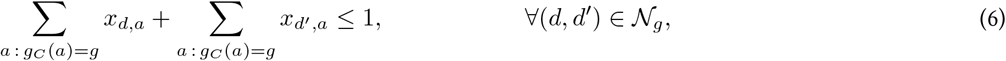

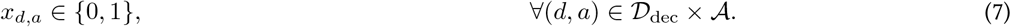

Here *c*_*d,a*_ is the action cost inherited from the rule specification, *B* is an optional global budget, *M*_*g*_ is an optional global maximum number of uses for intervention family *g*, and 𝒩_*g*_ is the set of within-trajectory state pairs whose decision days are separated by no more than the cooldown of family *g*. Costs do not appear as an explicit penalty in the objective; they enter only through optional feasibility constraints.

In the present configuration, the total budget is unconstrained, so the budget constraint is inactive, and all max-use constraints are also inactive. The active feasibility restrictions therefore come from exact-one-action selection and the cooldown rule for the action family represented in the current non-baseline menu. This optimisation layer is intended as a *proof-of-concept* decision module. It does not embed the epidemic dynamics directly within the optimisation problem, nor does it optimise ward-level resource allocation or latent transmission states. Instead, it translates model-predicted intervention gains into feasible selections under explicit operational constraints, yielding a transparent bridge between intervention-conditioned forecasting and constrained decision support. Therefore, the optimization model here connects with the pipeline to provide the end-point of decision making complementing predictions, and its power lies in the ability to encode easily new constraints and scale computationally very efficiently by solving optimization problems that map realistically to equivalent real world settings.

## Discussion

The present platform advances hospital AMR modelling in three aligned ways. First, it preserves mechanistic semantics at the graph level: admissions, discharges, screening delays, observation persistence, isolation, and antibiotic-driven selection are not post hoc annotations, but explicit generative mechanisms. Second, it constructs forecasting tasks directly from those mechanisms, enabling prediction targets that remain clinically interpretable rather than reducing everything to generic incidence proxies. Third, it uses a stable external benchmark that anchors cross-step comparison, making it possible to interpret ablations and train-side perturbation experiments against a common reference.

The strongest conclusion from the study is that the predictive endpoint is highly learnable under matched train-test conditions. In the canonical benchmark, external performance was near-ceiling in both tracks, but it showed a clearer sensitivity and accuracy cost at the fixed operating threshold when data availability was limited. This indicates that the relevant structural and temporal information is present in the evolving hospital graphs and can be learned robustly with latent-state access and, to a lesser degree, under a surveillance-derived observed-positive proxy.

In the present results, removing edge weights did not destroy predictive utility in either track. By contrast, compressing node features was costly in the ground-truth track and much more damaging under partial observation. This suggests that the benchmark is not dependent on edge-weight variation alone, but it does depend on sufficient node-level epidemiological and surveillance context, especially when latent state is unavailable.

The main weakness emerges under calibration and transfer. Step 6 showed that altered surveillance-linked training dynamics can still transfer well, particularly in the ground-truth track, but some partial-observation settings retain AUROC while operating at an unstable threshold. Step 7 further showed that broader regime diversity in training does not automatically stabilise the canonical operating point: the ground-truth sweep preserved excellent AUROC but failed at the threshold, and the partial-observation sweep was weak overall. Step 8 provides an important counterpoint for ground truth, where matched shifted training and testing remained strong, but partial-observation shifted performance was only moderate. The sharper failure is Step 9 under partial observation, where zero-shot transfer from the canonical model to the shifted test set approached random ranking. The implication is that the principal challenge is not only learning within a harder regime, but portability and calibration of a learned decision rule across regimes. That distinction matters in a methodological manner. Several perturbation settings preserved ranking ability while degrading the binary operating point. From a practical perspective, this points toward calibration, threshold retuning, repeated-seed evaluation, and class-balance diagnostics as immediate priorities. It also cautions against reading every performance drop as total loss of predictive structure. In some cases the model still orders risk reasonably well, but the chosen threshold no longer transfers. A further interpretive point concerns the semantics of the partial-observation track and the Step 6 perturbations. In the observation-limited track, the model does not receive a noisy copy of latent AMR state or latent same-day incident-event flags; it receives a surveillance-mediated proxy whose value depends on who is screened, when results become available, and whether positive findings persist in the recorded state. Likewise, the Step 6 experiments are not pure observability stress tests, because altered screening can also change isolation and therefore the epidemic trajectories seen during training. They should therefore be read as joint surveillance-and-response perturbation experiments. That interpretation is more accurate and more useful, because it clarifies that some of the transfer difficulty may arise from changed policy-sensitive dynamics rather than from missingness alone. More broadly, the work supports a view of AMR digital twins as mechanistic decision-support environments rather than black-box prediction tools. Their value lies not only in producing forecasts, but in linking those forecasts to explicit assumptions about admission pressure, patient turnover, contact structure, antibiotic exposure, screening, delayed reporting, and isolation. That makes it possible to ask why a prediction changes, what information it depends on, and how much of it survives under altered observational conditions.

Beyond forecasting future AMR dynamics, the causal policy experiment shows that the framework can be extended to support intervention choice at the level of a decision state. Across both ground-truth and partial-observation tracks, the policy selector achieved useful tie-aware accuracy, high top-2 recovery, and universal tie-aware non-worsening relative to baseline, but strict accuracy and oracle-best-row AUROC were more modest. Partial observation reduced policy-selection accuracy and top-2 recovery, while score-level AUROC remained 0.676 in both tracks. These results suggest that the learned state representation captures decision-relevant structure rather than only predictive signal, but that ranking margin and calibration remain limited under the current policy library. In practical terms, the causal layer indicates that the digital twin can be used not only to anticipate deterioration but also to rank feasible operational responses conditional on the current hospital state.

A second important observation is that the retained test set does not exercise all candidate actions equally. The 2-day same-day screening policy with resistant-only isolation dominates the oracle-best distribution, the baseline comparator accounts for most remaining states, the 2-day all-positive isolation variant appears as a smaller oracle-best group, and the 7-day delayed screening variants are never oracle-best in the retained test states. This pattern supports the model’s ability to recover the dominant decision boundary in the sampled regime, but it weakens any claim that all neighbouring policies have been equally stress-tested. At the same time, these results should be interpreted within the boundaries of the simulated environment. The oracle action is defined with respect to simulator-generated counterfactual outcomes and a restricted intervention library, so further work is needed to assess robustness to alternative reward definitions, broader and more balanced policy spaces, and calibration against real hospital operational data.

There are however also other important limitations. The study is synthetic and, although mechanistically structured, does not yet establish clinical validity in a real hospital environment. The benchmark is intentionally controlled, which improves interpretability but cannot capture the full complexity of routine practice. Several post-baseline analyses suggest problems of threshold transfer under distribution shift, indicating that calibration and validation strategy require more careful study, or training in a wider mixture of mechanistic drivers. Future work should examine repeated-seed robustness, formal calibration, threshold tuning, sensitivity analysis around surveillance assumptions, cleaner decoupling of observation from intervention when the scientific question is specifically about observability, and richer action spaces that couple constrained optimisation more directly to ward and staff level operational levers. A real-world extension would require mapping hospital information systems onto the graph representation, addressing missingness explicitly, and deciding which mechanism-aware endpoints are most useful to IPC teams in practice.

Even with those caveats, the main message is clear. Hospital AMR is a relational and time-dependent problem, and methods that preserve that structure can recover clinically meaningful signals that are difficult to express in static or aggregated form. A digital twin will not eliminate uncertainty from hospital AMR policy decision-making, but it can make the sources of uncertainty more explicit, scenario comparison more robust, and enhanced surveillance imminently actionable. In that sense, this work is best seen as a step toward digital twins that are both mechanistically transparent and clinically interpretable.

## Data Availability

All data produced in the present study are available upon reasonable request to the authors.

## Author contributions

Conceptualization: C.P.T., R.A. Methodology: C.P.T., R.A. Software: C.P.T. Formal analysis: C.P.T. Investigation: C.P.T., R.A. Writing—original draft: C.P.T. Writing—review & editing: R.A. Visualization: C.P.T. Supervision: C.P.T., R.A.

## Data and code availability

The repositories for this line of work can be found at https://github.com/harrytr/dt amr predictive and https://github.com/harrytr/dt amr causal for each experiment separately. We have included single driver scripts to simplify the process of supporting full reproducibility, both for the data and results in each repository respectively.

## References

[1] World Health Organization. Antimicrobial resistance. WHO Fact sheet (Newsroom), 2023. Accessed 2026-02-26.

[2] Centers for Disease Control and Prevention. Antibiotic Resistance Threats in the United States, 2019. Technical report, U.S. Centers for Disease Control and Prevention, 2019. Accessed 2026-02-26.

[3] World Health Organization. Global Antimicrobial Resistance and Use Surveillance System (GLASS). WHO Initiative page, 2025. Accessed 2026-02-26.

[4] Ikuta K et al. Naghavi M, Vollset S. Global burden of bacterial antimicrobial resistance 1990-2021: a systematic analysis with forecasts to 2050. The Lancet, 404(10459):1199–1226, Sep 2024.

[5] Iruka N. Okeke, Marlieke E. A. de Kraker, Thomas P. Van Boeckel, Chirag K. Kumar, Heike Schmitt, Ana C. Gales, Silvia Bertagnolio, Mike Sharland, and Ramanan Laxminarayan. The scope of the antimicrobial resistance challenge. The Lancet, 403(10442):2426–2438, Jun 2024.

[6] Charlotte S. Ho, Carlos T. H. Wong, Thet Tun Aung, Rajamani Lakshminarayanan, Jodhbir S. Mehta, Saaeha Rauz, Alan McNally, Balint Kintses, Sharon J. Peacock, Cesar de la Fuente-Nunez, Robert E. W. Hancock, and Darren S. J. Ting. Antimicrobial resistance: a concise update. The Lancet Microbe, 6(1), Jan 2025.

[7] Derek Cocker, Richard Fitzgerald, Colin S Brown, and Alison Holmes. Protecting healthcare and patient pathways from infection and antimicrobial resistance. BMJ, 387, 2024.

[8] Karina-Doris Vihta, Emma Pritchard, Koen B. Pouwels, Susan Hopkins, Rebecca L. Guy, Katherine Henderson, Dimple Chudasama, Russell Hope, Berit Muller-Pebody, Ann Sarah Walker, David Clifton, and David W. Eyre. Predicting future hospital antimicrobial resistance prevalence using machine learning. Communications Medicine, 4(1):197, Oct 2024.

[9] Yousra Kherabi, Michaël Thy, Donia Bouzid, David B Antcliffe, Timothy Miles Rawson, and Nathan Peiffer-Smadja. Machine learning to predict antimicrobial resistance: future applications in clinical practice? Infect Dis Now, 54(3):104864, February 2024.

[10] Philippe Vanhems, Alain Barrat, Ciro Cattuto, Jean-François Pinton, Nagham Khanafer, Corinne Régis, Byeul-a Kim, Brigitte Comte, and Nicolas Voirin. Estimating potential infection transmission routes in hospital wards using wearable proximity sensors. PLOS ONE, 8(9):1–9, 09 2013.

[11] Laura Temime, Lulla Opatowski, Yohan Pannet, Christian Brun-Buisson, Pierre Yves Boëlle, and Didier Guillemot. Peripatetic health-care workers as potential superspreaders. Proceedings of the National Academy of Sciences, 106(43):18420–18425, 2009.

[12] Donald E. Curtis, Christopher S. Hlady, Gaurav Kanade, Sriram V. Pemmaraju, Philip M. Polgreen, and Alberto M. Segre. Healthcare worker contact networks and the prevention of hospital-acquired infections. PLOS ONE, 8(12):1–11,12 2014.

[13] Eili Y Klein, Katie K Tseng, Jeremiah Hinson, Katherine E Goodman, Aria Smith, Matt Toerper, Joe Amoah, Pranita D Tamma, Scott R Levin, and Aaron M Milstone. The role of healthcare Worker-Mediated contact networks in the transmission of Vancomycin-Resistant enterococci. Open Forum Infect Dis, 7(3):ofaa056, February 2020.

[14] Mariano Ciccolini, Tjibbe Donker, Robin Köck, Martin Mielke, Ron Hendrix, Annette Jurke, Janette Rahamat-Langendoen, Karsten Becker, Hubert G.M. Niesters, Hajo Grundmann, and Alexander W. Friedrich. Infection prevention in a connected world: The case for a regional approach. International Journal of Medical Microbiology, 303(6):380–387, 2013. Special Issue Antibiotic Resistance.

[15] B S Cooper, S P Stone, C C Kibbler, B D Cookson, J A Roberts, G F Medley, G Duckworth, R Lai, and S Ebrahim. Isolation measures in the hospital management of methicillin resistant staphylococcus aureus (mrsa): systematic review of the literature. BMJ, 329(7465):533, 2004.

[16] Quentin J. Leclerc, Audrey Duval, Didier Guillemot, Lulla Opatowski, and Laura Temime. Using contact network dynamics to implement efficient interventions against pathogen spread in hospital settings: A modelling study. PLOS Medicine, 21(7):1–24, 07 2024.

[17] Evangelia Katsoulakis, Qi Wang, Huanmei Wu, Leili Shahriyari, Richard Fletcher, Jinwei Liu, Luke Achenie, Hongfang Liu, Pamela Jackson, Ying Xiao, Tanveer Syeda-Mahmood, Richard Tuli, and Jun Deng. Digital twins for health: a scoping review. npj Digital Medicine, 7(1):77, Mar 2024.

[18] Brant H. Tudor, Ryan Shargo, Geoffrey M. Gray, Jamie L. Fierstein, Frederick H. Kuo, Robert Burton, Joyce T. Johnson, Brandi B. Scully, Alfred Asante-Korang, Mohamed A. Rehman, and Luis M. Ahumada. A scoping review of human digital twins in healthcare applications and usage patterns. npj Digital Medicine, 8(1):587, Sep 2025.

[19] Emanuele Rossi, Ben Chamberlain, Fabrizio Frasca, Davide Eynard, Federico Monti, and Michael Bronstein. Temporal graph networks for deep learning on dynamic graphs, 2020.

[20] Charalampos P. Triantafyllidis and Ricardo Aguas. Causality-aware graph neural networks for functional stratification and phenotype prediction at scale. npj Systems Biology and Applications, 11(1):92, Aug 2025.

[21] Charalampos P. Triantafyllidis, Alessandro Barberis, Fiona Hartley, Ana Miar Cuervo, Enio Gjerga, Philip Charlton, Linda van Bijsterveldt, Julio Saez Rodriguez, and Francesca M. Buffa. A machine learning and directed network optimization approach to uncover TP53 regulatory patterns. iScience, 26(12), Dec 2023.

[22] Krista M English, Joanne M Langley, Allison McGeer, Nathaniel Hupert, Raymond Tellier, Bonnie Henry, Scott A Halperin, Lynn Johnston, and Babak Pourbohloul. Contact among healthcare workers in the hospital setting: developing the evidence base for innovative approaches to infection control. BMC Infect Dis, 18(1):184, April 2018.

[23] Jared K Wilson-Aggarwal, Nick Gotts, Wai Keong Wong, Chris Liddington, Simon Knight, Moira J Spyer, Catherine F Houlihan, Eleni Nastouli, and Ed Manley. Investigating healthcare worker mobility and patient contacts within a UK hospital during the COVID-19 pandemic. Commun Med (Lond), 2(1):165, December 2022.

[24] Ashleigh Myall, James R. Price, Robert L. Peach, Mohamed Abbas, Sid Mookerjee, Nina Zhu, Isa Ahmad, Damien Ming, Farzan Ramzan, Daniel Teixeira, Christophe Graf Andrea Y. Weiße, Stephan Harbarth, Alison Holmes, and Mauricio Barahona. Prediction of hospital-onset covid-19 infections using dynamic networks of patient contact: an international retrospective cohort study. The Lancet Digital Health, 4(8):e573–e583, Aug 2022.

[25] Jessica R. E. Bridgen, Joseph M. Lewis, Stacy Todd, Miriam Taegtmeyer, Chris P. Jewell, Lorenzo Pellis, and Thomas House. A bayesian approach to identifying the role of hospital structure and staff interactions in nosocomial transmission of SARS-CoV-2. Journal of The Royal Society Interface, 21(212):20230525, 2024.

[26] Thomas Hornbeck, David Naylor, Alberto M Segre, Geb Thomas, Ted Herman, and Philip M Polgreen. Using sensor networks to study the effect of peripatetic healthcare workers on the spread of hospital-associated infections. J Infect Dis, 206(10):1549–1557, October 2012.

[27] Rossana Mastrandrea, Alberto Soto-Aladro, Philippe Brouqui, and Alain Barrat. Enhancing the evaluation of pathogen transmission risk in a hospital by merging hand-hygiene compliance and contact data: a proof-of-concept study. BMC Res Notes, 8:426, September 2015.

[28] Christopher Illingworth, J., William L Hamilton, Ben Warne, Matthew Routledge, Ashley Popay, Chris Jackson, Tom Fieldman, Luke W Meredith, Charlotte J Houldcroft, Myra Hosmillo, Aminu S Jahun, Laura G Caller, Sarah L Caddy, Anna Yakovleva, Grant Hall, Fahad A Khokhar, Theresa Feltwell, Malte L Pinckert, Iliana Georgana, Yasmin Chaudhry, Martin D Curran, Surendra Parmar, Dominic Sparkes, Lucy Rivett, Nick K Jones, Sushmita Sridhar, Sally Forrest, Tom Dymond, Kayleigh Grainger, Chris Workman, Mark Ferris, Effrossyni Gkrania-Klotsas, Nicholas M Brown, Michael P Weekes, Stephen Baker, Sharon J Peacock, Ian G Goodfellow, Theodore Gouliouris, Daniela de Angelis, and M Estée Török. Superspreaders drive the largest outbreaks of hospital onset COVID-19 infections. Elife, 10, August 2021.

[29] Luis E. C. Rocha, Vikramjit Singh, Markus Esch, Tom Lenaerts, Fredrik Liljeros, and Anna Thorson. Dynamic contact networks of patients and mrsa spread in hospitals. Scientific Reports, 10(1):9336, Jun 2020.

[30] Alexandre Darbon, Davide Colombi, Eugenio Valdano, Lara Savini, Armando Giovannini, and Vittoria Colizza. Disease persistence on temporal contact networks accounting for heterogeneous infectious periods. Royal Society Open Science, 6(1):181404, 01 2019.

[31] Hannan Tahir, Luis Eduardo López-Cortés, Axel Kola, Dafna Yahav, André Karch Hanjue Xia, Johannes Horn, Konrad Sakowski, Monika J. Piotrowska, Leonard Leibovici, Rafael T. Mikolajczyk, and Mirjam E. Kretzschmar. Relevance of intra-hospital patient movements for the spread of healthcare-associated infections within hospitals-a mathematical modeling study. PLOS Computational Biology, 17(2):1–23, 02 2021.

[32] Tjibbe Donker, Jacco Wallinga, Richard Slack, and Hajo Grundmann. Hospital networks and the dispersal of hospital-acquired pathogens by patient transfer. PLOS ONE, 7(4):1–8, 04 2012.

[33] Elizabeth Scaria, Nasia Safdar, and Oguzhan Alagoz. Validating agent-based simulation model of hospital-associated clostridioides difficile infection using primary hospital data. PLOS ONE, 18(4):1–16, 04 2023.

[34] Tal T. Robin, Jaime Cascante-Vega, Jeffrey Shaman, and Sen Pei. System identifiability in a time-evolving agent-based model. PLOS ONE, 19(1):1–15, 01 2024.

[35] Sen Pei, Dwayne Seeram, Seth Blumberg, Bo Shopsin, Anne-Catrin Uhlemann, and Jeffrey Shaman. Inferring asymptomatic carriers of antimicrobial-resistant organisms in hospitals using genomic, microbiological and patient mobility data. Nature Communications, 16(1):10140, Nov 2025.

[36] Audrey Duval, Quentin J. Leclerc, Didier Guillemot, Laura Temime, and Lulla Opatowski. An algorithm to build synthetic temporal contact networks based on close-proximity interactions data. PLOS Computational Biology, 20(6):1–23, 06 2024.

[37] Genevieve Coorey, Gemma A. Figtree, David F. Fletcher, Victoria J. Snelson, Stephen Thomas Vernon, David Winlaw, Stuart M. Grieve, Alistair McEwan, Jean Yee Hwa Yang, Pierre Qian, Kieran O’Brien, Jessica Orchard, Jinman Kim, Sanjay Patel, and Julie Redfern. The health digital twin to tackle cardiovascular disease—a review of an emerging interdisciplinary field. npj Digital Medicine, 5(1):126, Aug 2022.

[38] David Drummond and Apolline Gonsard. Definitions and characteristics of patient digital twins being developed for clinical use: Scoping review. J Med Internet Res, 26:e58504, November 2024.

[39] Yanping Zheng, Lu Yi, and Zhewei Wei. A survey of dynamic graph neural networks. Frontiers of Computer Science, 19(6):196323, Dec 2024.

[40] William L. Hamilton, Rex Ying, and Jure Leskovec. Inductive representation learning on large graphs. In Advances in Neural Information Processing Systems 30 (NeurIPS 2017), pages 1024–1034, 2017.

[41] Ashish Vaswani, Noam Shazeer, Niki Parmar, Jakob Uszkoreit, Llion Jones, Aidan N Gomez,Łukasz Kaiser, and Illia Polosukhin. Attention is all you need. In I. Guyon, U. Von Luxburg, S. Bengio, H. Wallach, R. Fergus, S. Vishwanathan, and R. Garnett, editors, Advances in Neural Information Processing Systems, volume 30. Curran Associates, Inc., 2017.

[42] Dimitris Bertsimas and John N Tsitsiklis. Introduction to linear optimization. Athena Scientific, Belmont, Mass, 1997.

[43] Charalampos P Triantafyllidis and Lazaros G Papageorgiou. An integrated platform for intuitive mathematical programming modeling using latex. PeerJ Computer Science, 4:e161, 2018.

[44] Charalampos P Triantafyllidis and Nikolaos Samaras. A new non-monotonic infeasible simplex-type algorithm for linear programming. PeerJ Computer Science, 6:e265, 2020.

