## Supplementary Information for "A digital twin for hospital antimicrobial resistance forecasting and constrained intervention optimisation"

Research Article

**Charalampos P. Triantafyllidis***
Nuffield Department of Medicine,
University of Oxford, Oxford, OX3 7BN, UK


**Ricardo Aguas**
Nuffield Department of Medicine,
University of Oxford, Oxford, OX3 7BN, UK


May 15, 2026

#### Supplementary Methods: full simulator, tensorisation, and models

This supplement gives the complement to the implementation-level details corresponding to the *Methods* of the main manuscript. It preserves the full mathematical specification of graph generation, admission importation, transmission, surveillance, feature construction, temporal learning, causal policy modelling and constrained intervention selection.

##### S1. Daily hospital graph construction

**Nodes, roles, wards, and multi-ward staffing.** The simulator generates a sequence of daily directed weighted graphs

$$G_d = (V_d, E_d), \qquad d = 1, \dots, D,$$

where $V_d = P_d \cup S$ consists of patients $P_d$ present on day $d$ and staff $S$. Each patient $i \in P_d$ is assigned to exactly one ward

$$w(i) \in \{1, \dots, W\},$$

whereas each staff member $s \in S$ is assigned a home ward $w(s)$ together with a multi-ward coverage set

$$\mathcal{W}(s) \subseteq \{1, \dots, W\}, \qquad w(s) \in \mathcal{W}(s).$$

This induces a role-aware and ward-aware graph in which within-ward interactions are dense, but cross-ward connectivity also arises through staff whose coverage sets span multiple wards. Three contact classes are represented. First, within each ward $w$, patient–patient contacts are generated on the induced patient set

$$P_d^{(w)} = \{i \in P_d : w(i) = w\}.$$

The implementation samples a directed scale-free graph on $P_d^{(w)}$, so the ward-level patient subgraph is sparse and heterogeneous.

Second, staff–patient contacts are generated in two stages within each ward. Let $S_d^{(w)} \subseteq S_d^{\mathrm{act}}$, where $S_d^{\mathrm{act}} \subseteq S$ is the set of active staff present that day, denote the active staff assigned to ward $w$. In a first coverage stage, patients in $P_d^{(w)}$ are assigned in shuffled round-robin fashion to staff in $S_d^{(w)}$, ensuring that each patient in a staffed ward has at least one staff contact on that day. For each such assignment, the graph includes both $s \to i$ and $i \to s$, reflecting asymmetric but bidirectional interaction opportunities.

In a second augmentation stage, each staff member $s \in S_d^{(w)}$ receives additional ward-specific patient contacts up to a target load

$$k_{s,w} = \min\Big\{n_w,\ \max\big(m_{\mathrm{sp}}, \lfloor \phi_{\mathrm{sp}}\, n_w \rfloor\big)\Big\},$$

where $n_w = |P_d^{(w)}|$, $\phi_{\rm sp}$ is the staff–patient contact fraction, and $m_{\rm sp}$ is a minimum contact floor. Extra contacts are sampled without replacement from patients in ward $w$ not already linked to $s$ during the coverage stage. This produces daily staff–patient mixing that guarantees patient coverage in staffed wards while preserving sparse and heterogeneous contact structure.

Third, staff–staff interactions are sampled more sparsely. Each unordered pair $\{s,t\} \subseteq S_d^{\rm act}$ is included with probability $p_{\rm ss}$, yielding a sparse staff mixing layer. Each directed edge $e=(u,v) \in E_d$ carries a relation type $r_{uv}(d)$ and weight $\omega_{uv}(d)$. In the implementation,

$$r_{uv}(d) \in \{0,1,2\},$$

corresponding respectively to patient–patient, staff–patient in either direction, and staff–staff contacts. Edge weights are continuous random intensities:

$$\omega_{uv}(d) \sim \begin{cases} \mathrm{Unif}(0.3,1.0), & r_{uv}(d)=0, \\ \mathrm{Unif}(0.6,1.2), & u \in S,\ v \in P_d, \\ \mathrm{Unif}(0.2,0.8), & u \in P_d,\ v \in S, \\ \mathrm{Unif}(0.3,0.9), & r_{uv}(d)=2. \end{cases}$$

Hence the graph encodes not only adjacency but also interaction class and strength. Formally, the daily hospital graph is therefore a marked directed graph

$$G_d = \big(V_d, E_d, \{r_{uv}(d)\}_{(u,v)\in E_d}, \{\omega_{uv}(d)\}_{(u,v)\in E_d}\big).$$

**Dynamic census, admissions, and discharges.** The patient set evolves over time through ward-specific discharge and admission. Let $n_w(d) = |P_d^{(w)}|$. The number of discharges from ward $w$ on day $d$ is

$$\kappa_w(d) = \min\Big\{n_w(d),\ \max\big(\lfloor \rho_{\rm dis}\, n_w(d)\rfloor,\ m_{\rm dis}\big)\Big\},$$

where $\rho_{\rm dis}$ is the daily proportional turnover and $m_{\rm dis}$ is a minimum per-ward discharge floor. A uniformly sampled subset of $\kappa_w(d)$ patients is removed from ward $w$, and the same number of new patients is admitted into that ward, so that ward-level census remains approximately stable. Newly admitted patients receive fresh persistent identifiers and are initialised with latent AMR state, antibiotic status, isolation status, observation status, and event indicators. This makes node identity stable over time for continuing individuals, while still allowing realistic churn in the patient population.

##### S2. Admission importation and seasonal modulation

At admission, each new patient is assigned one of five entry states:

$$X_{\rm adm}(d) \in \{U, CS, CR, IS, IR\}.$$

The implementation parameterises importation using four non-negative state weights for colonised-sensitive, colonised-resistant, infected-sensitive, and infected-resistant admission states,

$$p_0^{CS}, \qquad p_0^{CR}, \qquad p_0^{IS}, \qquad p_0^{IR},$$

corresponding to the configuration fields `p_admit_import_cs`, `p_admit_import_cr`, `p_admit_import_is`, and `p_admit_import_ir`. When these fields are absent, the corresponding initial-importation parameters are used as fallbacks where available. Each weight can be modulated by a temporal multiplier $m(d)$ and capped independently:

$$u^x(d) = \min\{\bar{p}^x,\ p_0^x m(d)\}, \qquad x \in \{CS, CR, IS, IR\},$$

where $\bar{p}^x$ is the state-specific safety cap. The simulator supports several forms for $m(d)$. In the constant case,

$$m(d) = 1.$$

Under sinusoidal modulation with amplitude $a \in [0,1)$, period $T_{\rm sea}$, and phase $\varphi$,

$$m(d) = 1 + a \sin\Big(\frac{2\pi(d-\varphi)}{T_{\rm sea}}\Big).$$

Under a piecewise seasonal regime,

$$m(d) = \begin{cases} m_{\rm high}, & d \in \mathcal{H}, \\ m_{\rm low}, & d \notin \mathcal{H}, \end{cases}$$

where $\mathcal{H}$ is the designated high-season interval, possibly wrapping around the end of the cycle. Under a shock regime,

$$m(d) = \begin{cases} m_{\text{shock}}, & d_0 \le d \le d_0 + \ell - 1, \\ 1, & \text{otherwise}, \end{cases}$$

for a randomly sampled start day $d_0$, duration $\ell$, and shock multiplier $m_{\text{shock}} > 1$.

The four non-residual admission weights are sampled jointly rather than through sequential CR-before-CS assignment. Let

$$S(d) = u^{CS}(d) + u^{CR}(d) + u^{IS}(d) + u^{IR}(d).$$

The realised probabilities are

$$\pi^x(d) = \begin{cases} u^x(d), & S(d) \le 1, \\ \dfrac{u^x(d)}{S(d)}, & S(d) > 1, \end{cases} \qquad x \in \{CS, CR, IS, IR\},$$

and the residual uncolonised/uninfected probability is

$$\pi^U(d) = 1 - \sum_{x \in \{CS,CR,IS,IR\}} \pi^x(d).$$

A newly admitted patient is then drawn from this categorical distribution. Thus, if the total CS/CR/IS/IR admission pressure is below one, uncolonised admissions occupy the remaining probability mass; if the requested weights exceed one, the implementation preserves their relative composition while normalising the imported-state mass to one. This construction supports stable, seasonal, and shock-like exogenous pressure while avoiding order-dependent dominance of whichever imported state is checked first. For downstream resistant-importation targets, CR and IR importation are both counted as resistant importation.

##### S3. AMR state dynamics and antibiotic exposure

Each individual $i \in V_d$ occupies a latent AMR state

$$X_i(d) \in \mathcal{X} = \{U, CS, CR, IS, IR\},$$

where $U$ denotes uncolonised, $CS$ and $CR$ denote sensitive and resistant colonisation, and $IS$ and $IR$ denote sensitive and resistant infection. In parallel, each individual carries an antibiotic exposure state

$$A_i(d) \in \{0, 1, \ldots, C\},$$

where $A_i(d) = 0$ means no current treatment and $A_i(d) = c > 0$ denotes treatment under antibiotic class $c$. Antibiotic initiation and discontinuation are stochastic. If $A_i(d) = 0$, then

$$\Pr\big(A_i(d+1) > 0 \mid X_i(d) \in \{IS, IR\}\big) = p_{\text{start}}^{\text{inf}},$$

whereas for non-infected individuals

$$\Pr\big(A_i(d+1) > 0 \mid X_i(d) \notin \{IS, IR\}\big) = p_{\text{start}}^{\text{noninf}}.$$

When treatment is initiated, the class is sampled uniformly from $\{1, \ldots, C\}$. If $A_i(d) > 0$, then treatment stops with probability

$$\Pr\big(A_i(d+1) = 0 \mid A_i(d) > 0\big) = p_{\text{stop}}^{\text{abx}}.$$

Thus antibiotic exposure is not static, but a time-varying endogenous process coupled to infection status.

The latent-state dynamics combine within-host selection, progression from colonisation to infection, and stochastic clearance. Conditional on antibiotic exposure, sensitive colonisation may become resistant colonisation:

$$\Pr\big(X_i(d+1) = CR \mid X_i(d) = CS,\ A_i(d) > 0\big) = p_{\text{sel}}^{\text{col}}.$$

Similarly, sensitive infection may become resistant infection:

$$\Pr\big(X_i(d+1) = IR \mid X_i(d) = IS,\ A_i(d) > 0\big) = p_{\text{sel}}^{\text{inf}}.$$

These transitions represent treatment-associated selection rather than transmission. Colonised individuals may progress to infection while retaining bacterial phenotype:

$$\Pr\big(X_i(d+1) = IS \mid X_i(d) = CS\big) = p_{\text{prog}}, \qquad \Pr\big(X_i(d+1) = IR \mid X_i(d) = CR\big) = p_{\text{prog}}.$$

Infected individuals clear to the uncolonised state with probability

$$\Pr\big(X_i(d+1) = U \mid X_i(d) \in \{IS, IR\}\big) = p_{\text{clr}}^{\text{inf}},$$

and colonised individuals clear with probability

$$\Pr\big(X_i(d+1) = U \mid X_i(d) \in \{CS, CR\}\big) = p_{\text{clr}}^{\text{col}}.$$

Hence resistant emergence may be generated by at least three distinct mechanisms: direct resistant importation at admission, resistant transmission through the contact graph, and treatment-associated selection. The simulator contains both $CS \to CR$ and $IS \to IR$ treatment-associated transitions. For the mechanism-specific forecasting targets used downstream, however, the explicit selection-total channel is $N_{\text{sel}}^{CR}$, which records $CS \to CR$ resistant-colonisation selection; $IS \to IR$ is represented through the same-day resistant-infection indicator rather than through a separate $N_{\text{sel}}^{IR}$ graph-level total.

##### S4. Edge-weighted transmission and source-state infectivity

Transmission is evaluated over directed incoming contacts in the daily hospital graph. Let $i$ be an uncolonised target with $X_i(d) = U$, and let

$$\mathcal{N}_i^-(d) = \{j : (j, i) \in E_d\}$$

denote the set of incoming neighbours on day $d$. Only upstream agents in colonised or infected states can transmit, so the infectious source set for target $i$ is

$$\mathcal{I}_i^-(d) = \{j \in \mathcal{N}_i^-(d) : X_j(d) \in \{CS, CR, IS, IR\}\}.$$

For an incoming edge $j \to i$, the baseline transmission coefficient depends on relation type:

$$\beta_{ji} = \begin{cases} \beta_{pp}, & r_{ji}(d) = 0, \\ \beta_{sp}, & r_{ji}(d) = 1, \\ \beta_{ss}, & r_{ji}(d) = 2. \end{cases}$$

Here $r_{ji}(d)$ denotes patient–patient, staff–patient, or staff–staff contact type, and $\omega_{ji}(d) \geq 0$ denotes the edge weight. The current simulator separates clinical-state infectivity from resistance phenotype. The source-state multiplier is

$$\gamma(X_j(d)) = \begin{cases} 0.5, & X_j(d) = CS, \\ 0.5, & X_j(d) = CR, \\ 1.0, & X_j(d) = IS, \\ 1.0, & X_j(d) = IR, \end{cases}$$

so colonised carriers are less infectious than infected carriers within both sensitive and resistant bacterial phenotypes. Resistance is retained as a separate multiplicative term,

$$\eta_R^{\mathbf{1}\{X_j(d) \in \{CR, IR\}\}},$$

but in the present experiments

$$\eta_R = 1.0,$$

so resistant carriage or infection is not made globally more transmissible than sensitive carriage or infection.

Isolation and cross-ward hand hygiene attenuate the realised edge-level exposure probability after the baseline exponential dose-response calculation. If $q_j(d), q_i(d) \in \{0, 1\}$ indicate whether source and target are isolated, respectively, the isolation factor is

$$\zeta_{ji}(d) = \lambda^{q_j(d) + q_i(d)}, \qquad 0 < \lambda \leq 1,$$

where $\lambda$ is the isolation transmission multiplier. Cross-ward staff-mediated contacts can also be attenuated by a leaky hand-hygiene factor. Let

$$\eta_H(j, i, d) = \begin{cases} 0.55, & \text{if cross-ward hand hygiene is enabled and } j \to i \text{ is a cross-ward staff–patient contact}, \\ 1, & \text{otherwise}. \end{cases}$$

This reduces the effective transmission contribution of cross-ward staff contacts without deleting the contact edge.

The baseline infectious pressure before isolation and hand-hygiene attenuation is

$$\Lambda_{ji}^0(d) = \beta_{ji}\, \omega_{ji}(d)\, \gamma(X_j(d))\, \eta_R^{\mathbf{1}\{X_j(d) \in \{CR, IR\}\}}.$$

The unattenuated edge-level exposure probability is

$$p_{ji}^{0}(d) = 1 - \exp\{-\Lambda_{ji}^{0}(d)\}.$$

The probability actually used for the Bernoulli exposure draw is then

$$p_{ji}(d) = \min\big\{1, \max\big[0, p_{ji}^{0}(d)\,\zeta_{ji}(d)\,\eta_H(j,i,d)\big]\big\}\,.$$

Thus, isolation and hand hygiene are multiplicative attenuators of the edge-level probability rather than terms placed inside the exponential force of infection. Assuming conditional independence of edge-level exposure events given the daily graph and agent states, the probability that target $i$ receives no contact-mediated acquisition on day $d$ is

$$P_i^{\text{no acq}}(d) = \prod_{j \in \mathcal{I}_i^-(d)} \left(1 - p_{ji}(d)\right),$$

and therefore

$$P_i^{\text{acq}}(d) = 1 - P_i^{\text{no acq}}(d) = 1 - \prod_{j \in \mathcal{I}_i^-(d)} \left(1 - p_{ji}(d)\right).$$

Because attenuation and clipping occur after the baseline edge-level dose-response calculation, the implementation is represented by this product over edge-level Bernoulli probabilities rather than by collapsing all attenuated terms into a single summed force of infection.

The simulator is event-resolved rather than only aggregate-count based. For each infectious edge $j \to i$, a Bernoulli exposure event

$$B_{ji}(d) \sim \text{Bernoulli}(p_{ji}(d))$$

is sampled. When $B_{ji}(d) = 1$, a transmission event is recorded with source, target, source state, transmitted bacterial phenotype, target context, edge type, edge weight, isolation status, and antibiotic context. The transmitted phenotype associated with source $j$ is

$$\phi_j(d) = \begin{cases} \text{sens}, & X_j(d) \in \{CS, IS\}, \\ \text{res}, & X_j(d) \in \{CR, IR\}. \end{cases}$$

Let the set of successful exposure events received by target $i$ on day $d$ be

$$\mathcal{E}_i(d) = \{e = (j \to i) : j \in \mathcal{I}_i^-(d),\ B_{ji}(d) = 1\}.$$

If $\mathcal{E}_i(d) = \varnothing$, the target remains uncolonised through the contact-mediated transmission step. If exactly one successful event occurs, the newly colonised target inherits that event's bacterial phenotype. If multiple successful exposure events occur, including same-day competition between sensitive and resistant phenotypes, the implementation chooses one infecting event using antibiotic-context-dependent weights. With $\kappa_S$ denoting `no_abx_sensitive_selection_weight` and $\kappa_R$ denoting `abx_resistant_selection_weight`, each successful event $e \in \mathcal{E}_i(d)$ receives selector weight

$$a_e(d) = \begin{cases} \kappa_S, & A_i(d) = 0 \text{ and } \phi_e = \text{sens}, \\ 1, & A_i(d) = 0 \text{ and } \phi_e = \text{res}, \\ 1, & A_i(d) > 0 \text{ and } \phi_e = \text{sens}, \\ \kappa_R, & A_i(d) > 0 \text{ and } \phi_e = \text{res}. \end{cases}$$

The infecting event is sampled with probability

$$\Pr(e \text{ chosen} \mid \mathcal{E}_i(d)) = \frac{a_e(d)}{\sum_{e' \in \mathcal{E}_i(d)} a_{e'}(d)}.$$

The target becomes $CS$ if the chosen event is sensitive and $CR$ if the chosen event is resistant. Thus, antibiotic exposure favours resistant acquisition and absence of antibiotic exposure favours sensitive acquisition, but neither competing phenotype is made impossible unless its realised selector weight is zero. This makes transmission graph-based, weight-aware, relation-specific, state-dependent, phenotype-traceable, and stochastic under mixed same-day exposure.

##### S5. Superspreader dynamics

The simulator also supports an explicit superspreader mechanism used in the shifted benchmark. In this mode, a designated staff member is assigned an elevated contact intensity over a specified day interval. Operationally, this increases the number of patient contacts sampled by that staff member and can also amplify the associated edge weights. Let $s^\star$ denote the designated superspreader and let $\mathcal{T}^\star$ denote the active day interval. During days $d \in \mathcal{T}^\star$, the simulator scales the sampled patient-contact load for $s^\star$ and may apply an additional multiplicative factor to the corresponding edge weights. This mechanism does not alter the core graph formalism above, but it introduces a temporally localised, staff-centred amplification of exposure pressure that is useful for constructing more severe shift scenarios.

##### S6. Isolation, delayed surveillance, and observation persistence

The simulator distinguishes latent AMR state $X_i(d)$ from observed surveillance state $O_i(d)$, where

$$O_i(d) \in \{0, 1, 2\}$$

denotes unknown, observed negative, and observed positive, respectively. Screening can occur through admission testing, passive routine testing, and active symptom-triggered testing. Routine passive screening is triggered on scheduled surveillance days, either weekly on a fixed weekday or every $k$ days. When a positive passive-screening capacity is supplied, only a convenience sample of eligible individuals is screened within the configured passive-screening capacity; when the capacity is zero, all eligible individuals on a scheduled screening day are tested. Eligible individuals include patients and staff, with staff excluded when they are not present on that day. Admission screening can be enabled separately and is symptomatic-only by default, so only newly admitted patients already in $IS$ or $IR$ are admission-screen eligible under that setting. Active screening is additionally triggered when a patient progresses from colonisation to infection, so symptomatic progression can initiate testing even outside the passive screening schedule.

Given a test on day $d$, positivity is determined from latent colonisation or infection state using sensitivity $\mathrm{Se}$ and specificity $\mathrm{Sp}$:

$$\Pr(\text{test+} \mid X_i(d) \in \{CS, CR, IS, IR\}) = \mathrm{Se}, \qquad \Pr(\text{test+} \mid X_i(d) = U) = 1 - \mathrm{Sp}.$$

Results may be immediate or delayed by $L$ days. If a positive result is returned, then the observation state becomes

$$O_i(d + L) = 2,$$

whereas a negative result yields

$$O_i(d + L) = 1.$$

Under persistent observation, the most recent recorded result is carried forward until replaced by a later test; under non-persistent observation, observation fields are reset more aggressively, so the visible state can revert to unknown in the absence of fresh testing.

Observed positive status can trigger temporary isolation according to a configurable trigger rule. In the current baseline, isolation is resistant-only: isolation is activated only when the returned positive is associated with a resistant latent state. Candidate causal interventions can instead use an all-positives trigger, under which any positive result activates isolation. If the trigger condition is met for a test sampled on day $d$ with reporting delay $L$, isolation is activated when the result is applied on day $d + L$. For a fixed duration $\tau_{\text{iso}}$, this corresponds to

$$q_i(d') = 1, \qquad d' \in \{d + L, \ldots, d + L + \tau_{\text{iso}} - 1\},$$

up to the simulator's daily timer update. Isolation then feeds back into transmission through the multiplier $\zeta_{ji}(d)$ above, so surveillance affects future spread mechanistically rather than merely altering what is measured.

##### S7. Mechanism-specific graph totals

At the end of each day, the graph stores event totals that are later used to define prediction tasks. Let

$$N_{\text{acq}}^{CR}(d) = \sum_{i \in V_d} \mathbf{1}\{\text{new resistant acquisition at } i \text{ on day } d\},$$

$$N_{\text{inf}}^{IR}(d) = \sum_{i \in V_d} \mathbf{1}\{\text{new resistant infection at } i \text{ on day } d\}.$$

Admission importation totals are stored separately for all four non-residual imported states,

$$N_{\text{imp}}^{CS}(d), \qquad N_{\text{imp}}^{CR}(d), \qquad N_{\text{imp}}^{IS}(d), \qquad N_{\text{imp}}^{IR}(d).$$

The resistant importation total used downstream is therefore

$$N_{\text{imp}}^{R}(d) = N_{\text{imp}}^{CR}(d) + N_{\text{imp}}^{IR}(d).$$

Contact-mediated transmission totals are stored for sensitive and resistant colonisation events,

$$N_{\text{trans}}^{CS}(d), \qquad N_{\text{trans}}^{CR}(d),$$

and the mechanism-specific resistant-selection total used by the prediction labels is

$$N_{\text{sel}}^{CR}(d),$$

which records $CS \to CR$ within-host selection. Infected-state selection $IS \to IR$ contributes to the same-day resistant-infection indicator $N_{\text{inf}}^{IR}(d)$, but the checked-in graph totals do not export a separate $N_{\text{sel}}^{IR}(d)$ mechanism channel. The simulator also stores the daily resistant fraction

$$R(d) = \frac{N_{CR}(d) + N_{IR}(d)}{N_{CS}(d) + N_{CR}(d) + N_{IS}(d) + N_{IR}(d)},$$

where $N_{CS}(d), N_{CR}(d), N_{IS}(d), N_{IR}(d)$ are state counts on day $d$. These graph-level summaries preserve the mechanism of emergence and make it possible to define downstream tasks that distinguish importation-driven from endogenously generated resistant burden.

##### S8. Graph tensorisation and temporal windows

Each daily GraphML snapshot is converted into a graph-learning object

$$\mathcal{G}_d = (X_d,\ E_d,\ A_d,\ Y_d,\ M_d),$$

where $X_d \in \mathbb{R}^{n_d \times p}$ is the node-feature matrix, $E_d \in \mathbb{N}^{2 \times m_d}$ is the directed edge-index tensor, $A_d \in \mathbb{R}^{m_d \times q}$ is the edge-feature matrix, $Y_d$ contains graph-level horizon labels, and $M_d$ stores metadata such as trajectory identifier, day index, stable node names and roles, and ward-linked node metadata. Here $n_d = |V_d|$ and $m_d = |E_d|$.

In the fully observed predictive representation, each node $i \in V_d$ is encoded by the feature vector

$$x_i(d) = \big[\mathbf{1}\{i \in S\},\ X_i^{\text{enc}}(d),\ A_i(d),\ q_i(d),\ \mathbf{1}\{\text{new CR acquisition at } i \text{ on } d\},\ \mathbf{1}\{\text{new IR infection at } i \text{ on } d\}\big],$$

where $X_i^{\text{enc}}(d) \in \{0, 1, 2, 3, 4\}$ is the numeric encoding of the latent AMR state $X_i(d)$, $A_i(d)$ is the antibiotic class indicator, and $q_i(d) \in \{0, 1\}$ denotes isolation status. Thus $p = 6$ node channels are used in this representation. The node-feature matrix is

$$X_d = \begin{bmatrix} x_1(d)^\top \\ \vdots \\ x_{n_d}(d)^\top \end{bmatrix} \in \mathbb{R}^{n_d \times 6}.$$

A second representation is used to study observation-limited learning. In that setting, the latent-state channel is replaced by a binary observed-positivity encoding,

$$\widetilde{X}_i^{\text{enc}}(d) = \mathbf{1}\{O_i(d) = 2\}.$$

Hence unknown and observed-negative individuals are collapsed into the same value 0, while observed-positive individuals retain value 1. The feature dimensionality is retained, but the two same-day latent incident-event channels are zero-filled in the partial-observation track:

$$\tilde{x}_i(d) = \big[\mathbf{1}\{i \in S\},\ \mathbf{1}\{O_i(d) = 2\},\ A_i(d),\ q_i(d),\ 0,\ 0\big].$$

Thus, the partial-observation model is not given ground-truth same-day resistant acquisition or resistant-infection indicators. This produces a principled degradation of observability without altering the tensor shape expected by the predictive model.

Each directed edge $e = (u, v) \in E_d$ is encoded by

$$a_{uv}(d) = \big[\omega_{uv}(d),\ r_{uv}(d)\big] \in \mathbb{R}^2,$$

so

$$A_d = \begin{bmatrix} a_{e_1}(d)^\top \\ \vdots \\ a_{e_{m_d}}(d)^\top \end{bmatrix} \in \mathbb{R}^{m_d \times 2}.$$

The learning model therefore receives both interaction intensity and interaction class rather than only bare adjacency.

In addition to these model-input tensors, the conversion step preserves ward-aware metadata for each node. For node $i$, the converter stores a scalar home-ward identifier

$$w_i^{\text{home}} \in \{1, \ldots, W\},$$

together with a serialised representation of the full ward-coverage set

$$\mathcal{W}_i \subseteq \{1, \ldots, W\},$$

and its cardinality

$$c_i = |\mathcal{W}_i|.$$

For patients, $w_i^{\mathrm{home}}$ coincides with the unique ward assignment and $\mathcal{W}_i$ is a singleton. For staff, $w_i^{\mathrm{home}}$ records the designated home ward while $\mathcal{W}_i$ records the full multi-ward coverage set inherited from the simulator. These quantities are preserved as metadata rather than injected directly into the default learning feature vector, so they can be used later for post hoc aggregation and translational visualisation without changing the core predictive representation.

The graph object also stores trajectory-level metadata. Each daily graph is assigned a stable simulation identifier $s$ and day index $d$, so that a temporal sequence can be written

$$\{\mathcal{G}_{s,1}, \mathcal{G}_{s,2}, \ldots, \mathcal{G}_{s,D_s}\}.$$

For clarity, when graph totals are evaluated within a specific simulated trajectory, we write the same quantities with explicit trajectory index $s$, for example $N_{\mathrm{acq}}^{CR}(s,d)$, $N_{\mathrm{inf}}^{IR}(s,d)$, $N_{\mathrm{imp}}^{CR}(s,d)$, $N_{\mathrm{imp}}^{IR}(s,d)$, $N_{\mathrm{trans}}^{CR}(s,d)$, $N_{\mathrm{sel}}^{CR}(s,d)$, and $R(s,d)$.

Prediction targets are constructed from future graph totals over a horizon $H$. For example, for a graph observed on day $d$, the future resistant-acquisition target is

$$Y_{s,d;H}^{CR} = \sum_{h=1}^{H} N_{\mathrm{acq}}^{CR}(s, d+h),$$

the future resistant-infection target is

$$Y_{s,d;H}^{IR} = \sum_{h=1}^{H} N_{\mathrm{inf}}^{IR}(s, d+h),$$

and the mechanism-specific horizon totals are

$$Y_{s,d;H}^{\mathrm{imp}} = \sum_{h=1}^{H} \left[ N_{\mathrm{imp}}^{CR}(s, d+h) + N_{\mathrm{imp}}^{IR}(s, d+h) \right], \qquad Y_{s,d;H}^{\mathrm{trans}} = \sum_{h=1}^{H} N_{\mathrm{trans}}^{CR}(s, d+h), \qquad Y_{s,d;H}^{\mathrm{sel}} = \sum_{h=1}^{H} N_{\mathrm{sel}}^{CR}(s, d+h).$$

The endogenous resistant burden over the horizon is then

$$Y_{s,d;H}^{\mathrm{endog}} = Y_{s,d;H}^{\mathrm{trans}} + Y_{s,d;H}^{\mathrm{sel}}.$$

Mechanism shares are formed whenever the denominator is non-zero, for example

$$\pi_{s,d;H}^{\mathrm{trans}} = \frac{Y_{s,d;H}^{\mathrm{trans}}}{Y_{s,d;H}^{\mathrm{trans}} + Y_{s,d;H}^{\mathrm{imp}}}, \qquad \pi_{s,d;H}^{\mathrm{endog}} = \frac{Y_{s,d;H}^{\mathrm{endog}}}{Y_{s,d;H}^{\mathrm{endog}} + Y_{s,d;H}^{\mathrm{imp}}}.$$

For early-warning style tasks, the horizon resistant-fraction signal is defined as the maximum future resistant fraction within the next $H$ days:

$$Y_{s,d;H}^{R} = \max_{1 \le h \le H} R(s, d+h).$$

Temporal windows are then formed from contiguous graphs within the same trajectory. For window length $T$ and sliding step $\Delta$,

$$\mathcal{W}_{s,d} = \left( \mathcal{G}_{s,d-T+1}, \ldots, \mathcal{G}_{s,d} \right),$$

with windows included only if all graphs belong to the same simulation $s$ and satisfy strict day contiguity,

$$(d-T+1), (d-T+2), \ldots, d.$$

Equivalently, if the day indices in a candidate window are $(t_1, \ldots, t_T)$, the admissibility condition is

$$t_{k+1} = t_k + 1, \qquad k = 1, \ldots, T-1.$$

This ensures that each learning example corresponds to a genuine temporally ordered hospital history rather than an artificial concatenation of unrelated snapshots. The resulting supervised dataset is therefore a collection of pairs

$$\left( \mathcal{W}_{s,d},\ Y_{s,d;H} \right),$$

where the target $Y_{s,d;H}$ may be scalar, vector-valued, continuous, or binary, depending on the forecasting task.

##### S9. Forecast labels and mechanism-aware tasks

For any horizon $H$, the converter aggregates future event totals over days $d+1,\ldots,d+H$ and stores them on the tensor corresponding to day $d$. For readability, we suppress the trajectory index $s$ in the formulas below when the target is defined pointwise within a fixed trajectory. Let

$$Y_{d,H}^{\text{imp}} = \sum_{k=1}^{H} \left[N_{\text{imp}}^{CR}(d+k) + N_{\text{imp}}^{IR}(d+k)\right], \qquad Y_{d,H}^{\text{trans}} = \sum_{k=1}^{H} N_{\text{trans}}^{CR}(d+k), \qquad Y_{d,H}^{\text{sel}} = \sum_{k=1}^{H} N_{\text{sel}}^{CR}(d+k),$$

$$Y_{d,H}^{\text{endog}} = Y_{d,H}^{\text{trans}} + Y_{d,H}^{\text{sel}}.$$

The primary task used in this study is endogenous-versus-importation majority at horizon $H=7$. It is based on the endogenous share

$$\pi_{d,H}^{\text{endog}} = \begin{cases} \dfrac{Y_{d,H}^{\text{endog}}}{Y_{d,H}^{\text{endog}} + Y_{d,H}^{\text{imp}}}, & Y_{d,H}^{\text{endog}} + Y_{d,H}^{\text{imp}} > 0, \\ 0, & \text{otherwise}, \end{cases}$$

and the corresponding binary label

$$y_{d,H}^{\text{endog\_maj}} = \mathbb{1}\left\{\pi_{d,H}^{\text{endog}} \geq 0.5\right\}.$$

This endpoint asks whether endogenous resistant emergence, defined here as contact-mediated resistant transmission plus within-host resistant selection, dominates importation-driven resistant emergence over the future horizon. Importation-driven resistant emergence includes both CR and IR importation on admission.

In addition, the converter stores three-way shares. Let

$$T_{d,H}^{\text{all}} = Y_{d,H}^{\text{imp}} + Y_{d,H}^{\text{trans}} + Y_{d,H}^{\text{sel}}.$$

When $T_{d,H}^{\text{all}} > 0$,

$$\pi_{d,H}^{\text{imp}} = \frac{Y_{d,H}^{\text{imp}}}{T_{d,H}^{\text{all}}}, \qquad \pi_{d,H}^{\text{trans},3} = \frac{Y_{d,H}^{\text{trans}}}{T_{d,H}^{\text{all}}}, \qquad \pi_{d,H}^{\text{sel}} = \frac{Y_{d,H}^{\text{sel}}}{T_{d,H}^{\text{all}}},$$

and otherwise all three shares are set to zero. The framework also stores standard future totals such as resistant acquisitions, resistant infections, maximum resistant fraction, and several auxiliary or placeholder outputs designed to preserve a stable interface for future work. These are implemented but are not primary endpoints unless explicitly reported as such.

##### S10. Temporal graph model

Let a patient-contact trajectory over a window of length $T$ be represented as an ordered sequence of daily directed graphs

$$\mathcal{G}_{1:T} = (\mathcal{G}_1, \ldots, \mathcal{G}_T), \qquad \mathcal{G}_t = (V_t, E_t).$$

For each day $t$, every node $i \in V_t$ carries a feature vector $\mathbf{x}_{i,t} \in \mathbb{R}^F$, and every directed edge $(j,i) \in E_t$ carries an edge-feature vector $\mathbf{e}_{ji,t} \in \mathbb{R}^S$. The model maps the sequence $\mathcal{G}_{1:T}$ to a prediction by combining two components: a spatial encoder that learns a representation for each daily graph, and a temporal encoder that models dependencies across days.

Each daily graph is encoded independently by a stack of edge-aware GraphSAGE-style message-passing layers. Denote by

$$\mathbf{h}_{i,t}^{(0)} = \mathbf{x}_{i,t}$$

the input representation of node $i$ on day $t$. At graph-convolution layer $\ell$, each incoming neighbour $j \in \mathcal{N}_t^-(i)$ sends a message to node $i$ given by

$$\mathbf{m}_{j\to i,t}^{(\ell)} = W_{\text{neigh}}^{(\ell)} \left[\mathbf{h}_{j,t}^{(\ell)} \,\|\, \mathbf{e}_{ji,t}\right] + \mathbf{b}_{\text{neigh}}^{(\ell)},$$

where $\|$ denotes concatenation. Messages from all incoming neighbours are aggregated by arithmetic mean,

$$\bar{\mathbf{m}}_{i,t}^{(\ell)} = \frac{1}{|\mathcal{N}_t^-(i)|} \sum_{j \in \mathcal{N}_t^-(i)} \mathbf{m}_{j\to i,t}^{(\ell)},$$

and combined with a separate linear transformation of the node's current state,

$$\tilde{\mathbf{h}}_{i,t}^{(\ell+1)} = W_{\text{root}}^{(\ell)} \mathbf{h}_{i,t}^{(\ell)} + \mathbf{b}_{\text{root}}^{(\ell)} + \bar{\mathbf{m}}_{i,t}^{(\ell)}.$$

The layer output is then

$$\mathbf{h}_{i,t}^{(\ell+1)} = \mathrm{Dropout}\Big(\mathrm{LayerNorm}\Big(\mathrm{ReLU}\Big(\tilde{\mathbf{h}}_{i,t}^{(\ell+1)}\Big)\Big)\Big).$$

Thus, the spatial encoder follows the GraphSAGE principle of neighbourhood aggregation, but augments each neighbour contribution with edge attributes before projection. This is particularly relevant here because transmission-relevant interactions are carried not only by node states, but also by edge-level information describing the contact relation itself.

After $L_{\mathrm{GNN}}$ such layers, each node has a final spatial embedding

$$\mathbf{h}_{i,t} \in \mathbb{R}^D.$$

In the predictive benchmark, these GraphSAGE updates were not evaluated on the full in-neighbourhood of every destination node during training. Instead, predictive training used GraphSAGE-style neighbour sampling, so each optimization step operated on sampled multi-hop subgraphs with fanouts of up to 15 neighbours at the first sampled layer and up to 10 neighbours at the second sampled layer. Concretely, random seed nodes were selected from each training graph, their sampled two-hop computation graphs were constructed, and message passing was applied on those induced subgraphs. In the checked-in causal policy configuration, true neighbour sampling is disabled and `max_neighbors` is set to zero, so the causal-policy model is trained without this sampled-neighbour approximation. This distinction preserves the shared GraphSAGE learning formulation while matching the runtime configuration used in each branch.

The node embeddings of day $t$ are pooled into a single graph-level representation by learned attention. For each node,

$$\mathbf{u}_{i,t} = \tanh(P\mathbf{h}_{i,t} + \mathbf{b}_P), \qquad s_{i,t} = \mathbf{w}^\top \mathbf{u}_{i,t} + b_s,$$

and the attention coefficient is obtained by a graph-wise softmax,

$$\alpha_{i,t} = \frac{\exp(s_{i,t})}{\sum_{k \in V_t} \exp(s_{k,t})}.$$

The daily graph embedding is then

$$\mathbf{z}_t = \sum_{i \in V_t} \alpha_{i,t} \mathbf{h}_{i,t} \in \mathbb{R}^D.$$

This readout is permutation-invariant with respect to node ordering and allows the model to assign higher weight to nodes whose local state is more informative for downstream prediction. The learned coefficients $\alpha_{i,t}$ also provide a direct interpretability mechanism by quantifying each node's contribution to the day-level representation.

The sequence of daily embeddings is arranged into

$$Z = \begin{bmatrix} \mathbf{z}_1 \\ \mathbf{z}_2 \\ \vdots \\ \mathbf{z}_T \end{bmatrix} \in \mathbb{R}^{T \times D}.$$

Because self-attention is permutation-equivariant, temporal order is injected through learned positional embeddings. Let $\mathbf{p}_1, \ldots, \mathbf{p}_T \in \mathbb{R}^D$ denote these positional vectors. The Transformer input is

$$\mathbf{r}_t^{(0)} = \mathbf{z}_t + \mathbf{p}_t, \qquad t = 1, \ldots, T.$$

The temporal encoder is a stack of $L_{\mathrm{Tr}}$ Transformer encoder blocks. In each block, multi-head self-attention first computes interactions between all pairs of days in the window. For attention head $m$,

$$Q^{(m)} = RW_Q^{(m)}, \qquad K^{(m)} = RW_K^{(m)}, \qquad V^{(m)} = RW_V^{(m)},$$

where $R \in \mathbb{R}^{T \times D}$ collects the current token representations row-wise. The head output is

$$\mathrm{Attn}^{(m)}(R) = \mathrm{softmax}\left(\frac{Q^{(m)} K^{(m)\top}}{\sqrt{d_m}}\right) V^{(m)}.$$

The multi-head outputs are concatenated and linearly projected. As in the standard Transformer encoder, this attention sublayer is followed by a position-wise feed-forward network, with residual connections, layer normalisation, and dropout applied around the sublayers. No causal mask is imposed, so every day within the window may attend to every other day. This is appropriate here because the task is sequence-to-one prediction from a fully observed retrospective window rather than autoregressive next-step generation.

The output of the final Transformer layer is a sequence

$$\widetilde{Z} = \begin{bmatrix} \tilde{\mathbf{z}}_1 \\ \vdots \\ \tilde{\mathbf{z}}_T \end{bmatrix} \in \mathbb{R}^{T \times D}.$$

Two sequence-level summarisation modes are supported. In one mode, a learned classification token $\mathbf{c} \in \mathbb{R}^D$ is prepended before the positional embeddings are added; the final representation is then the transformed CLS token. In the other mode, the final temporal representation is obtained by mean pooling across the $T$ transformed daily tokens:

$$\bar{\mathbf{z}} = \frac{1}{T} \sum_{t=1}^{T} \tilde{\mathbf{z}}_t.$$

The resulting representation, whether CLS-based or mean-pooled, is passed through a linear head

$$\hat{\mathbf{y}} = W_{\text{out}} \bar{\mathbf{z}} + \mathbf{b}_{\text{out}}$$

or, in the CLS case,

$$\hat{\mathbf{y}} = W_{\text{out}} \tilde{\mathbf{c}} + \mathbf{b}_{\text{out}}.$$

##### S11. Intervention-conditioned causal policy model

For the current causal policy-response experiments, supervision is aligned to baseline-relative improvement in future transmission-plus-importation resistant burden over horizon $H = 7$. For decision state $d$ and action $a$, the implemented horizon burden is

$$Y_{d,7}^{\text{burden}}(a) = \sum_{h=1}^{7} \left[ N_{\text{trans}}^{CR}(d+h, a) + N_{\text{imp}}^{CR}(d+h, a) + N_{\text{imp}}^{IR}(d+h, a) \right],$$

equivalently $Y_{d,7}^{\text{burden}}(a) = Y_{d,7}^{\text{trans}}(a) + Y_{d,7}^{\text{imp}}(a)$ with $Y^{\text{imp}}$ defined as CR plus IR importation. The baseline-relative gain target is

$$y_d(a) = Y_{d,7}^{\text{burden}}(a_0) - Y_{d,7}^{\text{burden}}(a),$$

where $a_0$ denotes the baseline action. Larger values therefore correspond to greater predicted improvement. The primary supervised task is grouped oracle-action classification; the continuous gain target is retained for within-state ranking regularization and downstream evaluation. Ties are resolved once at dataset construction by a fixed action ordering and the same stored oracle label is reused throughout training and strict evaluation.

We formalised the causal policy-response task as intervention-conditioned temporal forecasting from hospital graph sequences. Let

$$\mathcal{W}_d = (G_{d-T+1}, \ldots, G_d), \qquad T = 7,$$

denote the observed graph window at decision state $d$. Let $x_a \in \mathbb{R}^p$ denote the action-feature vector attached to action $a$, and let $s_d \in \mathbb{R}^q$ denote an explicit state-summary vector derived from the same decision window. The temporal graph backbone produces a graph-history embedding $z_d \in \mathbb{R}^h$. In the present implementation, action conditioning is therefore based on three inputs: the temporal graph embedding, the action descriptor, and a summary of the operational testing-and-isolation context.

The action descriptor encodes intervention type, target scope, screening cadence, result delay, admission-screening flag, isolation-strength parameters, temporal bounds, and target identity. In the checked-in feature encoding it does not include an explicit `isolation_trigger_policy` channel. However, actions remain distinguishable through the encoded action-identifier fields, including hashed action-name, action-ID, and target-identity features. Thus, sibling interventions that differ by trigger policy are not forced to share an identical action vector, even though trigger policy is not represented as a dedicated semantic channel. The state-summary vector aggregates moments of the node features, staff–patient composition, graph size and edge structure, current new-case indicators when available, testing burden, pending-test countdowns and, where present, pending-test results, admission-screening need, presence status, isolation load, admission age, and an explicit nine-feature operational-context block recording the active screening cadence, weekly screening phase, admission-screening flag, reporting delay, isolation parameters, observation persistence, whether the current day is a screening day, and the time to the next scheduled screen. In the ground-truth track, the pending test result is also retained explicitly; in the partial-observation track, the representation remains restricted to what is observable from surveillance.

At the level of the daily graph tensor, the enriched causal representation did not alter the raw edge channels: each directed contact edge retained the same two attributes as in the predictive branch, namely contact weight and edge type. The augmentation occurred on the node and graph side. In the partial-observation track, each node was represented by a 16-dimensional vector comprising role indicator, observed-positive status, antibiotic exposure, isolation status, zero-filled same-day new-resistant-acquisition and new-resistant-infection channels, normalized ward identity and ward-cover count, an observation-known indicator, a screened-today flag, normalized days since last test, a normalized pending-test countdown, an admission-screening requirement indicator, a presence indicator, normalized remaining isolation days, and normalized admission age. In the ground-truth track, the node vector was expanded to 22 dimensions by replacing the observed-status channel with a five-state AMR one-hot encoding and by retaining both observed-positive and observation-known indicators together with the pending-test-result flag.

From these daily tensors we formed a 49-dimensional explicit state-summary vector composed of the first six node-feature means and standard deviations, staff–patient composition, burden and new-case moments, graph size and connectivity summaries, edge-weight and edge-type summaries, testing and isolation burden summaries, and the nine-dimensional operational-context block. Action descriptors were encoded in a fixed 33-dimensional vector containing intervention-family indicators, target-scope indicators, normalized screening cadence, reporting-delay, multiplier, isolation, admission-screening, temporal-bound fields, and hashed action-name, action-ID, and target-identity fields. The checked-in vector contains no dedicated trigger-policy channel, although trigger-policy variants can still be separated indirectly through the action-identifier encodings.

The graph, summary, and action pathways are projected separately,

$$\tilde{z}_d = \phi_g(z_d), \qquad \tilde{s}_d = \phi_s(s_d), \qquad \tilde{x}_a = \phi_a(x_a), \qquad \bar{z}_d = \tfrac{1}{2}(\tilde{z}_d + \tilde{s}_d),$$

and the prediction head receives the joint representation

$$r_{d,a} = \big[\tilde{z}_d,\ \tilde{s}_d,\ \bar{z}_d,\ \tilde{x}_a,\ \tilde{z}_d \odot \tilde{s}_d,\ |\tilde{z}_d - \tilde{s}_d|,\ \bar{z}_d \odot \tilde{x}_a,\ |\bar{z}_d - \tilde{x}_a|\big],$$

where $\odot$ denotes element-wise multiplication. A multilayer perceptron then produces the intervention-conditioned score

$$\hat{u}_{d,a} = f_\theta(r_{d,a}).$$

Let $a_d^\star$ denote the simulator-defined oracle-best action for decision state $d$. The primary causal loss is a state-wise softmax cross-entropy over the action set available at that state,

$$\mathcal{L}_{\text{oracle}} = -\frac{1}{|\mathcal{D}|}\sum_{d\in\mathcal{D}} \log \frac{\exp(\hat{u}_{d,a_d^\star})}{\sum_{a\in\mathcal{A}(d)} \exp(\hat{u}_{d,a})}.$$

In the current checked-in configuration, the auxiliary continuous gain-matching loss is disabled, but a within-state pairwise/listwise ranking regularizer on `y_h7_trans_import_res_gain` remains active with weight 1, pairwise margin 0.02, and minimum target gap 0.01. The total causal training objective is therefore

$$\mathcal{L}_{\text{total}} = \mathcal{L}_{\text{oracle}} + \mathcal{L}_{\text{rank}},$$

where $\mathcal{L}_{\text{rank}}$ denotes the within-state ranking regularizer.

Train, validation, and test partitions are defined by the manifest generated during causal dataset construction. Specifically, all samples inherit the split label of the seed from which they were generated, so that all action-conditioned branches from the same underlying simulator trajectory remain in the same partition. In the present configuration, those seed ranges are 4000–4019 for training, 5000–5003 for validation, and 6000–6009 for testing.

After model fitting, policy evaluation is performed on held-out decision states that retain one valid row per candidate action and finite oracle quantities. For each such state $\mathcal{W}_d$, the trained model is queried once for every action $a \in \mathcal{A}$, producing a set of predicted intervention-conditioned utilities

$$\{\hat{u}_{d,a} : a \in \mathcal{A}\}.$$

The predicted action is

$$\hat{a}_d = \arg\max_{a\in\mathcal{A}} \hat{u}_{d,a},$$

and the oracle-best action is

$$a_d^\star = \arg\max_{a\in\mathcal{A}} y_d(a),$$

with ties again broken by the fixed action ordering recorded in the manifest for strict-label evaluation. Because multiple actions can share the same oracle gain, policy evaluation reports both tie-aware metrics, which count any oracle-best action as correct, and strict metrics, which compare against the fixed tie-broken oracle label stored in the manifest. It also records top-2 accuracy, regret, and whether the selected action improves on baseline. The per-state action-score tables forwarded to the decision layer are restricted to this same evaluable state set, so downstream optimisation operates on exactly the states used for held-out policy assessment.

### A Parameter catalogue for the Predictive Experiment

#### A.1 Configuration parameters

Table 1 lists the high-level simulator configuration used throughout the endogenous-versus-importation benchmark, including the hospital size, staff allocation, superspreader settings, and importation seasonality controls. It provides the broadest view of the synthetic environment in which all downstream experiments were conducted.

Table 1: High-level configuration parameters for the endogenous-versus-importation benchmark.

| Parameter | Value | Meaning |
|---|---|---|
| num_regions | 1 | Number of hospital regions simulated. |
| num_wards | 10 | Number of wards in the hospital simulation. |
| num_patients | 200 | Number of patient agents. |
| num_staff | 300 | Number of staff agents. |
| export_yaml | True | Export YAML metadata from the simulator. |
| export_gif | False | Whether animated GIF output is produced. Disabled here. |
| override_staff_wards_per_staff | True | Whether to explicitly override staff-to-ward assignment count. |
| staff_wards_per_staff | 2 | Number of wards each staff member is assigned to when override is active. |
| p_import_cs | 0.3 | Simulator default day-0 CS seeding weight, used unless explicitly overridden. |
| p_import_cr | 0.1 | Simulator default day-0 CR seeding weight, used unless explicitly overridden. |
| p_import_is | 0.3 | Simulator default day-0 IS seeding weight, used unless explicitly overridden. |
| p_import_ir | 0.1 | Simulator default day-0 IR seeding weight, used unless explicitly overridden. |
| p_admit_import_cs | 0.15 | Simulator default CS admission-importation weight; canonical and shifted families override this value. |
| p_admit_import_cr | 0.1 | Simulator default CR admission-importation weight; canonical and shifted families override this value. |
| p_admit_import_is | 0.15 | Simulator default IS admission-importation weight; canonical and shifted families override this value. |
| p_admit_import_ir | 0.1 | Simulator default IR admission-importation weight; canonical and shifted families override this value. |

| Parameter | Value | Meaning |
|---|---|---|
| enable_superspreader | False | Enables a designated superspreader mechanism in the global simulator configuration. The canonical benchmark leaves this disabled. |
| superspreader_staff | ”” | Identifier of the staff member designated as superspreader. Empty here in the global configuration. |
| superspreader_state | ”IR” | Colonisation/infection state assigned to the superspreader. |
| superspreader_staff_contacts | 50 | Number of staff contacts for the superspreader profile. |
| superspreader_patient_frac_mult | 3.0 | Multiplier on patient-contact fraction for the superspreader. |
| superspreader_patient_min_add | 10 | Minimum additional patient contacts for the superspreader. |
| superspreader_edge_weight_mult | 1.5 | Multiplier on transmission edge weights for the superspreader. |
| superspreader_start_day | 1 | Day on which superspreader activity starts. |
| superspreader_end_day | 9999 | Day on which superspreader activity ends. Effectively unbounded here. |
| enable_admit_import_seasonality | False | Enables seasonal modulation of admission importation probabilities in the global simulator configuration. Disabled in the canonical benchmark. |
| admit_import_seasonality | ”none” | Seasonality mode; no seasonality is active in the canonical global configuration. |
| admit_import_period_days | 7 | Period of seasonality cycle in days when used. |
| admit_import_pmax_cs | 1.0 | Maximum CS import weight under seasonal modulation. |
| admit_import_pmax_cr | 1.0 | Maximum CR import weight under seasonal modulation. |
| admit_import_pmax_is | 1.0 | Maximum IS import weight under seasonal modulation. |
| admit_import_pmax_ir | 1.0 | Maximum IR import weight under seasonal modulation. |
| admit_import_amp | 0.5 | Amplitude for sinusoidal seasonality when enabled. |
| admit_import_phase_day | 0 | Phase offset for sinusoidal seasonality. |
| admit_import_high_start_day | 1 | Start day of high-importation period in piecewise mode. |
| admit_import_high_end_day | 180 | End day of high-importation period in piecewise mode. |
| admit_import_high_mult | 1.5 | Multiplier during the high-importation interval. |
| admit_import_low_mult | 1.0 | Multiplier during the low-importation interval. |
| admit_import_shock_min_days | 7 | Minimum duration of a shock episode in shock mode. |

| Parameter | Value | Meaning |
|---|---|---|
| admit_import_shock_max_days | 30 | Maximum duration of a shock episode in shock mode. |
| admit_import_shock_mult_min | 1.5 | Minimum shock multiplier. |
| admit_import_shock_mult_max | 5.0 | Maximum shock multiplier. |
| beta_res_mult | 1.0 | Resistant phenotype multiplier used in transmission. The current benchmark keeps this neutral, so resistance is not globally more infectious. |
| beta_state_mult_cs | 0.5 | Source-state transmission multiplier for susceptible colonisation. |
| beta_state_mult_cr | 0.5 | Source-state transmission multiplier for resistant colonisation. |
| beta_state_mult_is | 1.0 | Source-state transmission multiplier for susceptible infection. |
| beta_state_mult_ir | 1.0 | Source-state transmission multiplier for resistant infection. |
| crossward_hand_hygiene_enabled | 1 | Enables leaky hand-hygiene attenuation for cross-ward staff–patient transmission. |
| crossward_hand_hygiene_mult | 0.55 | Multiplicative attenuation applied to eligible cross-ward staff–patient transmission contributions. |
| passive_screen_capacity_per_week | 0 | Weekly passive-screening capacity. A value of zero denotes no capacity cap, so all eligible individuals on a scheduled screening day may be tested. |
| canonical screen_every_k_days | 7 | Canonical routine screening cadence used in Steps 1–5, held fixed in Steps 7–8, and used as the fixed cadence in the Step 6 delay arm. |
| canonical screen_result_delay_days | 2 | Canonical reporting delay used in Steps 1–5, held fixed in Steps 7–8, and used as the fixed delay in the Step 6 frequency arm. |
| canonical screen_on_admission | 1 | Universal admission screening is enabled in the canonical predictive benchmark and fixed unless a step explicitly varies screening. |
| canonical persist_observations | 1 | Observed surveillance statuses persist across days in the predictive benchmark. |
| active_symptom_screening | 1 | Enables active testing when a patient progresses from carriage to infection. |
| admission_screen_symptomatic_only | 1 | Enables symptomatic-only admission testing of newly admitted IS/IR patients when universal admission screening is off; universal admission screening still tests all newly admitted patients. |

| Parameter | Value | Meaning |
| --- | --- | --- |
| isolation_trigger_policy | "resistant_only" | Baseline isolation trigger policy; only positive tests associated with resistant latent states initiate isolation. |

##### A.2 Conversion parameters

Table 2 reports the conversion-layer settings used to turn daily graphs into tensorised learning objects. These settings are intentionally simple because the central benchmark varies mainly through simulation regime and training conditions rather than through alternative conversion logic.

Table 2: Conversion parameters for graph-to-tensor preprocessing.

| Parameter | Value | Meaning |
| --- | --- | --- |
| horizons | "7" | Prediction horizon(s) passed to conversion; default horizon is 7 days. |
| workers | None | Conversion worker count is left unset in the driver, so execution falls back to the runtime default rather than using a fixed hard-coded worker count. |

##### A.3 Model and training parameters

Table 3 summarises the model architecture, optimisation settings, scheduler and early-stopping controls, and attribution-export settings used in the main benchmark. These values define the temporal window, embedding size, Transformer depth, GraphSAGE depth, optimisation schedule, post hoc full-graph attribution pass, and the validation-driven stopping logic used after neighbour-sampled training.

Table 3: Model and training parameters for the endogenous-versus-importation task.

| Parameter | Value | Meaning |
| --- | --- | --- |
| use_task_hparams | False | If True, downstream training uses task-specific default hyperparameters. |
| train_model | True | Whether model training is executed in standard train-and-evaluate mode. |
| task | "endogenous_importation_majority_h7" | Classification task used for prediction and label validation. |
| pred_horizon | 7 | Default prediction horizon when not inferred from task name. |
| early_outbreak_fixed_threshold | 0.55 | Fixed threshold used only for the alternative early-outbreak task family; retained in the configuration although not used in the endogenous-versus-importation benchmark. |
| T | 7 | Temporal input window length. |
| T_list | "7" | Default comma-separated list of temporal window sizes for multi-T runs. |

| Parameter | Value | Meaning |
|---|---|---|
| sliding_step | 1 | Sliding-window stride used in sequence construction. |
| hidden | 64 | Hidden embedding dimension. |
| heads | 2 | Number of attention heads. |
| dropout | 0.3 | Dropout probability. |
| transformer_layers | 2 | Number of Transformer layers. |
| sage_layers | 2 | Number of GraphSAGE layers. |
| use_cls | False | Whether a CLS-style token/representation is used. |
| batch_size | 16 | Batch size for training. |
| epochs | 50 | Number of training epochs. |
| lr | 1e-4 | Learning rate. |
| max_neighbors | 20 | Maximum neighborhood size used when neighbor sampling is not in true sampled mode. |
| neighbor_sampling | True | Master switch for GraphSAGE-style neighbor sampling. |
| num_neighbors | ”15,10” | Per-layer neighbor sampling fan-out when sampling is enabled. |
| seed_count | 256 | Number of seed nodes/anchors used by sampling loader. |
| seed_strategy | ”random” | Seed-selection strategy for sampling. |
| seed_batch_size | 64 | Batch size for seed nodes in neighbor sampling mode. |
| max_sub_batches | 4 | Maximum number of sub-batches under sampling mode. |
| attn_top_k | 10 | Number of top attention relationships retained for analysis/reporting. |
| attn_rank_by | ”abs_diff” | Criterion used to rank attention relationships. |
| fullgraph_attribution_pass | True | Enables the dedicated full-graph attribution pass used after training for translational explanation export. |
| emit_translational_figures | True | Exports translational attribution figures from the post hoc full-graph inference pass. |
| translational_top_k | 20 | Number of highest-ranked translational attribution items retained for reporting. |
| early_stopping | True | Enables validation-based early stopping during training. |
| patience | 7 | Number of validation checks without sufficient improvement before stopping. |
| min_delta | 1e-4 | Minimum validation improvement required to reset early-stopping patience. |
| save_best_only | True | Restores and retains the best validation checkpoint rather than the final epoch by default. |
| lr_scheduler_on_plateau | True | Enables learning-rate reduction when validation progress stalls. |
| lr_scheduler_factor | 0.5 | Multiplicative reduction factor for the plateau scheduler. |
| lr_scheduler_patience | 3 | Number of validation checks without improvement before reducing learning rate. |
| lr_scheduler_min_lr | 1e-6 | Lower bound for the scheduled learning rate. |

###### Attribution-export interpretation

Although the main predictive runs use GraphSAGE-style neighbour sampling for computational tractability, translational attribution figures are not computed by aggregating sampled subgraph attention directly. Instead, after training, the saved checkpoint is evaluated in a dedicated full-graph attribution pass, and node-attention coefficients from that pass are used to build the ward- and staff-level translational summaries. The resulting figures should therefore be interpreted as full-graph post hoc explanations of a model trained under neighbour sampling, rather than as sampled-attribution approximations or causal estimates of ward- or staff-level effect.

##### A.4 Test and validation parameters

Table 4 collects the test-side and validation-side control settings. Although some of these are retained mainly for workflow compatibility, they remain part of the executable benchmark configuration and therefore should be stated explicitly.

Table 4: Test and validation parameters used in the benchmark workflow.

| Parameter | Value | Meaning |
|---|---|---|
| `test_frac_per_class` | `0.5` | Legacy fraction argument retained for compatibility; canonical baseline construction ignores it. |
| `min_per_class` | `10` | Minimum class count target for testing logic in related workflows. |
| `seed` | `1337` | Random seed used for test-related operations. |
| `balance_tolerance` | `1` | Allowed absolute class-count imbalance when balanced testing is enforced. |
| `require_balanced_test` | `False` | Whether the test set must be exactly or near-balanced. |

##### A.5 Step 1 canonical trajectory parameters

###### Global Step 1 settings

Table 5 records the global settings governing the number and duration of canonical simulations in Step 1. These values define the scale of the raw trajectory pool before any training-side perturbations are applied.

Table 5: Global Step 1 settings for canonical trajectory generation.

| Parameter | Value | Meaning |
|---|---|---|
| `n_sims_per_trajectory` | `10` | Number of simulation replicates generated for each canonical trajectory family. |
| `num_days` | `60` | Number of days per simulation in Step 1. |
| `outbreak_expected_label_min_frac` | `0.3` | Retained threshold for the alternative early-outbreak task family; unused in the endogenous-versus-importation benchmark but still part of the executable configuration. |

| Parameter | Value | Meaning |
|---|---|---|
| outbreak_require_two_class_pooled_baseline | True | Retained safeguard for the alternative early-outbreak task family; unused here but still part of the executable configuration. |

##### Canonical train/test trajectory families

Table 6 is the core regime table for the benchmark. It shows exactly how the endogenous-favouring and importation-favouring canonical families differ through admission importation and discharge turnover on both the training and test sides. Admission-importation entries are CS/CR/IS/IR state weights passed to the joint importation sampler. If their total exceeds one, the simulator normalises them to preserve composition; otherwise the residual mass corresponds to uncolonised admissions.

Table 6: Canonical Step 1 train/test trajectory families for the endogenous-versus-importation benchmark.

| Trajectory | seed_base | p_admit_import CS/CR/IS/IR | daily_discharge_frac | daily_discharge_min_per_ward |
|---|---|---|---|---|
| endog_high_train | 4100 | 0.005/0.005/0.005/0.005 | 0.02 | 0 |
| import_high_train | 5100 | 0.60/0.60/0.60/0.60 | 0.25 | 1 |
| endog_high_test | 6100 | 0.005/0.005/0.005/0.005 | 0.02 | 0 |
| import_high_test | 7100 | 0.60/0.60/0.60/0.60 | 0.25 | 1 |

Interpretation:

- The endog_high trajectories are designed to have very low CS/CR/IS/IR admission-importation weights and low discharge turnover, favouring endogenous spread.
- The import_high trajectories use high CS/CR/IS/IR admission-importation weights and higher discharge turnover, favouring importation-dominant resistant emergence. Because the four weights sum above one, they act as composition weights under saturated importation rather than as independent probabilities.

#### A.6 Step 6 and Step 7 perturbation parameters

Step 6 regenerates training cohorts under altered surveillance assumptions while keeping the Step 4 epidemiological regimes fixed. In the active driver profile, each delay or frequency setting generates paired endogenous-favouring and importation-favouring cohorts using the same admission-importation and turnover values as the canonical Step 1 training families. Step 7 instead varies the epidemiological regime itself by sweeping admission-importation and turnover settings while holding the canonical surveillance policy fixed.

Table 7: Active Step 6 surveillance-perturbation settings.

| Component | Value | Meaning |
|---|---|---|
| DT_STEP6_PROFILE | canonical_baseline | Active default profile; the legacy shifted profile exists only as an optional override and is not the default design. |

| Component | Value | Meaning |
|---|---|---|
| num_days | 60 | Step 6 training cohorts have the same temporal length as the canonical Step 1 trajectories. |
| n_sims_per_regime | 10 | Number of simulations generated for each endogenous/importation regime at each delay or frequency setting. |
| Delay arm | screen_result_delay_days = 0,2,5 | Reporting delay is varied while screen_every_k_days = 7, screen_on_admission = 1, and persist_observations = 1 are held fixed. |
| Frequency arm | screen_every_k_days = 3,7,14 | Routine screening frequency is varied while screen_result_delay_days = 2, screen_on_admission = 1, and persist_observations = 1 are held fixed. |
| Endogenous-favouring regime | CS/CR/IS/IR = 0.005/0.005/0.005/0.005; discharge=0.02; min=0 | Same admission-importation and turnover regime as the Step 1 endogenous-favouring training family. |
| Importation-favouring regime | CS/CR/IS/IR = 0.60/0.60/0.60/0.60; discharge=0.25; min=1 | Same admission-importation and turnover regime as the Step 1 importation-favouring training family. |

Table 8: Active Step 7 epidemiological-regime sweep settings.

| Band | daily_discharge_frac | p_admit_import CS/CR/IS/IR | replicates | Fixed surveillance policy |
|---|---|---|---|---|
| 1 | 0.05 | 0.05/0.05/0.05/0.05 | 4 | screen=7, delay=2, admission=1, persist=1 |
| 2 | 0.08 | 0.08/0.08/0.08/0.08 | 4 | screen=7, delay=2, admission=1, persist=1 |
| 3 | 0.10 | 0.12/0.12/0.12/0.12 | 4 | screen=7, delay=2, admission=1, persist=1 |
| 4 | 0.12 | 0.18/0.18/0.18/0.18 | 4 | screen=7, delay=2, admission=1, persist=1 |
| 5 | 0.15 | 0.25/0.25/0.25/0.25 | 4 | screen=7, delay=2, admission=1, persist=1 |

For every Step 7 band, daily_discharge_min_per_ward = 1 and num_days = 60. Thus Step 7 changes admission-importation and turnover diversity, not the surveillance policy or temporal span.

##### A.7 Step 8 and Step 9 extended-benchmark parameters

The current pipeline extends the canonical benchmark with a complete distribution-shift benchmark (Step 8) and a baseline-to-shift transfer probe (Step 9). In the current driver, Step 8 is defined directly by the active CONFIG["STEP8"] block. The executable shifted benchmark documented below uses 360-day shifted simulations, two shifted training families, two shifted test families, explicit CS/CR/IS/IR admission-importation weights, superspreader contact amplification, and temporal modulation of admission importation. The canonical surveillance policy, screen_every_k_days = 7, screen_result_delay_days = 2, screen_on_admission = 1, and persist_observations = 1, is held fixed inside Step 8.

##### Global Step 8 settings

Table 9: Global settings for the active Step 8 complete distribution-shift benchmark.

| Parameter | Value | Meaning |
|---|---|---|
| step8_num_days | 360 | Number of days per shifted simulation under the active Step 8 configuration. |
| step8_n_sims_per_trajectory | 12 | Number of shifted simulation replicates generated for each Step 8 trajectory family. |
| step8_train_families | 2 | Number of shifted training trajectory families pooled into synthetic_step8_shift_train. |
| step8_test_families | 2 | Number of shifted test trajectory families pooled into synthetic_step8_shift_test. |
| step8_uses_superspreader | True | All active Step 8 families inject superspreader-driven contact amplification. |
| step8_uses_seasonality | True | All active Step 8 families use either piecewise or sinusoidal admission-importation seasonality. |
| step8_max_majority_frac | 0.70 | Pre-training label-balance gate for shifted train/test cohorts. |
| step8_continue_to_step9_on_balance_failure | True | If the Step 8 balance gate fails, the current driver can skip Step 8 training/evaluation while retaining the shifted test folder for the Step 9 transfer probe. |

##### Active Step 8 shifted trajectory families

Table 10: Active Step 8 shifted trajectory families used for the complete distribution-shift benchmark.

| Trajectory | seed_base | p_admit_import CS/CR/IS/IR | daily_discharge_frac | daily_discharge_min_per_ward | Seasonality |
|---|---|---|---|---|---|
| shift_train_endog_a | 8100 | 0.010/0.010/0.010/0.010 | 0.04 | 0 | piecewise |
| shift_train_import_a | 9100 | 0.20/0.20/0.20/0.20 | 0.12 | 1 | sinusoid |
| shift_test_endog_a | 10100 | 0.012/0.012/0.012/0.012 | 0.05 | 0 | piecewise |
| shift_test_import_a | 11100 | 0.140/0.140/0.140/0.140 | 0.10 | 1 | sinusoid |

##### Step 8 mechanism-specific modifiers

Table 11: Mechanism-specific modifiers applied in the active Step 8 shifted trajectory families.

| Trajectory | Modifier | Active values |
|---|---|---|
| shift_train_endog_a | superspreader | staff=s0, state=IR, start=30, end=240, patient_frac_mult=4.0, patient_min_add=15, staff_contacts=80, edge_weight_mult=2.5 |
| shift_train_endog_a | piecewise seasonality | period=360, high_start=60, high_end=180, high_mult=1.2, low_mult=0.8, pmax_cs=1.0, pmax_cr=1.0, pmax_is=1.0, pmax_ir=1.0 |
| shift_train_import_a | superspreader | staff=s1, state=IR, start=90, end=330, patient_frac_mult=3.5, patient_min_add=12, staff_contacts=70, edge_weight_mult=2.0 |
| shift_train_import_a | sinusoid seasonality | period=360, amp=0.30, phase_day=30, pmax_cs=1.0, pmax_cr=1.0, pmax_is=1.0, pmax_ir=1.0 |
| shift_test_endog_a | superspreader | staff=s0, state=IR, start=45, end=270, patient_frac_mult=4.5, patient_min_add=18, staff_contacts=90, edge_weight_mult=2.6 |
| shift_test_endog_a | piecewise seasonality | period=360, high_start=75, high_end=210, high_mult=1.3, low_mult=0.8, pmax_cs=1.0, pmax_cr=1.0, pmax_is=1.0, pmax_ir=1.0 |
| shift_test_import_a | superspreader | staff=s1, state=IR, start=120, end=345, patient_frac_mult=4.0, patient_min_add=15, staff_contacts=80, edge_weight_mult=2.2 |
| shift_test_import_a | sinusoid seasonality | period=360, amp=0.35, phase_day=45, pmax_cs=1.0, pmax_cr=1.0, pmax_is=1.0, pmax_ir=1.0 |

##### Step 8 and Step 9 training logic

When lightweight tuning is enabled, Step 8 performs a validation-only hyperparameter search before shifted training, whereas Step 9 never tunes or retrains. Step 8 validates the shifted benchmark using the configured majority-fraction gate before shifted-model training. Step 9 first resolves the archived Step 4 baseline checkpoint for each active temporal window and horizon, then evaluates that frozen model directly on the shifted Step 8 test cohort. This makes Step 9 a strict transfer experiment rather than a re-estimation experiment.

##### Step 9 baseline-to-shift transfer protocol

Table 12: Active Step 9 baseline-to-shift transfer protocol.

| Component | Value | Meaning |
|---|---|---|
| Step 9 training data folder | BASELINE_TRAIN_REL | Step 9 reuses the canonical baseline training folder only as the task-compatible data context required by the evaluation wrapper. |
| Step 9 evaluation mode | no_train=True | Step 9 is always executed as evaluation-only; the baseline model is not retrained on shifted data. |

| Component | Value | Meaning |
|---|---|---|
| Checkpoint source | step4_baseline/trained_model.pt | For each active T and horizon, Step 9 loads the archived Step 4 baseline checkpoint. |
| Evaluation test folder | STEP8_TEST_REL | The baseline model is evaluated directly on the shifted Step 8 external test cohort. |
| Archived test folder | STEP8_TEST_REL | Archived test copies for Step 9 correspond to the shifted Step 8 test folder, not the canonical frozen benchmark. |
| Interpretation | zero-shot transfer | Step 9 measures how much predictive structure learned under the canonical benchmark transfers to the shifted external benchmark without adaptation. |

##### A.8 Path and pipeline constants

Table 13 lists the deterministic folder names and pipeline constants used to organise benchmark outputs. These values are not scientific parameters in the same sense as the simulator settings, but they matter for exact reproducibility of the workflow and for interpreting archived outputs.

Table 13: Path and pipeline constants used by the experimental workflow.

| Constant | Value | Meaning |
|---|---|---|
| DEFAULT_RESULTS_PARENT | experiments_results | Deterministic root directory for all experimental outputs. |
| DEFAULT_OVERLEAF_DIRNAME | overleaf_package | Default directory name for Overleaf export package. |
| TRAINING_OUT_REL | training_outputs | Default relative folder for training outputs. |
| CANONICAL_TRAJECTORY_NAMES | [endog_high_train, import_high_train, endog_high_test, import_high_test] | The four canonical Step 1 trajectory families. |
| CANONICAL_TRAIN_TRAJECTORIES | [endog_high_train, import_high_train] | Canonical train trajectory families. |
| CANONICAL_TEST_TRAJECTORIES | [endog_high_test, import_high_test] | Canonical test trajectory families. |
| BASELINE_TRAIN_REL | synthetic_amr_graphs_train | Pooled baseline training PT dataset. |
| BASELINE_TEST_REL | synthetic_amr_graphs_test_frozen | Frozen baseline test PT dataset. |
| LIVE_TEST_REL | synthetic_amr_graphs_test | Live test path expected by the trainer; restored from the frozen baseline before each run. |
| STEP5_ABLATION_REL | step5_ablation | Folder holding Step 5 ablation datasets. |
| STEP8_TRAIN_REL | synthetic_step8_shift_train | Pooled Step 8 shifted training PT dataset. |
| STEP8_TEST_REL | synthetic_step8_shift_test | Pooled Step 8 shifted test PT dataset. |
| STEP6_DELAY_GLOB | synthetic_endog_import_step6_delay*_pt[_flat] | Search patterns used to locate Step 6.1 delay datasets in PT or PT-flat form. |
| STEP6_FREQ_GLOB | synthetic_endog_import_step6_freq*_pt[_flat] | Search patterns used to locate Step 6.2 frequency datasets in PT or PT-flat form. |
| STEP7_SWEEP_GLOB | synthetic_endog_import_step7c_sweep*_pt[_flat] | Search patterns used to locate Step 7 sweep datasets in PT or PT-flat form. |

| Constant | Value | Meaning |
|---|---|---|
| DATASET_FIGURES_REL | _dataset_graph_figures | Archive folder for raw-graph summary figures. |

##### A.9 Command-line arguments

Table 14 provides the exposed command-line interface for running the benchmark pipeline. This is useful operationally because many of the comparisons discussed in the main text correspond directly to different combinations of these arguments.

Table 14: Command-line arguments supported by the benchmark pipeline.

| CLI argument | Default | Meaning |
|---|---|---|
| –start | "1" | First pipeline step to execute. Accepts step labels such as 1, 6.2, 8, and 9. |
| –stop | "9" | Last pipeline step to execute. |
| –dry_run | False | Validate setup and report intended actions without executing them. |
| –keep_graphml | False | If set, preserve GraphML files instead of purging them after conversion/collection. |
| –keep_step_train_graphml | False | If set, keep only the GraphML datasets needed to preserve each generated train set per step and one frozen test set per track. |
| –run_both_state_modes | False | Runs both ground_truth and partial_observation tracks. |
| –archive_train_test_folders | False | Archive copies of the train and test PT folders for each run. |
| –run_graph_folder_figures | False | Enables execution of graph-folder summary figures before GraphML purge. |
| –emit_latex | False | Export Overleaf-ready figure package and latex.txt. |
| –emit_latex_only | False | Skip the full pipeline and only rebuild the Overleaf package from an already completed results root. |
| –no_train | False | Reuse existing trained checkpoints and rerun evaluation/export logic only; all data-generation and conversion stages are skipped. |
| –no_simulation | False | Reuse all existing datasets and skip all simulation/data-generation/conversion stages while still retraining/evaluating requested models. |
| –overleaf_dir | overleaf_package | Output directory for the Overleaf package. |
| –test_frac_per_class | 0.5 | Legacy compatibility flag; canonical baseline construction ignores it. |
| –run_all_horizons | False | Train and evaluate over multiple horizons rather than one. |
| –horizons | "" | Optional comma-separated list of horizons overriding the default configuration. |
| –run_all_T | False | Train and evaluate over multiple temporal window lengths T. |
| –T_list | "" | Optional comma-separated list of T values overriding the default configuration. |
| –results_parent | experiments_results | Root folder into which deterministic experiment outputs are written. |

| CLI argument | Default | Meaning |
|---|---|---|
| –tune_step4 | False | Enables lightweight validation-only hyperparameter tuning before Step 4 baseline training. |
| –tune_step8 | False | Enables lightweight validation-only hyperparameter tuning before Step 8 shifted training. |
| –tune_trials_quick | script default | Number of quick-search trials used during lightweight tuning. |
| –tune_finalists | script default | Number of finalist configurations promoted from quick search to the longer tuning stage. |
| –tune_quick_epochs | script default | Epoch budget for quick-search screening runs. |
| –tune_full_epochs | script default | Epoch budget for finalist tuning runs. |
| –tune_split_seed | script default | Random seed controlling the validation split used during lightweight tuning. |
| –timestamped | False | Retained for compatibility but effectively ignored; deterministic result roots are used. |
| –no_timestamp | False | Retained for compatibility but effectively ignored; deterministic result roots are used. |

##### A.10 Runtime environment variables

Finally, Table 15 lists the runtime environment variables injected into the workflow. These variables determine how simulation, conversion, state-mode selection, and output locations are passed across pipeline stages.

Table 15: Runtime environment variables used across pipeline stages.

| Environment variable | Value/source | Meaning |
|---|---|---|
| DT_SIM_EXTRA_ARGS | built from simulation settings | Serialized simulator arguments passed to the data generator. |
| DT_CONVERT_EXTRA_ARGS | built from conversion settings and active horizons | Serialized conversion arguments passed to the graph-to-tensor stage. |
| DT_KEEP_GRAPHML | "1" if –keep_graphml, else "0" | Controls GraphML cleanup: purge is skipped when –keep_graphml is set. |
| DT_STATE_MODE | ground_truth or partial_observation | Selects the state-mode track being executed. |
| DT_PT_OUT_DIR | track-specific path | Scratch/output PT directory for a given track. |
| PYTHONPATH | work_dir | Set so staged per-track code is importable during execution. |
| PYTHONUTF8 | 1 | Forces UTF-8 mode for subprocesses. |
| PYTHONIOENCODING | utf-8 | Forces UTF-8 stdout/stderr encoding in subprocesses. |

### B Parameter catalogue for the causal experiment

#### B.1 Configuration parameters

Table 16 summarises the high-level dataset-construction parameters used in the current causal workflow, including dataset naming, intervention specifications, explicit split seeds, branching schedule, and the simulator regime used to generate action-conditioned decision states.

Table 16: High-level configuration parameters for the causal policy experiment.

| Parameter | Value | Meaning |
|---|---|---|
| out_name | ”causal_policy_monthly_shock_superspreader” | Name of the causal-policy dataset folder. |
| candidate_interventions_json | ”candidate_interventions.json” | Intervention specification listing the candidate actions evaluated at each decision state. |
| baseline_intervention_json | ”” | Optional non-empty baseline intervention. Empty here, so the baseline corresponds to no additional action. |
| include_baseline | True | Includes the baseline action alongside the candidate interventions. |
| use_balanced_base_families | False | The causal dataset is generated directly from explicit seed ranges rather than from balanced base families. |
| train_seeds | ”4000:4019” | Simulator seeds used for the causal training split. |
| val_seeds | ”5000:5003” | Simulator seeds used for the causal validation split. |
| test_seeds | ”6000:6009” | Simulator seeds used for the causal held-out test split. |
| window_T | 7 | Length of the pre-intervention temporal input window. |
| horizons | ”7” | Prediction horizon used during conversion and policy-target extraction. |
| decision_days | ”7:60:7” | Decision schedule; actions branch every 7 days from day 7 to day 49. |
| decision_stride | 1 | Auxiliary stride parameter used when parsing decision-day specifications. |
| action_start_mode | ”branch_at_decision_day” | Causal branching mode in which all actions share the same pre-intervention history and diverge only at the decision state. |
| decision_applies_from | ”next_day” | Applies the chosen action from the day immediately after the decision point. |
| force | True | Overwrites any existing dataset folder before rebuilding. |
| require_two_class_splits | False | Disables the binary two-class split check because the causal task is not a conventional two-class endpoint. |
| jobs | 4 | Number of parallel jobs used during causal dataset construction. |
| shared_noise_seed_offset | 910000 | Base offset used to construct the shared-noise seeds that align exogenous stochastic streams across counterfactual branches from the same state. |
| sim_args.num_days | 60 | Total simulation length used when generating each causal base trajectory. |
| sim_args.num_regions | 1 | Number of hospital regions in the simulator. |

| Parameter | Value | Meaning |
|---|---|---|
| sim_args.num_wards | 5 | Number of wards in the hospital simulation. |
| sim_args.num_patients | 50 | Number of patient agents. |
| sim_args.num_staff | 100 | Number of staff agents. |
| sim_args.staff_wards_per_staff | 2 | Number of wards covered per staff member in the simulated hospital. |
| sim_args.export_yaml | False | Disables YAML export during simulator runs. |
| sim_args.export_gif | False | Disables animated GIF export during simulator runs. |
| sim_args.isolation_mult | 0.8 | Baseline isolation multiplier before any branched intervention is applied. |
| sim_args.isolation_days | 3 | Baseline isolation duration before any branched intervention is applied. |
| sim_args.p_import_cs | 0.3 | Day-0 patient CS seeding weight defined explicitly by the causal driver. |
| sim_args.p_import_cr | 0.1 | Day-0 patient CR seeding weight defined explicitly by the causal driver. |
| sim_args.p_import_is | 0.3 | Day-0 patient IS seeding weight defined explicitly by the causal driver. |
| sim_args.p_import_ir | 0.1 | Day-0 patient IR seeding weight defined explicitly by the causal driver. |
| Fixed staff day-0 CS seeding | 0.01 | Fixed simulator probability that a staff member is initialised in CS at day 0. |
| Fixed staff day-0 CR seeding | 0.003 | Fixed simulator probability that a staff member is initialised in CR at day 0; a successful CR draw overwrites a CS draw. |
| sim_args.p_admit_import_cs | 0.3 | Baseline CS admission-importation weight used by the joint admission sampler. |
| sim_args.p_admit_import_cr | 0.4 | Baseline CR admission-importation weight used by the joint admission sampler. |
| sim_args.p_admit_import_is | 0.3 | Baseline IS admission-importation weight used by the joint admission sampler. |
| sim_args.p_admit_import_ir | 0.4 | Baseline IR admission-importation weight used by the joint admission sampler. |
| sim_args.daily_discharge_frac | 0.30 | Fraction of patients discharged each simulated day. |
| sim_args.daily_discharge_min_per_ward | 3 | Minimum daily discharges enforced per ward. |
| sim_args.screen_every_k_days | 7 | Baseline ward-wide screening frequency in days. |
| sim_args.screen_on_admission | 0 | Baseline configuration disables admission screening. |
| sim_args.screen_result_delay_days | 3 | Baseline delay between screening and observed result. |
| sim_args.persist_observations | 1 | Observed positives persist in the surveillance representation. |
| sim_args.beta_res_mult | 1.0 | Resistant phenotype multiplier used in transmission; neutral in the current causal workflow. |
| sim_args.beta_state_mult_cs | 0.5 | Source-state transmission multiplier for susceptible colonisation. |
| sim_args.beta_state_mult_cr | 0.5 | Source-state transmission multiplier for resistant colonisation. |
| sim_args.beta_state_mult_is | 1.0 | Source-state transmission multiplier for susceptible infection. |

| Parameter | Value | Meaning |
|---|---|---|
| sim_args.beta_state_mult_ir | 1.0 | Source-state transmission multiplier for resistant infection. |
| sim_args.crossward_hand_hygiene_enabled | 1 | Enables leaky hand-hygiene attenuation for cross-ward staff–patient transmission. |
| sim_args.crossward_hand_hygiene_mult | 0.55 | Multiplicative attenuation applied to eligible cross-ward staff–patient transmission contributions. |
| sim_args.passive_screen_capacity_per_week | 0 | Weekly passive-screening capacity. A value of zero denotes no capacity cap, so all eligible individuals on a scheduled screening day may be tested. |
| sim_args.active_symptom_screening | 1 | Enables active testing when a patient progresses from carriage to infection. |
| sim_args.admission_screen_symptomatic_only | 1 | Restricts admission screening to patients already in symptomatic infection states when admission screening is enabled. |
| sim_args.isolation_trigger_policy | "resistant_only" | Baseline isolation trigger policy before candidate actions are applied. |
| sim_args.admit_import_seasonality | "shock" | Admission-importation forcing regime used for the causal simulator. |
| sim_args.admit_import_shock_min_days | 20 | Minimum duration of the importation shock window. |
| sim_args.admit_import_shock_max_days | 30 | Maximum duration of the importation shock window. |
| sim_args.admit_import_shock_mult_min | 2.0 | Minimum importation multiplier during the shock window. |
| sim_args.admit_import_shock_mult_max | 3.0 | Maximum importation multiplier during the shock window. |
| sim_args.superspreader_staff | "s1" | Staff identifier used for the superspreader episode. |
| sim_args.superspreader_state | "CR" | State associated with the superspreader mechanism. |
| sim_args.superspreader_start_day | 14 | First day of the superspreader episode. |
| sim_args.superspreader_end_day | 21 | Final day of the superspreader episode. |
| sim_args.superspreader_patient_frac_mult | 2 | Multiplier on patient contact fraction during the superspreader window. |
| sim_args.superspreader_patient_min_add | 1 | Minimum additional patient contacts during the superspreader window. |
| sim_args.superspreader_staff_contacts | 30 | Staff-contact intensity during the superspreader episode. |
| sim_args.superspreader_edge_weight_mult | 2 | Edge-weight amplification during the superspreader episode. |

##### B.2 Model and training parameters

Table 17 summarises the model architecture, optimisation settings, action-conditioning controls, and loss configuration used in the causal policy experiment. In the current workflow, training is framed primarily as state-wise oracle-action classification, with an additional within-state ranking regularizer on the continuous gain target.

Table 17: Model and training parameters for the causal policy experiment.

| Parameter | Value | Meaning |
| --- | --- | --- |
| `require_complete_action_set` | `True` | Retains only decision states with one valid row per candidate action during grouped policy training. |
| `task` | `”oracle_best_action_h7”` | Horizon-7 grouped policy task in which the target is the oracle-best action for each decision state. |
| `T` | `7` | Temporal input window length used by the trainer. |
| `sliding_step` | `1` | Sliding-window stride used when constructing temporal samples from the policy manifest. |
| `use_action_conditioning` | `True` | Enables intervention-conditioned prediction. |
| `hidden` | `128` | Hidden embedding dimension of the graph-temporal model. |
| `heads` | `2` | Number of attention heads in the temporal encoder. |
| `dropout` | `0.2` | Dropout probability. |
| `transformer_layers` | `2` | Number of Transformer layers in the temporal encoder. |
| `sage_layers` | `4` | Number of GraphSAGE layers used in the graph encoder. |
| `batch_size` | `8` | Batch size argument supplied to the trainer. |
| `epochs` | `100` | Number of training epochs. |
| `lr` | `1e-4` | Learning rate for optimisation. |
| `neighbor_sampling` | `False` | Disables GraphSAGE-style neighbour sampling; the current causal workflow trains in full-graph mode. |
| `num_neighbors` | `”20,10”` | Retained in the configuration but inactive because neighbour sampling is disabled. |
| `seed_count` | `256` | Retained in the configuration but inactive because neighbour sampling is disabled. |
| `seed_strategy` | `”random”` | Retained in the configuration but inactive because neighbour sampling is disabled. |
| `seed_batch_size` | `64` | Retained in the configuration but inactive because neighbour sampling is disabled. |
| `max_sub_batches` | `4` | Retained for sampled-sub-batch control but inactive in the present full-graph workflow. |
| `max_neighbors` | `0` | Disables the legacy edge-thinning fallback path. |
| `emit_translational_figures` | `False` | Disables translational attribution figure export during the main causal-policy training run. |
| `out_name` | `”policy_training_large_ns”` | Name of the training-output folder. |
| `oracle_best_action_loss` | `True` | Enables the state-wise softmax cross-entropy loss against the simulator-defined oracle-best action. |
| `oracle_best_action_loss_weight` | `1` | Weight applied to the oracle-best-action loss. |
| `aux_policy_loss` | `False` | Disables the auxiliary gain-matching loss path. |
| `aux_policy_target_name` | `”y_h7_trans_import_res_gain”` | Continuous gain quantity retained for bookkeeping and policy evaluation. |
| `aux_policy_loss_weight` | `2` | Auxiliary loss weight; retained in the configuration but inactive because `aux_policy_loss = False`. |
| `pairwise_policy_ranking_loss` | `True` | Enables a within-state pairwise/listwise ranking regularizer on the continuous gain target. |

| Parameter | Value | Meaning |
|---|---|---|
| pairwise_policy_ranking_weight | 1 | Weight applied to the within-state ranking regularizer. |
| pairwise_policy_margin | 0.02 | Pairwise ranking margin used inside the within-state ranking regularizer. |
| pairwise_policy_min_target_gap | 0.01 | Minimum oracle-target gap required before a pair contributes to the ranking loss. |
| action_hidden_dim | 256 | Hidden dimension used to project action features before interaction with the state embedding. |
| action_interaction_hidden_dim | 256 | Hidden dimension of the action-conditioned prediction head. |
| action_interaction_dropout | 0.1 | Dropout inside the action-conditioned prediction head. |

##### B.3 Feature construction for the causal policy model

Each supervised causal-policy row comprised a length-$T$ sequence of daily contact graphs, an explicit state-summary vector extracted from the final day of the observed window, and a fixed-width descriptor of the candidate intervention applied after the decision point. Let $G_t = (V_t, E_t)$ denote the daily contact graph at day $t$. For every directed edge $(i, j) \in E_t$, the raw edge attribute was unchanged from the predictive branch and remained

$$e_{ij,t} = (w_{ij,t}, \tau_{ij,t}) \in \mathbb{R}^2,$$

where $w_{ij,t}$ is the contact weight and $\tau_{ij,t}$ is the edge-type indicator. No additional edge channels were introduced for the causal-policy experiments.

For scalar normalisation we used the clipped map

$$\nu_c(z) = \frac{\min\{\max(z, 0),\ c\}}{c},$$

with the corresponding cap $c$ stated below. Day-valued quantities were therefore mapped to $[0, 1]$ by clipping at the chosen cap. For strictly positive multiplier-like quantities, including the isolation transmission multiplier and intervention-specific multiplicative effects, we used the neutral-centred map

$$\ell(z) = \log z,$$

so that the neutral value $z = 1$ is encoded as 0, values below 1 map to negative values, and values above 1 map to positive values. When a multiplier parameter was absent for a given intervention family, the encoder used the neutral value $z = 1$, so the encoded feature remained 0. Intervention start and end days were normalised by 365.

In the partial-observation track, each node $i \in V_t$ was represented by a 16-dimensional vector

$$x^{\mathrm{PO}}_{i,t} = \big[r_i,\ o^{+}_{i,t},\ a_{i,t},\ q_{i,t},\ 0,\ 0,\ \tilde{w}_{i,t},\ \tilde{c}_{i,t},\ o^{\mathrm{known}}_{i,t},\ s^{\mathrm{today}}_{i,t},\ \nu_{30}(d^{\mathrm{last\_test}}_{i,t}),\ \nu_{30}(d^{\mathrm{pending}}_{i,t}),\ m^{\mathrm{adm}}_{i,t},\ p_{i,t},\ \nu_{30}(\ell^{\mathrm{iso}}_{i,t}),\ \nu_{30}(a^{\mathrm{adm}}_{i,t})\big].$$

Here $r_i$ indicates staff versus patient role, $o^{+}_{i,t}$ is observed positivity, $a_{i,t}$ is antibiotic exposure, and $q_{i,t}$ is current isolation status. The fifth and sixth positions are retained for dimensional compatibility with the ground-truth incident-event slots but are fixed to zero in the partial-observation track. Thus the partial-observation encoder does not feed the latent same-day new resistant acquisition and resistant infection indicators $n^{\mathrm{CR}}_{i,t}$ and $n^{\mathrm{IR}}_{i,t}$. The remaining coordinates are as follows: $\tilde{w}_{i,t}$ is ward identity normalised by the maximum ward index present in the graph, $\tilde{c}_{i,t}$ is the ward-cover count normalised by the maximum cover count in the graph, $o^{\mathrm{known}}_{i,t}$ indicates whether surveillance status is currently known, $s^{\mathrm{today}}_{i,t}$ indicates whether the individual was screened on day $t$, $d^{\mathrm{last\_test}}_{i,t}$ is the number of days since the last test, $d^{\mathrm{pending}}_{i,t}$ is the remaining delay for a pending test result, $m^{\mathrm{adm}}_{i,t}$ indicates whether admission screening is currently required, $p_{i,t}$ indicates presence in the hospital on day $t$, $\ell^{\mathrm{iso}}_{i,t}$ is the remaining isolation duration, and $a^{\mathrm{adm}}_{i,t}$ is admission age in days. When admission day was unavailable, $\nu_{30}(a^{\mathrm{adm}}_{i,t})$ was set to 1.

In the ground-truth track, the node representation was expanded to 22 dimensions:

$$x^{\mathrm{GT}}_{i,t} = \big[r_i,\ \mathbf{1}\{c_{i,t}=0\},\ \mathbf{1}\{c_{i,t}=1\},\ \mathbf{1}\{c_{i,t}=2\},\ \mathbf{1}\{c_{i,t}=3\},\ \mathbf{1}\{c_{i,t}=4\},\ a_{i,t},\ q_{i,t},\ n^{\mathrm{CR}}_{i,t},\ n^{\mathrm{IR}}_{i,t},$$
$$\tilde{w}_{i,t},\ \tilde{c}_{i,t},\ o^{+}_{i,t},\ o^{\mathrm{known}}_{i,t},\ s^{\mathrm{today}}_{i,t},\ \nu_{30}(d^{\mathrm{last\_test}}_{i,t}),\ \nu_{30}(d^{\mathrm{pending}}_{i,t}),\ y^{\mathrm{pending}}_{i,t},\ m^{\mathrm{adm}}_{i,t},\ p_{i,t},\ \nu_{30}(\ell^{\mathrm{iso}}_{i,t}),\ \nu_{30}(a^{\mathrm{adm}}_{i,t})\big],$$

where $c_{i,t} \in \{0,1,2,3,4\}$ is the latent AMR state and $y^{\mathrm{pending}}_{i,t}$ records the latent result attached to a pending test.

The model also received a 49-dimensional explicit state-summary vector $s_t$. This vector was computed from the final graph in the observed window and consisted of: (i) the means and standard deviations of the first six node channels; (ii) staff and patient fractions; (iii) burden moments comprising the mean, standard deviation, minimum, maximum, and positivity fraction of the state variable together with the fractions on antibiotics and in isolation; (iv) per-node rates of new colonisation and new infection computed from the active-track incident slots, which are masked to zero in the partial-observation track; (v) graph size and connectivity summaries given by the train-calibrated clipped log features

$$\min\left\{\frac{\log(1+|V_t|)}{Q^{\mathrm{train}}_{0.95}[\log(1+|V|)]},\,1\right\},\qquad \min\left\{\frac{\log(1+|E_t|)}{Q^{\mathrm{train}}_{0.95}[\log(1+|E|)]},\,1\right\},\qquad \min\left\{\frac{\log(1+\bar{d}_t)}{Q^{\mathrm{train}}_{0.95}[\log(1+\bar{d})]},\,1\right\},$$

together with directed density $|E_t|/|V_t|^2$, where $\bar{d}_t = |E_t|/\max(|V_t|,1)$ and the empirical $95^{\mathrm{th}}$ percentiles are computed on the training split only; (vi) edge summaries given by the squashed mean and standard deviation of edge weight,

$$\frac{\bar{w}_t}{1+\bar{w}_t},\qquad \frac{\sigma(w_t)}{1+\sigma(w_t)},$$

together with the normalized Shannon entropy of the edge-type distribution,

$$H^{\mathrm{edge}}_t = \begin{cases} -\dfrac{1}{\log K_t}\sum_{k=1}^{K_t} p_{t,k}\log p_{t,k}, & K_t \ge 2,\\ 0, & K_t = 1,\end{cases}$$

where $K_t$ is the number of edge types with non-zero empirical mass on day $t$ and $p_{t,k}$ is the empirical proportion of edge type $k$; (vii) testing-and-isolation summaries given by the means of observed positivity, observation-known status, screened-today status, days since last test, pending-test countdown, pending-test result, admission-screening requirement, presence status, remaining isolation days, and admission age; and (viii) a 9-dimensional operational-context block

$$g_t = \big[\nu_{30}(k_t),\ \nu_7(\phi_t),\ u_t,\ \nu_{30}(\delta_t),\ \ell(\eta_t),\ \nu_{30}(\lambda_t),\ \rho_t,\ \chi_t,\ \nu_{30}(\omega_t)\big],$$

where $k_t$ is the active screening cadence, $\phi_t$ is the weekly screening phase, $u_t$ indicates whether admission screening is active, $\delta_t$ is the reporting delay, $\eta_t$ is the isolation transmission multiplier, $\lambda_t$ is the isolation duration, $\rho_t$ indicates whether observed surveillance statuses persist across days in the absence of fresh testing, $\chi_t$ indicates whether day $t$ is a screening day, and $\omega_t$ is the time until the next scheduled screen. Thus

$$s_t = [\,\text{node moments, burden and graph summaries, testing summaries}, g_t\,] \in \mathbb{R}^{49}.$$

Each candidate intervention $a$ was encoded by a fixed 33-dimensional action descriptor $h_a$. This comprised a 10-dimensional intervention-family one-hot block (baseline, screening-frequency change, screening-delay change, isolation disablement, isolation-parameter change, ward-level importation reduction, cross-ward staff removal, specific-staff removal, edge removal, other), an 8-dimensional target-type one-hot block (none, global, ward, staff, edge, patient, region, hospital), and 15 scalar components:

$$h_a = \big[\text{family one-hot, target one-hot},\ b_a,\ v_a,\ \nu_{30}(f_a),\ \nu_{30}(\delta_a),\ \ell(\mu_a),\ \ell(\mu^{\mathrm{CR}}_a),\ \ell(\mu^{\mathrm{CS}}_a),\ \ell(\eta_a),\ \nu_{30}(\lambda_a),\ u_a,\ \tilde{t}^{\mathrm{start}}_a,\ \tilde{t}^{\mathrm{end}}_a,\ \kappa^{\mathrm{name}}_a,\ \kappa^{\mathrm{id}}_a,\ \kappa^{\mathrm{target}}_a\big] \in \mathbb{R}^{33},$$

where $b_a$ indicates the baseline action, $v_a$ indicates whether the action is valid for the state, $f_a$ is screening frequency, $\delta_a$ is reporting delay, $\mu_a$, $\mu^{\mathrm{CR}}_a$, and $\mu^{\mathrm{CS}}_a$ are multiplicative intervention strengths when relevant, $\eta_a$ is the encoded isolation or transmission multiplier field, falling back to the generic intervention multiplier when no explicit isolation multiplier is supplied, $\ell(\eta_a)$ is the log-encoded isolation/transmission multiplier field, $\lambda_a$ is isolation duration, $u_a$ indicates admission screening, $\tilde{t}_a^{\text{start}} = t_a^{\text{start}}/365$ and $\tilde{t}_a^{\text{end}} = t_a^{\text{end}}/365$ are normalized temporal bounds, and $\kappa_a^{\text{name}}$, $\kappa_a^{\text{id}}$, and $\kappa_a^{\text{target}}$ are lightweight hashed identifiers mapped to the unit interval to distinguish sibling actions within the same intervention family.

For compact reference, the per-node, per-graph, and per-action inputs can be written abstractly as

$$x_{i,t}^{\text{PO}} = \Big[ \underbrace{r_i,\ o_{i,t}^{+},\ a_{i,t},\ q_{i,t},\ 0,\ 0}_{\text{role, observed state, and masked incident slots (6)}},\ \underbrace{\tilde{w}_{i,t},\ \tilde{c}_{i,t}}_{\text{ward context (2)}},\ \underbrace{o_{i,t}^{\text{known}},\ s_{i,t}^{\text{today}},\ \nu_{30}(d_{i,t}^{\text{last\_test}}),\ \nu_{30}(d_{i,t}^{\text{pending}}),\ m_{i,t}^{\text{adm}}}_{\text{testing and observability (5)}},\ \underbrace{p_{i,t},\ \nu_{30}(\ell_{i,t}^{\text{iso}}),\ \nu_{30}(a_{i,t}^{\text{adm}})}_{\text{presence, isolation, and stay history (3)}} \Big] \in \mathbb{R}^{16},$$

$$x_{i,t}^{\text{GT}} = \Big[ \underbrace{r_i,\ \mathbf{1}\{c_{i,t}=0\},\dots,\mathbf{1}\{c_{i,t}=4\}}_{\text{role and latent AMR state (6)}},\ \underbrace{a_{i,t},\ q_{i,t},\ n_{i,t}^{\text{CR}},\ n_{i,t}^{\text{IR}}}_{\text{treatment, isolation, and incident burden (4)}},\ \underbrace{\tilde{w}_{i,t},\ \tilde{c}_{i,t}}_{\text{ward context (2)}},\ \underbrace{o_{i,t}^{+},\ o_{i,t}^{\text{known}},\ s_{i,t}^{\text{today}},\ \nu_{30}(d_{i,t}^{\text{last\_test}}),\ \nu_{30}(d_{i,t}^{\text{pending}}),\ y_{i,t}^{\text{pending}},\ m_{i,t}^{\text{adm}}}_{\text{testing and observability (7)}},\ \underbrace{p_{i,t},\ \nu_{30}(\ell_{i,t}^{\text{iso}}),\ \nu_{30}(a_{i,t}^{\text{adm}})}_{\text{presence, isolation, and stay history (3)}} \Big] \in \mathbb{R}^{22},$$

$$e_{ij,t} = \Big[ \underbrace{w_{ij,t}}_{\text{contact weight (1)}},\ \underbrace{\tau_{ij,t}}_{\text{edge type (1)}} \Big] \in \mathbb{R}^2, \qquad s_t = \Big[ \underbrace{\text{first-six node-channel means and standard deviations}}_{12},\ \underbrace{\text{staff/patient composition}}_{2},\ \underbrace{\text{burden moments}}_{7},\ \underbrace{\text{new-case rates}}_{2},\ \underbrace{\text{graph size/connectivity}}_{4},\ \underbrace{\text{edge summaries}}_{3},\ \underbrace{\text{testing/isolation summaries}}_{10},\ \underbrace{\text{operational context}}_{9} \Big] \in \mathbb{R}^{49},$$

$$h_a = \Big[ \underbrace{\text{intervention-family one-hot}}_{10},\ \underbrace{\text{target-type one-hot}}_{8},\ \underbrace{\text{scalar intervention parameters and metadata}}_{15} \Big] \in \mathbb{R}^{33}.$$

Thus, for either track, one supervised policy row is the tuple

$$\Big(\{(X_\tau, E_\tau, A_\tau)\}_{\tau=t-T+1}^{t},\ s_t,\ h_a\Big),$$

where $X_\tau$ is formed from $x_{i,\tau}^{\text{PO}}$ in the partial-observation track and from $x_{i,\tau}^{\text{GT}}$ in the ground-truth track, and $A_\tau$ contains the two edge channels $(w_{ij,\tau}, \tau_{ij,\tau})$. In the state-summary pathway, the 12 node-moment coordinates are computed from the first six node channels of the active track, namely $(r, o^{+}, a, q, 0, 0)$ in partial observation and $(r, \mathbf{1}\{c=0\}, \dots, \mathbf{1}\{c=4\})$ in ground truth.

##### B.4 Policy-evaluation parameters

Table 18 reports the policy-selection settings used after model training. These values determine the held-out split, the oracle-comparison target, score export, and the naming of the evaluation-output folder.

Table 18: Policy-evaluation parameters for the causal policy experiment.

| **Parameter** | **Value** | **Meaning** |
|---|---|---|
| `split` | `"test"` | Held-out split used for policy-selection evaluation. |
| `oracle_metric` | `"y_h7_trans_import_res_gain"` | Simulator-derived oracle target used to identify the best action for each held-out state. |
| `selection_direction` | `"auto"` | Automatically infers that larger values are preferable for this gain-based oracle target. |
| `emit_action_scores_csv` | `True` | Exports per-state, per-action scores for downstream constrained optimisation. |
| `out_name` | `"policy_selector_eval_large"` | Name of the policy-evaluation output folder. |

The exported per-action score tables and per-state summaries are restricted to evaluable held-out states that retain a complete action set and finite oracle values. The same filtered state set is passed unchanged to the downstream decision layer.

##### B.5 Workflow stages

Table 19 lists the four stages of the causal workflow. These stages correspond to dataset construction, action-conditioned training, held-out policy evaluation, and MILP-based constrained intervention selection.

Table 19: Execution stages of the causal workflow.

| Stage | Name | Meaning |
|---|---|---|
| `C1` | `build causal policy dataset` | Generates causal-policy trajectories under the 60-day shock-plus-superspreader regime, branches candidate interventions from shared decision states using matched shared-noise counterfactual rollout, converts the branched trajectories to tensor objects, and writes the policy manifest. |
| `C2` | `train action-conditioned predictor` | Trains the GraphSAGE+Transformer policy model with action conditioning, operational-state summaries, the state-wise oracle-best-action loss, and an active within-state ranking regularizer on the continuous gain target. |
| `C3` | `evaluate policy selector` | Scores all candidate actions on the held-out split, compares model-selected actions with simulator-oracle actions, and exports action-score tables restricted to evaluable held-out states. |
| `C4` | `optimize policy MILP` | Solves a constrained binary action-selection problem over the exported held-out action scores using baseline-delta utilities and intervention-family cost/cooldown rules. |

### C Appendix: MILP decision-layer configuration

Table 20 summarises the reduced intervention library used by the current constrained decision layer, and Table 21 summarises the corresponding optimisation parameters. These appendix tables document the proof-of-concept decision layer as used in the present causal workflow. In the current implementation, the optimisation layer operates on intervention-conditioned scores aligned to baseline-relative improvement in future transmission-plus-importation resistant burden over a 7-day horizon, using the held-out action-score export from policy evaluation and a baseline-delta score transform before optimisation. The active intervention menu contains four screening-based actions. Each action couples a screening schedule with an explicit isolation response for test-positive individuals, allowing the decision layer to compare screening cadence, result delay, and isolation trigger policy while holding the isolation intensity fixed at `isolation_mult = 0.75` for `isolation_days = 3`.

Table 20: Intervention library used by the MILP decision layer.

| Action | Current implementation |
|---|---|
| Baseline | Default policy with no additional intervention applied beyond the base simulator regime. Action identifier: `baseline`. |
| Screen 2d delay 0d + iso resistant only, mult 0.75, 3d | `set_screening_frequency`; `frequency_days = 2`, `screen_on_admission = 1`, `screen_result_delay_days = 0`, `isolation_mult = 0.75`, `isolation_days = 3`, `isolation_trigger_policy = resistant_only`. |
| Screen 7d delay 2d + iso resistant only, mult 0.75, 3d | `set_screening_frequency`; `frequency_days = 7`, `screen_on_admission = 1`, `screen_result_delay_days = 2`, `isolation_mult = 0.75`, `isolation_days = 3`, `isolation_trigger_policy = resistant_only`. |
| Screen 2d delay 0d + iso all positives, mult 0.75, 3d | `set_screening_frequency`; `frequency_days = 2`, `screen_on_admission = 1`, `screen_result_delay_days = 0`, `isolation_mult = 0.75`, `isolation_days = 3`, `isolation_trigger_policy = all_positives`. |
| Screen 7d delay 2d + iso all positives, mult 0.75, 3d | `set_screening_frequency`; `frequency_days = 7`, `screen_on_admission = 1`, `screen_result_delay_days = 2`, `isolation_mult = 0.75`, `isolation_days = 3`, `isolation_trigger_policy = all_positives`. |

Table 21: MILP decision-layer formulation and configurable parameters.

| Component | Current implementation |
|---|---|
| Objective | Maximise total predicted action utility. Under `score_transform = baseline_delta`, utility for state–action pair $(d, a)$ is the model-predicted score for $a$ minus the predicted score of the baseline action for the same state. There is no explicit cost-penalty term in the objective. |
| Selection rule | Exactly one action is selected for each retained decision state. |
| Score orientation | `selection_direction = auto`. For the current oracle metric `y_h7_trans_import_res_gain`, this resolves to maximisation, so larger predicted scores are better. |
| Utility input | Predicted state–action scores exported by the held-out policy-evaluation stage after restricting to evaluable states with complete action sets and finite oracle values, then transformed relative to baseline before optimisation. |
| Oracle reference | On held-out policy-evaluation states, oracle-best action is defined using `y_h7_trans_import_res_gain`, so better actions are those with larger baseline-relative improvement in transmission-plus-importation resistant burden. |
| Active intervention menu | Four screening-based intervention actions are evaluated, in addition to the baseline comparator. The menu varies screening cadence, result delay, and isolation trigger policy, while keeping the post-test isolation multiplier and duration fixed. |
| Default action cost | `0.0`. Actions without an explicit rule inherit zero cost. |
| Default cooldown | `0` days. Actions without an explicit rule inherit no cooldown restriction. |
| Default max uses | `null`. Actions without an explicit rule inherit no global use cap. |
| Budget | `budget_total = null` in the current configuration, so the global budget constraint is inactive unless overridden. |
| Explicit rule: baseline | `baseline` has `cost = 0.0`, `cooldown_days = 0`, and `max_uses = null`. |
| Named rule: screening family | The active intervention family `set_screening_frequency` has `cost = 1.0`, `cooldown_days = 7`, and `max_uses = null`. All four screening variants inherit this shared family rule. |
| Named rule: isolation family | The configuration also contains a `set_isolation_parameters` family rule with `cost = 1.5`, `cooldown_days = 7`, and `max_uses = null`; it is inactive under the present reduced menu because no standalone isolation action is generated. |
| Standalone isolation actions | No standalone `set_isolation_parameters` action is present in the reduced intervention menu. Isolation is represented only as a parameterised response embedded within each screening action. |
| Constraint grouping | Identifier-specific rules take precedence when present; otherwise cooldown and max-use constraints are enforced by intervention family rather than by raw action identifier. Under the reduced menu, the constrained non-baseline family is `set_screening_frequency`. |
| Cooldown constraints | Cooldown rules are enforced within each trajectory by forbidding repeated use of the same constrained family across decision states whose day difference is no greater than the family-specific cooldown. In the current setup, this binds all four screening variants jointly under a shared 7-day screening cooldown. |
| Solver | Pyomo MILP model solved with CPLEX in the default configuration; a SciPy MILP fallback is retained for robustness. |
| Time limit | `300` seconds. |
| MIP gap | `0.0`. |
